# Who funded the research behind the Oxford-AstraZeneca COVID-19 vaccine? Approximating the funding to the University of Oxford for the research and development of the ChAdOx vaccine technology

**DOI:** 10.1101/2021.04.08.21255103

**Authors:** Samuel Cross, Yeanuk Rho, Henna Reddy, Toby Pepperrell, Florence Rodgers, Rhiannon Osborne, Ayolola Eni-Olotu, Rishi Banerjee, Sabrina Wimmer, Sarai Keestra

## Abstract

**Objectives:** The Oxford-AstraZeneca COVID-19 vaccine (ChAdOx1 nCoV-19 or Vaxzevira) builds on nearly two decades of research and development (R&D) into Chimpanzee adenovirus-vectored vaccine (ChAdOx) technology at the University of Oxford. This study aims to approximate the funding for the R&D of the ChAdOx technology and the Oxford-AstraZeneca vaccine, and assess the transparency of funding reporting mechanisms.

**Design:** We conducted a scoping review and publication history analysis of the principal investigators to reconstruct the funding for the R&D of the ChAdOx technology. We matched award numbers with publicly-accessible grant databases. We filed Freedom Of Information (FOI) requests to the University of Oxford for the disclosure of all grants for ChAdOx R&D.

**Results:** We identified 100 peer-reviewed articles relevant to ChAdOx technology published between 01/2002 and 10/2020, extracting 577 mentions of funding bodies from funding acknowledgement statements. Government funders from overseas were mentioned 158 (27.4%), the U.K. government 147 (25.5%) and charitable funders 138 (23.9%) times. Grant award numbers were identified for 215 (37.3%) mentions, amounts were available in the public realm for 121 (21.0%) mentions. Based on the FOIs, until 01/2020, the European Commision (34.0%), Wellcome Trust (20.4%) and CEPI (17.5%) were the biggest funders of ChAdOx R&D. From 01/2020, the U.K. Department of Health and Social Care was the single largest funder (89.3%). The identified R&D funding was £104,226,076 reported in the FOIs, and £228,466,771 reconstructed from the literature search.

**Conclusions:** Our study identified that public funding accounted for 97.1-99.0% of the funding towards the R&D of ChAdOx and the Oxford-AstraZeneca vaccine. We furthermore encountered a severe lack of transparency in research funding reporting mechanisms.

**Strengths and limitations of this study:**

- This is the first study that analysed the R&D funding and funders contributing to the Oxford-AstraZeneca vaccine and the underlying ChAdOx technology.
- We used multiple sources and methods to approximate the R&D funding of the Oxford-AstraZeneca Vaccine and ChAdOx technology.
- We cross-matched award numbers with all publicly-accessible databases by major funders of R&D.
- Freedom Of Information requests were a useful method to identify R&D funding, but face limitations in their scope of data collection.
- Integration of the two data sets was not possible due to insufficient grant information and lack of award numbers in funding acknowledgement statements in peer-reviewed articles.

## Introduction

The ChAdOx1 nCoV-19 vaccine, commonly known as the Oxford-AstraZeneca vaccine, Covishield, or Vaxzevira, is one of three vaccines that has received conditional approval for the prevention of COVID-19 in the U.K. (March 2021)[1]. The Oxford-AstraZeneca vaccine has received regulatory approval in over 100 countries as of late March 2021, and approximately 50 million doses have been administered across the U.K., the E.U., and India[2]. This vaccine makes use of a novel technology that relies on a Chimpanzee Adenovirus-vector (ChAdOx) to encode the production of the SARS-CoV-2 spike protein, which induces an immune response[3]. The Oxford-AstraZeneca vaccine is particularly important because it does not require the same cold chain management and is more affordable than the other early-approved COVID-19 vaccines from Pfizer/BioNTech and Moderna[4]. It is therefore more feasible to distribute the vaccine for use globally, particularly in resource-limited settings[5].

Although the Oxford-AstraZeneca vaccine itself was developed in response to the COVID-19 pandemic, the underlying ChAdOx platform relies on two decades of virus-vectored vaccine research at the Jenner Institute, University of Oxford, led by Professor Sarah Gilbert (S.G.) and Professor Adrian Hill (A.H.). The ChAdOx technology has previously been trialled for other infectious diseases in human participants including Hepatitis C virus and malaria, where it was shown to induce a powerful immune response during phase 1 clinical trials[6, 7]. Before the emergence of SARS-CoV-2, the vaccine was under development for Middle East Respiratory Syndrome coronavirus (MERS-CoV)[8]. When the pandemic emerged, the ChAdOx1 MERS-CoV vaccine had already undergone clinical trials in non-human primates and humans (phase I) and was rapidly adapted to induce an immune response to SARS-CoV-2[9].

Previous studies have shown that public funding has played a significant role in the medical innovation system for many decades, particularly in vaccine research[10-12]. Between 2000 and 2019, the U.S. National Institute of Health (NIH) funded over $17.2 billion in published research on vaccine technologies, providing the foundation for the COVID-19 vaccines currently entering the market[13]. Despite a number of public statements involving funding pledges for the development of the Oxford-AstraZeneca vaccine[4], it remains largely unknown which funding bodies have contributed to the ChAdOx technology. In this study, we therefore aim to identify the funding to the University of Oxford for R&D of the ChAdOx technology with a specific focus on research conducted at the Jenner Institute, and its subsequent application to the Oxford-AstraZeneca vaccine. This study has three objectives: (i.) to approximate the funding for the R&D of the Oxford-AstraZeneca COVID-19 vaccine and the underlying ChAdOx platform, with a specific focus on research led by S.G. and A.H.; (ii.) to identify the main funders based on grants mentioned in academic publications on the ChAdOx technology and their award numbers; (iii.) to assess the transparency in R&D funding reporting mechanisms by comparing information available in the public realm with disclosures by the University of Oxford in response to Freedom Of Information requests.

## Methods

### Scoping review of the academic literature to identify primary research on ChAdOx and the Oxford-AstraZenecavaccine

We performed a scoping review of the literature using a systematic search between the 26th of October and the 30th of November 2020 to identify all relevant academic publications which included primary research involving the ChAdOx technology. Our search strategies (Supplementary File 1) were developed in collaboration with an academic librarian from Imperial College London, and included Medline and Embase database searches for all publications mentioning the ChAdOx technology. To identify further articles, we conducted a PubMed search of the complete publication history of S.G and A.H, the primary investigators of the ChAdOx technology at the Jenner Institute. Abstracts were manually screened by two independent reviewers using Rayyan QCRI[14] based on the following inclusion criteria: (i.) peer-reviewed primary research articles, (ii.) mentioning of the relevant vaccine technology as identified in preliminary background research and described in the search strategy (i.e. using the terms ChAdOx1, ChAdOx2, Chimpanzee adenovirus-vectored, etc.), (iii.) including at least one author affiliated to the University of Oxford (Supplementary File 1). Non-English studies and review articles, conference abstracts, clinical trial registry entries, and opinion pieces not containing any primary data were excluded.

### Data extraction from funding acknowledgement statements in the academic literature

The full-text of all selected articles were downloaded into Endnote version 7.8 and duplicates were removed. Two authors extracted information from all acknowledgement sections, funding statements, and conflict of interests declarations from the academic publications on the ChAdOx technology and entered them into an Excel sheet (Supplementary File 2). Firstly, we ranked funding bodies and other actors by the absolute number of mentions extracted from the included articles. Next, we quantified the proportion of grants that listed anaward number, and conducted a separate analysis in which we removed any duplicate mentions of funder names if they were linked to the same award number. Meanwhile, using the award numbers, we searched the following publicly-available databases to identify grants towards the development of the ChAdOx technology; U.K. Research and Innovation (UKRI), European Commission, Wellcome Trust, Bill & Melinda Gates Foundation, Coalition for Epidemic Preparedness Innovations (CEPI), and World Report, the latter of which includes all grants administered by the U.S. National Institutes of Health. Grants in currencies other than GBP were converted into GBP using the following conversion rates on the date 28/02/2021: 1 U.S. Dollar = 0.72 GBP, 1 Euro = 0.87 GBP[15]. Funding declarations from the academic literature were matched to grant amounts where publicly available (Supplementary File 2). Additionally, we utilised previously collected open-access data from publicmeds4covid.com, which tracks government investment in COVID-19 research[16]. Funders were categorised into the following funding types: overseas government (including the EU), U.K. government, charity, public-private partnership (PPP), research institution, and industry.

### Freedom Of Information (FOI) Requests

To gain access to the internal records held by the University of Oxford on grants received for ChAdOx R&D, we filed several requests under the Freedom Of Information Act (2000) to ask for the disclosure of all funding (including all financial support, grants, donations, etc.) for both the ChAdOx technology and the ChAdOx1 nCoV-19 vaccine. The FOIs are listed in Supplementary File 4.

#### FOI on the funding for the ChAdOx technology

On the 25th of October 2020, we filed a request under the Freedom Of Information Act (2000) to the University of Oxford for the disclosure of all funding (including all financial support, grants, donations, etc.) that had been received by the University of Oxford and the Jenner institute relating to the ChAdOx vaccine platform, from the earliest date for which information is available to the present day[17]. This request was rejected as, according to the Information Compliance Team at the University of Oxford, it exceeded the maximum amount of time (18 hours) a public authority is legally required to spend on responding to a single FOI request (19/11/2020). To limit the scope of the first part of the request, we instead requested the disclosure of all funding (including all financial support, grants, donations, etc.) to the principal investigators, Sarah Gilbert and Adrian Hill, since 2000 to the most recent date available. We received a list of relevant grants received on the 27th of January 2021.

#### FOI on the funding for the ChAdOx1 nCoV-19 vaccine

The original FOI filed on 25/10/2020 also requested disclosure of all grants received from public entities and AstraZeneca for the development of the ChAdOx1 nCoV-19 vaccine specifically since January 1, 2020 to the date of the request. We received a response from the University of Oxford disclosing all pandemic funding for ChAdOx research at the university on the 19th of November 2020. On the 5th of February 2021, we filed a further FOI request to the University of Oxford to disclose all funding, monetary or in-kind contributions, received from AstraZeneca until February 2020 for the development of the ChAdOx1 nCoV-19 vaccine. We received a response to this request on the 2nd of March 2021 [18].

## Analysis of grant disclosures by the University of Oxford

Two authors independently classified the grants into the following categories based on the project names pertaining to each grant, provided by the University of Oxford: (i.) funding towards the COVID-19 vaccine specifically, (ii.) funding towards the research & development (R&D) of the ChAdOx technology, (iii.) funding for the fellowships/salary/research equipment/infrastructure (later coded as “other vaccine research”) that may have contributed to the development of the ChAdOx technology but is not directly identifiable (not displayed) (iv.) other funding not relevant to the ChAdOx technology (not displayed). Funders were additionally categorised into the following funding types: overseas government (including the EU), U.K. government, charity, PPP, research institution, industry, and other, which included anonymous funders that could not be classified. From the first dataset, which included all grants to S.G. and A.H. from 2000 to October 2020, we analysed all grants up to January 2020 which were relevant to the R&D of the ChAdOx technology, excluding grants that mentioned COVID-19. From the second dataset, which included all grants relevant to ChAdOx1 nCoV-19 from January 2020 to October 2020, we analysed all grants that were relevant to the R&D of the Oxford-Astrazeneca vaccine specifically.

## Results

### Funding based on disclosure statements in academic publications on the ChAdOx technology

We identified 100 published peer-reviewed articles relevant to the Oxford-AstraZeneca vaccine or the ChAdOx technology (Supplementary File 1 & 2). Publication dates ranged from January 2002 to November 2020. The concordance between the two independent reviewers was 93.61%. Funding acknowledgement statements differed in completeness between articles, with some only noting funding bodies and others detailing specific grants using grant titles or award numbers. In total, we extracted 577 mentions of funding bodies, with or without reference to specific grants. Of these, we were able to identify award numbers for 215 mentions (37.3%). Grant amounts were available in the public realm for 121 mentions (21.0%). Of the 215 mentions for which we ascertained award numbers, 73 mentions (12.7% of total mentions) corresponded to a previously identified award number. These mentions were not excluded from the total number due to the low proportion of mentions for which we were able to identify award numbers. However, grants identified as being duplicates based on having the same award numbers were excluded when calculating the amount of funding provided by that funding body. The total amount of funding we were able to reconstruct was £228,466,771.

Overseas government bodies were mentioned in funding acknowledgement statements of peer-reviewed articles on ChAdOx 158 times (27.4%), followed by the U.K. government (147 mentions (25.5%)), and charities (138 mentions (23.9%)) (Table 1). Funders from industry were mentioned 6 times (1.0%), and public-private partnership (PPP) funders (including CEPI, PATH malaria vaccine initiative and CGIAR) 15 times (2.6%). Grant amounts could be matched with 27.9% of U.K. government mentions, 19.0% of overseas government (including EU) mentions, and 36% of charity mentions. Overseas government funders contributed the most funding for which grant amounts could be identified, namely £105,715,805 (46.3%). This was followed by the U.K. government who contributed £69,773,203 (30.5%) and charitable organisations who contributed £52,977,763 (23.2%) based on traceable grants that could be linked to amounts in publicly available grant databases.

**Table 1.** Number of mentions and amount of funding identified for each funder type from the academic literature identified in the scoping review.

| Funder type | Number of mentions from the literature | Percentage of mentions matched to a grant amount | Total value of matched grants |
| --- | --- | --- | --- |
| Overseas Government (including EU) | 158 (27.4%) | 19.0% | £105,715,805 (46.3%) |
| UK government | 147 (25.5%) | 27.9% | £69,773,203 (30.5%) |
| Charity | 138 (23.9%) | 36.2% | £52,977,763 (23.2%) |
| Research institution | 113 (19.6%) | 0.0% | £0 (0.0%) |
| PPP | 15 (2.6%) | 0.0% | £0 (0.0%) |
| Industry | 6 (1.0%) | 0.0% | £0 (0.0%) |
| **Total** | **577** | **21.4% of all mentions matched** | **£228,466,771** |

Table 2 provides an overview of individual funders for whom grant amounts were identified from publicly available databases, ranked based on the total number of mentions. Here, we have only displayed funders mentioned across more than 7 articles. The most frequently named funding body was the Wellcome Trust (107 (18.5%)), followed by the Jenner Institute (73 (12.7%)), the Medical Research Council (MRC) (66 (11.4%)) and the U.S. NIH (64 (11.4%)). The top three funders for which we could retrieve most grant amounts from publicly available databases to match them with funder mentions in the acknowledgement section were UKRI (72.2%), the European Commision (58.6%) and the Wellcome Trust (44.9%).

**Table 2.**
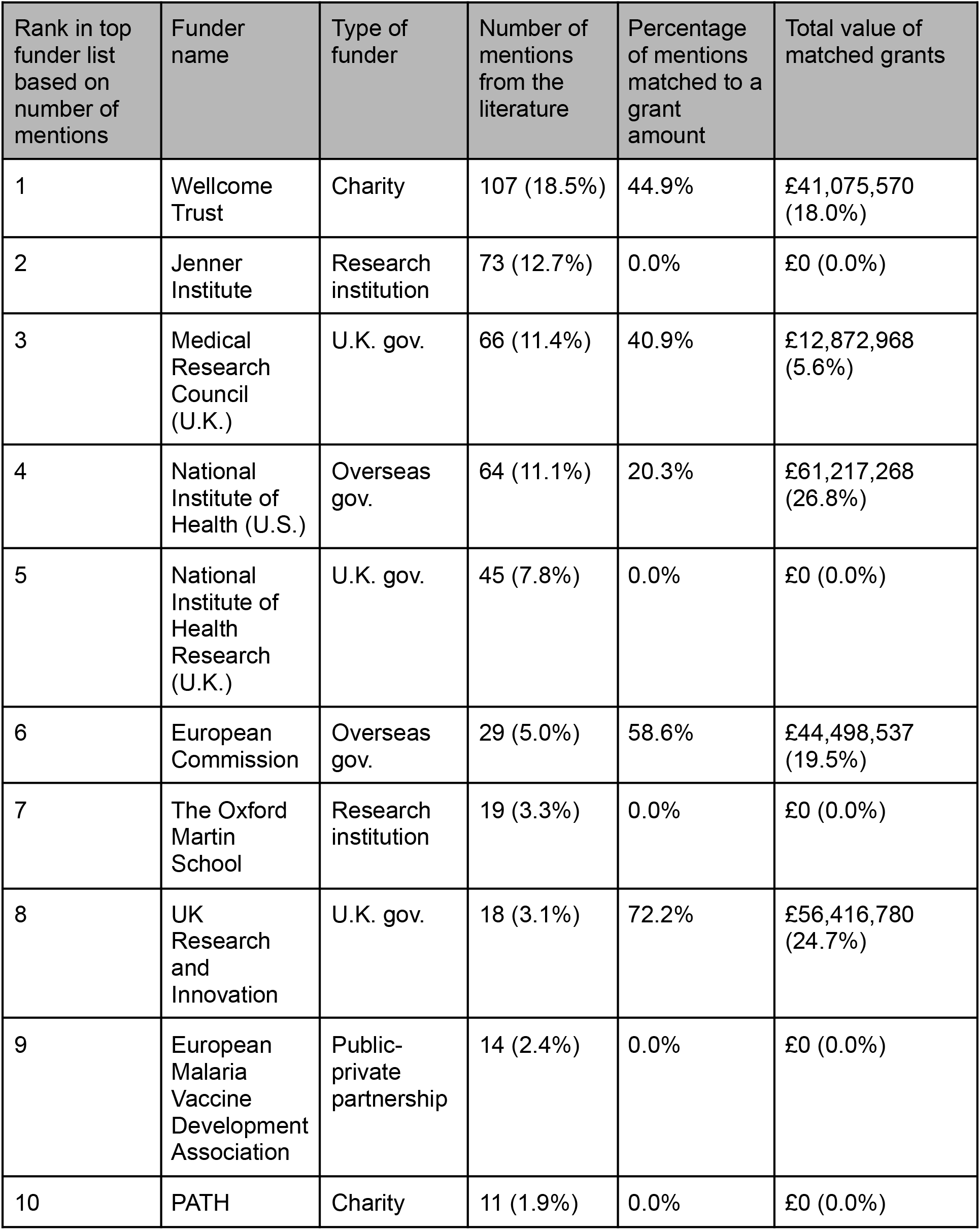
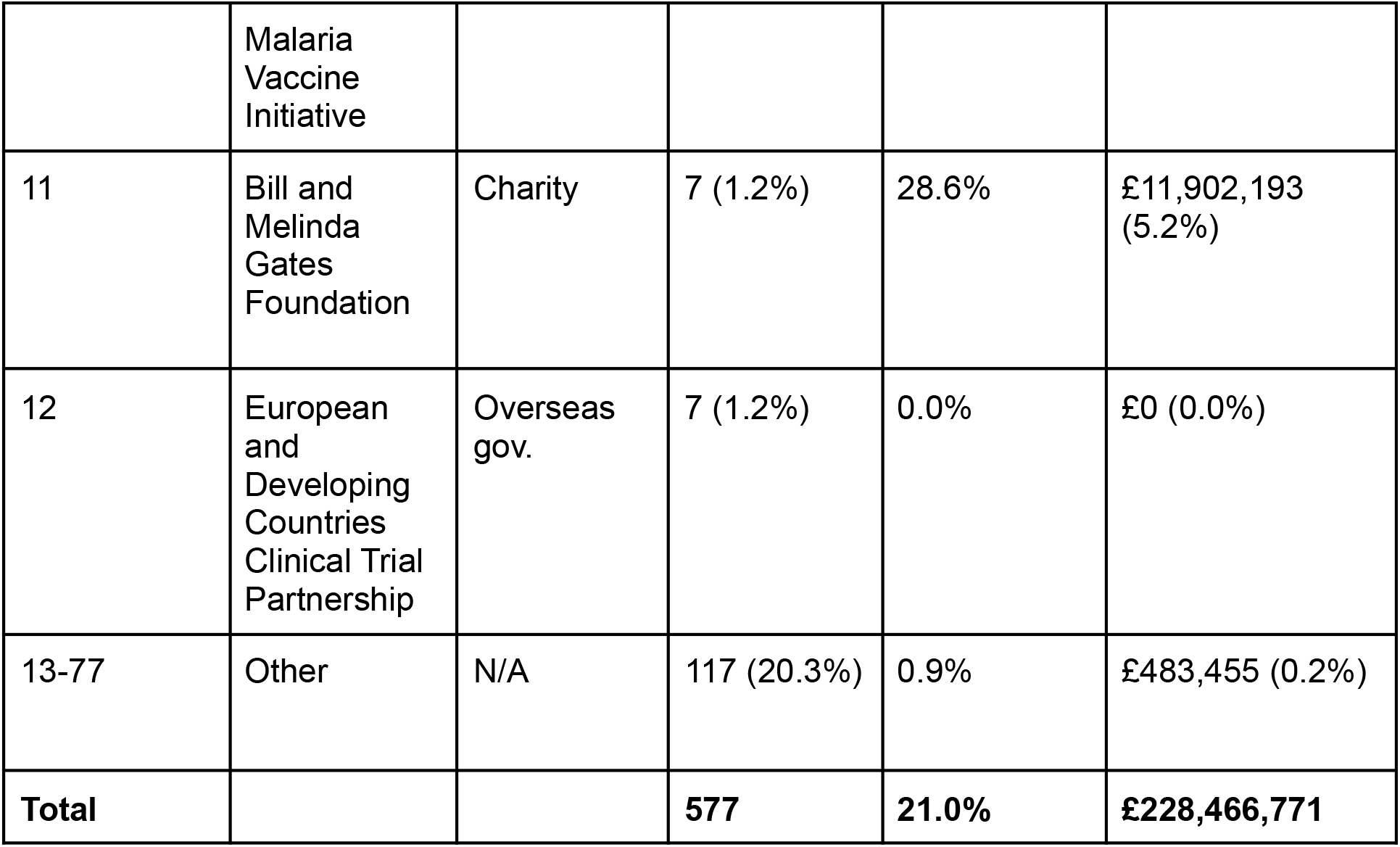
Number of mentions and amount of funding identified for the top 12 funders from the academic literature identified in the scoping review, ranked by number of mentions.

## Funding based on Freedom of Information requests to the University of Oxford

There were two datasets that the University of Oxford disclosed in response to our FOI requests, which related to pre-pandemic and pandemic grants towards ChAdOx R&D respectively. The first dataset includes all the grants that were received by S.G. and A.H since the year 2000, from which we extracted the grants relevant to the R&D of the ChAdOx technology based on the project numbers and grant names with a cut-off of January 2020. The second dataset is of grants received by the University of Oxford between January 2020 and October 2020 for the development of the Oxford-AstraZeneca vaccine. In total, the University of Oxford reported 189 grants, donations and payments to the university, 133 of which contributed to the development of the Oxford-AstraZeneca vaccine and the underlying ChAdOx technology (Table 3). The reported grants spanned a period from January 2004 until October 2020 and are included in Supplementary File 3&4. R&D of the ChAdOx technology and the Oxford-AstraZeneca vaccine at the University of Oxford cost at least £104,226,076, of which £69,313,380 was provided before January 1st 2020 and £34,912,696 on or after that date.

**Table 3.** Funding given to support the R&D of the ChAdOx technology and the Oxford-AstraZeneca vaccine based on FOIs to University of Oxford, sorted by funder type.

| Funder type | ChAdOx technology<br>(to S.G. and A.H. only) | Oxford-AstraZeneca<br>vaccine | Total |
| --- | --- | --- | --- |
| U.K. government | £5,511,316 (8.0%) | £33,354,469 (95.5%) | **£38,865,785 (37.3%)** |
| Overseas<br>government | £26,252,085 (37.9%) | £0 (0.0%) | **£26,252,085 (25.2%)** |
| Charity | £21,468,904 (31.0%) | £1,217,835 (3.5%) | **£22,686,739 (21.8%)** |
| PPP | £12,943,763 (18.7%) | £272,286 (0.8%) | **£13,216,049 (12.7%)** |
| Research<br>institution | £0 (0.0%) | £68,106 (0.2%) | **£68,106 (0.1%)** |
| Industry | £1,970,370 (2.8%) | £0 (0.0%) | **£1,970,370 (1.9%)** |
| Other | £1,166,941 (1.7%) | £0 (0.0%) | **£1,166,941 (1.1%)** |
| **Total** | **£69,313,380** | **£34,912,696** | **£104,226,076** |

The total amount of funding received for the Oxford-AstraZeneca vaccine and adenovirus technology, for each funder type, is given in the Total column.

The largest funding source for the R&D investment into the pre-pandemic ChAdOx technology research by S.G. and A.H. before January 1, 2020, was overseas governments, including the EU, which contributed £26,252,085 (37.9%). During the same period charitable funding accounted for £21,468,904 (31.0%), PPPs (including CEPI, CGIAR, and PATH malaria vaccine initiative) contributed £12,943,763 (18.7%), and the U.K. government was the fourth largest funding source with £5,511,316 (8.0%). Industry funding accounted for £1,970,370 (2.8%).

Since January 1, 2020, the largest funding source for pandemic R&D into ChAdOx for pandemic R&D into ChAdOx was the U.K. government which contributed £33,354,469 (95.5%). On or after this date, charitable funders (Wellcome Trust) accounted for £1,217,835 (3.5%), PPP, specifically CEPI, accounted for £272,286 (0.8%) and research institutions accounted for £68,106 (0.2%) of R&D funding for the Oxford-AstraZeneca vaccine.

Taking pre-pandemic and pandemic R&D funding together, the U.K. government provided £38,865,785 (37.3%) of the R&D funding for the Oxford-AstraZeneca vaccine and ChAdOx technology, making it the largest funder type identified. Overseas government ranked the second highest funder type, providing £26,252,085 (25.2%%) of R&D funding for the Oxford-AstraZeneca vaccine and ChAdOx technology, while charitable funders contributed £22,686,739 (21.8%). Industry funders contributed £1,970,370 (1.9%).

Overall, public funding sources accounted for 97.1% of the R&D funding towards the ChAdOx technology and its application to SARS-CoV-2, as disclosed by the University of Oxford per FOI. Direct government funding accounted for 62.5%, equalling a total of £65,117,870, whilst charitable sources accounted for £22,686,739 (21.8%). The PPPs CEPI and PATH malaria vaccine initiative accounted for 12.7% of R&D funding. Private industry contributed 1.9% of R&D funding, 1.2% was from other sources.

Together, the top 9 funders were responsible for 95.6% of the disclosed funding for the ChAdOx technology and the Oxford-AstraZeneca vaccine (Table 4). The remaining ten funders contributed £4,574,803 (4.4%). Of the top funders identified, three were U.K. government funders, two E.U. funders, and three charities. Before January 1, 2020, the biggest funders of the R&D into the ChAdOx technology were the European Commission (22.6%), Wellcome Trust (14.7%) and CEPI (11.9%). Since January 1, 2020, the Department of Health and Social Care was the largest funder as declared by the University of Oxford contributing 89.3% of R&D funding. The University of Oxford disclosed that they had not received any funding for the Oxford-AstraZeneca vaccine in the period from January 1st 2020 to the 5th of February 2021 (Supplementary File 3).

**Table 4.** Top 9 funders ranked by total amount of funding given to support the R&D of the ChAdOx technology and Oxford-AstraZeneca vaccine, based on FOIs to theUniversity of Oxford. Funders which contributed >£1,000,000 are shown.

| Rank based on total amount | Funder | ChAdOx technology (to S.G. and A.H. only) | Oxford-AstraZeneca vaccine | Total |
| --- | --- | --- | --- | --- |
| 1 | Department of Health and Social Care | £0 (0.0%) | £31,179,621 (89.3%) | **£31,179,621 (29.9%)** |
| 2 | European Commission | £23,545,255 (34.0%) | £0 (0.0%) | **£23,545,255 (22.6%)** |
| 3 | Wellcome Trust | £14,144,606 (20.4%) | £1,217,835 (3.5%) | **£15,362,440 (14.7%)** |
| 4 | Coalition for Epidemic Preparedness and Innovation (CEPI) | £12,098,260 (17.5%) | £272,286 (0.8%) | **£12,370,546 (11.9%)** |
| 5 | Medical Research Council | £3,080,837 (4.4%) | £2,174,848 (6.2%) | **£5,255,685 (5.0%)** |
| 6 | Foundation for National Institute of Health (U.S.) | £5,729,292 (8.3%) | £0 (0.0%) | **£5,729,292 (5.5%)** |
| 7 | Innovate UK | £2,403,678 (3.5%) | £0 (0.0%) | **£2,403,678 (2.3%)** |
| 8 | European & Developing Countries Clinical Trials Partnership | £2,209,747 (3.2%) | £0 (0%) | **£2,209,747 (2.1%)** |
| 9 | Bill and Melinda Gates Foundation | £1,595,006 (2.3%) | £0 (0.0%) | **£1,595,006 (1.5%)** |
| 10-20 | Other | £4,506,697 (6.5%) | £68,106 (0.2%) | **£4,574,803 (4.4%)** |
| **Total** |  | **£69,313,379** | **£34,912,696** | **£104,226,076** |

## Discussion

Research conducted at the Jenner Institute of the University of Oxford provided the ChAdOx platform on which the Oxford-AstraZeneca vaccine is built. Our study identified that public funding accounted for 97.1-99.0% of the funding towards the R&D of the ChAdOx technology and its application for SARS-CoV-2. These include government and charitable funders, and the PPPs CEPI, PATH malaria vaccine initiative and CGIAR. Our study identified £104,226,076 of R&D funding reported in FOIs to the University of Oxford and £228,466,771 from the 21.0% of mentions with a matched grant amount in the scoping review for the ChAdOx technology and the Oxford-AstraZeneca vaccine.

Due to insufficient identifiable information that could link the two datasets, we were not able to cross-match the funding reported in academic articles and the FOIs, which is a major limitation of our study. Furthermore, exact grant amounts were retrievable from publicly available information for only 21.0% of grants mentioned in academic publications on ChAdOx. Receiving funding information through FOIs was largely successful, making it a useful method for reconstructing funders of R&D at public research institutions. The restriction we faced was the maximum amount of 18 hours a public institution in the U.K. is legally required to spend collecting the data, limiting the scope of these requests. It is therefore likely that we missed further public funding received by other departments of the University of Oxford working on the clinical trials and manufacturing of the vaccine. For example, grants to research groups working on the manufacturing of ChAdOx, such as the one led by Dr. Alexander Douglas at the Nuffield Department of Medicine[19], were not included in the FOI. However, some of these grants are captured within the funding acknowledgement statements in peer-reviewed articles, which did cover other research groups at the University of Oxford as well. By applying a methodology that included data collection through two different mechanisms, this should have captured most of the R&D costs of the fundamental research into the ChAdOx technology conducted at the Jenner Institute. Finally, it was not possible to measure relevant non-monetary contributions to the ChAdOx R&D, such as the participation in clinical trials, most recently in South Africa and Brazil for the Oxford-AstraZeneca vaccine[20]. There is also circa £18m worth of funding in the FOI regarding S.G and A.H that may be linked to the development of the vaccine, consisting of fellowship grants and general vaccine grants with descriptions too vague to attribute them to the development of ChAdOx specifically (listed in full in Supplementary File 4).

Beyond the funding captured in our study, the University of Oxford received at least £65.5 million from the U.K. Department of Business, Energy and Industrial Strategy for the development of the COVID-19 vaccine and the relevant clinical trials[21]. The UKRI database further listed two UKRI grants to the University of Oxford, worth £657,388[22]. These were not identified in the scoping review or FOI as they were awarded for the scale-up of manufacturing process of the Oxford-AstraZeneca vaccine. Additionally, the U.S. government awarded US$125.6 million and over US$1.2 billion in funding to AstraZeneca for vaccine trials, manufacturing and distribution of vaccine doses to the U.S. government[23, 24]. A further 9 donations totalling £1.8-2.9 million (included in Supplementary File 4) were reported by the University of Oxford in their response to our FOI, 2 of which came from charitable sources, totalling £50,000-100,000. The remaining 7 donations were private or anonymous funders. All 9 donations were not integrated into the FOI dataset as exact amounts were not provided and donor names or amounts were missing for 44.4% of donations.

The lack of transparency around the costs of R&D of novel health technologies is a prevailing issue, with large disparities in estimates reported[25]. Although there have been improvements in funding reporting in the past years, there are still major obstacles to investigating the funding of biomedical innovation based on disclosures made in the published scientific literature[26, 27]. Furthermore, the cumulative nature of scientific research makes it difficult to ascertain the R&D costs of previous innovation which may have enabled the development of the ChAdOx technology and the Oxford-AstraZeneca vaccine[28]. Of the grant mentions relevant to the R&D of ChAdOx identified through the scoping review, 79.0% could not be matched to an amount using searchable online grant databases. Attempting to match grants without award numbers was unreliable and inconsistent. Another issue was a lack of publicly available grant information, especially from the two main research institution funding bodies that contributed to the ChAdOx technology based on the funding acknowledgement statements, the Jenner Institute and The Oxford Martin School. Funding amounts from the private sector and PPPs were especially difficult to identify in this study as they often do not disclose their grants in publicly accessible databases. Initiatives to address the lack of transparency in R&D funding have been initiated, such as a World Health Assembly resolution[29]. However, the voluntary nature of such initiatives and opposition from the private sector and high-income governments limit efforts to increase R&D transparency[30].

Despite a lack of research funding transparency, our findings show the dominance of government and charity funding throughout the R&D process of the ChAdOx technology, which accelerated during the pandemic. Public funding has been especially critical for vaccine research, where the failure rate is as high as 94%, and has enabled the rapid development of many COVID-19 vaccines[13, 31]. Prior to the pandemic, the WHO identified emerging infectious diseases requiring urgent R&D efforts[32]. Their Blueprint for Action to Prevent Epidemics listed diseases on which the ChAdOx technology has been studied, including Nipah, MERS, and Ebola[8, 33, 34]. In addition to government and charitable funders, PPPs are growing global health actors prominent in R&D efforts for diseases endemic to lower-income populations, for which a funding gap prevails[35, 36]. Launched in 2017 as an innovative partnership between public, private, philanthropic, and civil organisations, CEPI’s mission is accelerating vaccine development to address pathogens of pandemic potential, whereas the PATH malaria vaccine initiative was created to develop malaria vaccines[37]. Since the PPPs that contributed to ChAdOx were largely supported by public funding, we categorised them as public in our study[38–40]. To recognise the public contributions and risk-taking in the R&D of the ChAdOx technology on which the Oxford-AstraZeneca vaccine relies, the benefits of this research should be shared fairly and equitably with the global population[41].

In response to the pandemic, Oxford University Innovation, a subsidiary of the University of Oxford managing the university’s technology transfer, published a statement outlining the university’s default approach for the licensing of COVID-19 related intellectual property to third parties[42]. In this statement, the university committed to non-exclusive, royalty-free licensing and price-setting at-cost or cost plus small a profit margin for the duration of the pandemic. Despite these commitments, the university chose to enter an exclusive licensing agreement with the British-Swedish pharmaceutical company AstraZeneca for the COVID-19 vaccine[43]. As of March 2021, the University of Oxford has not made the licensing agreement publicly available, however, the university has expressed the intention to publish a redacted version[44]. While AstraZeneca pledged to sell the vaccine globally at no profit during the pandemic, the price of the vaccine reportedly includes a profit margin of 20% on top of the production cost[45, 46]. The Oxford-AstraZeneca vaccine is offered at the lowest price of $5 per course, making it one of the most affordable vaccines available for COVID-19[4]. Vaccine prices paid by countries are kept confidential, yet discrepancies in pricing have been reported with lower income countries seemingly paying more than higher income countries[47]. AstraZeneca has, in collaboration with the Serum Institute of India, committed a large number of vaccine doses to the COVAX facility[48]. However, the Oxford-AstraZeneca vaccine is facing global supply issues following manufacturing delays and export disputes[49, 50]. Global equitable access is further hindered by bilateral purchasing agreements made between AstraZeneca and countries outside of COVAX[51]. Given that the Oxford-AstraZeneca vaccine price is determined by the pandemic status and SARS-CoV-2 will likely become an endemic virus requiring repeated vaccinations, affordability of the vaccine post-pandemic remains a concern[52].

While the Oxford-AstraZeneca vaccine has been licensed exclusively to AstraZeneca, the type and any conditions included in the licenses of the ChAdOx vaccine platform patents to the University of Oxford’s spin out company, Vaccitech, remain unknown[53]. As the ChAdOx vaccine platform is potentially applicable to many more global health challenges, including emerging infectious diseases and pathogens of pandemic potential other than SARS-CoV-2, its mode of technology transfer is of global public health relevance with potential impact for global equitable access and affordability.

## Conclusion

Approximating the funding of ChAdOx offers a relevant and timely case study to understand wider trends in R&D taking place at universities and the importance of transparency in funding reporting. We found that government and charitable funders provided the majority of funding to the University of Oxford towards the R&D of the Oxford-AstraZeneca vaccine and the underlying ChAdOx technology, which may have significant implications for the global discourse around vaccine nationalism and COVID-19 health technology access. Understanding who contributed to the development of ChAdOx is of importance to other global health challenges as well, considering that the vaccine platform may be used for multiple applications beyond SARS-CoV-2, offering an opportunity to rapidly and equitably develop affordable solutions to other existing and emerging infectious disease threats. However, a lack of transparency of funding reporting mechanisms hinders the discourse surrounding public and private contributions towards R&D and the cost of R&D. We therefore urge medical journal editors and research funders to further improve their funding reporting mechanisms by publishing funding and grant information more widely in a publicly accessible manner.

## Data Availability

Supplementary Information is available for this paper. Any correspondence and requests for materials should be addressed to Ms. Sarai Keestra, Amsterdam UMC, Meibergdreef 9, 1105 AZ Amsterdam, The Netherlands, or by email at.

## Acknowledgements

The authors wish to thank Molly Pugh-Jones for filing one of the FOI requests to the University of Oxford and Manuel Martin for his comments on an earlier version of the manuscript.

## Authors’ contributions

S.K. and S.W. conceived of the study. F.R. and S.K. devised the methodology. F.R. made the search strategy. S.K. filed the FOI requests and managed communication with the University of Oxford. F.R. and S.K. screened all articles. S.C. and H.R. extracted the data from the articles. Y.R. searched grant databases using award numbers. S.C. and Y.R. classified all the grants from the FOI. T.P. contributed to the data management. S.C. and S.K. wrote up the methods and results. S.W., S.C., S.K., R.O., A.E. and R.B. wrote the first draft of the manuscript. All authors contributed to and edited the final manuscript.

---

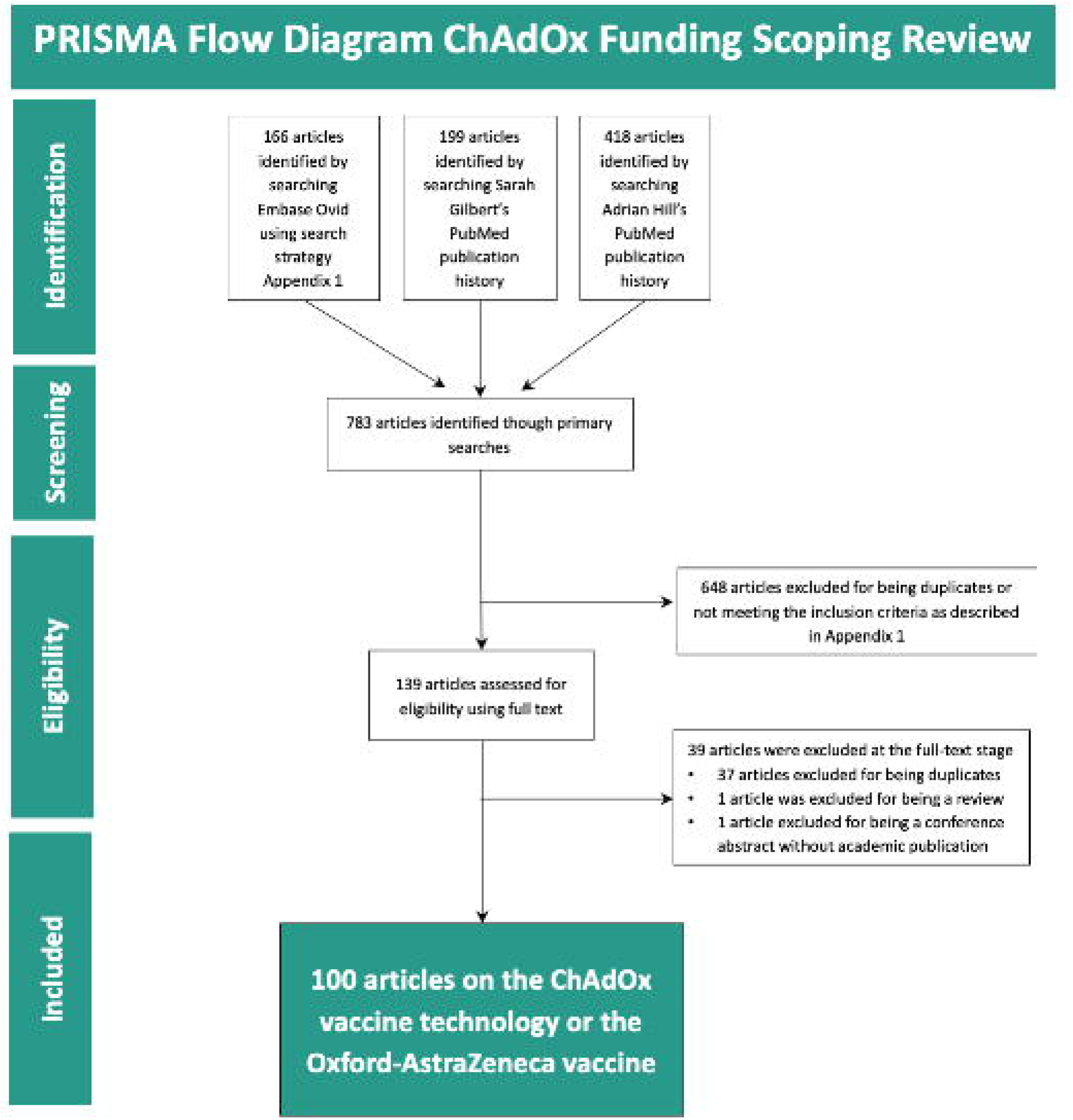

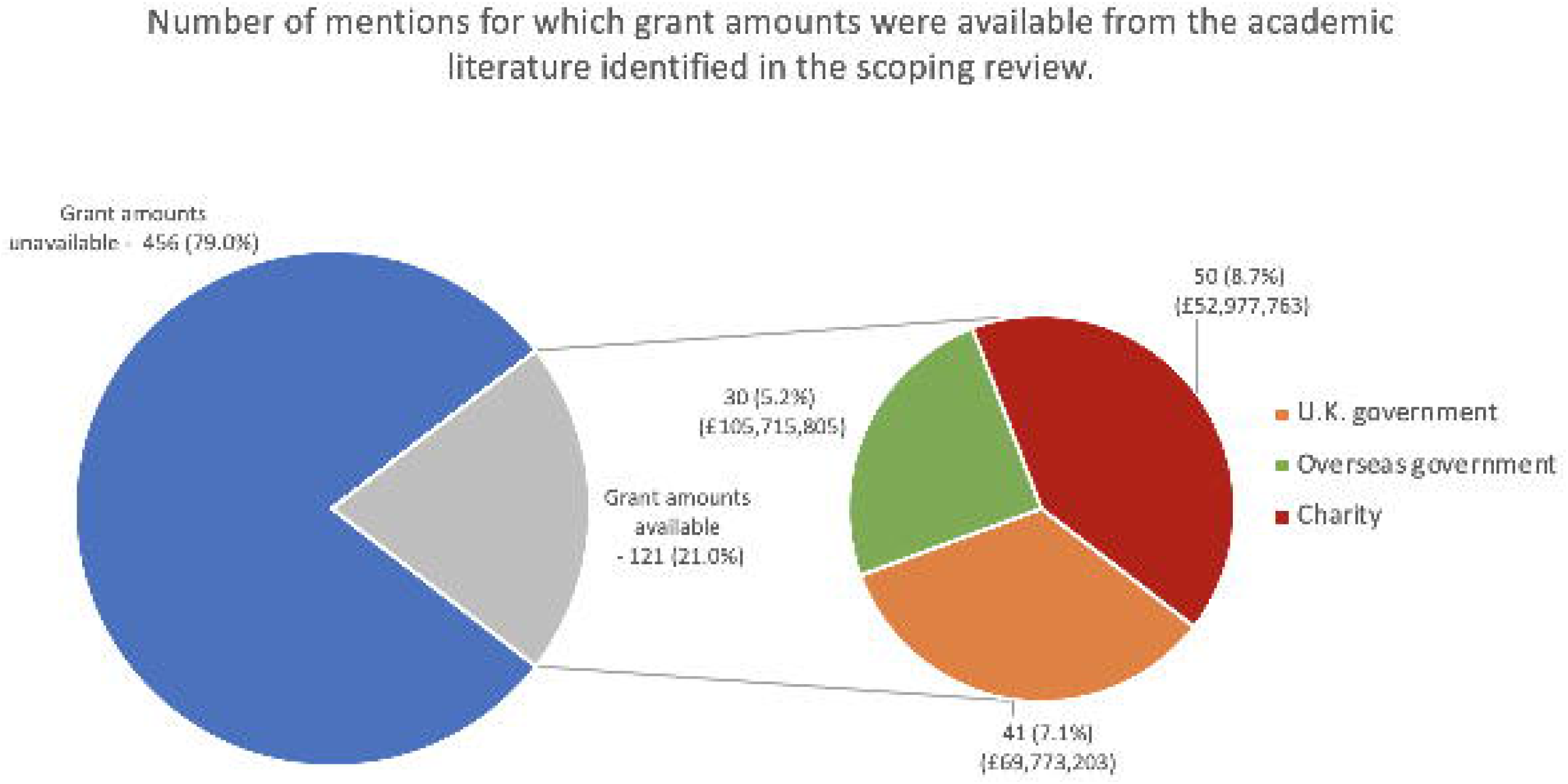

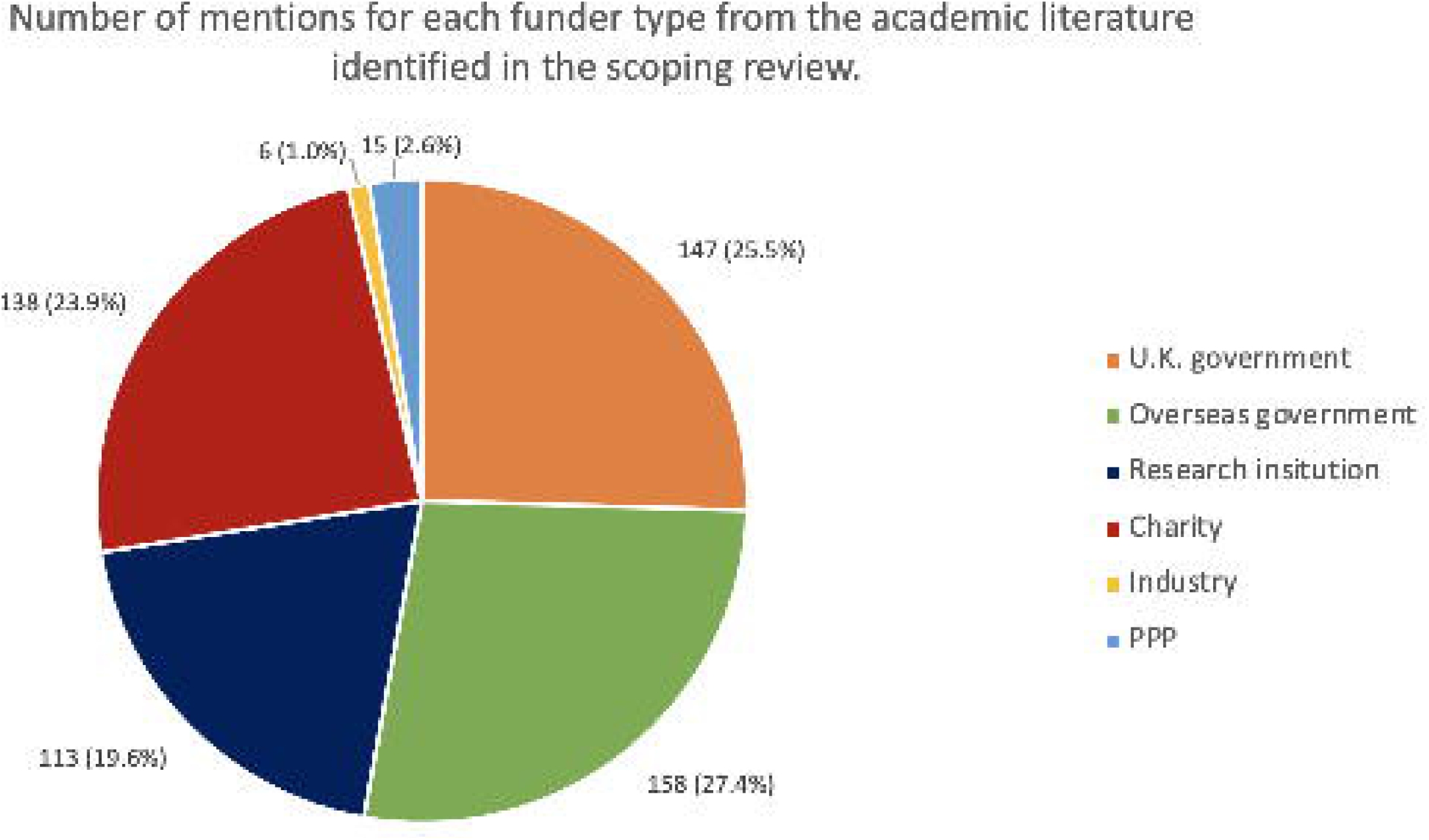

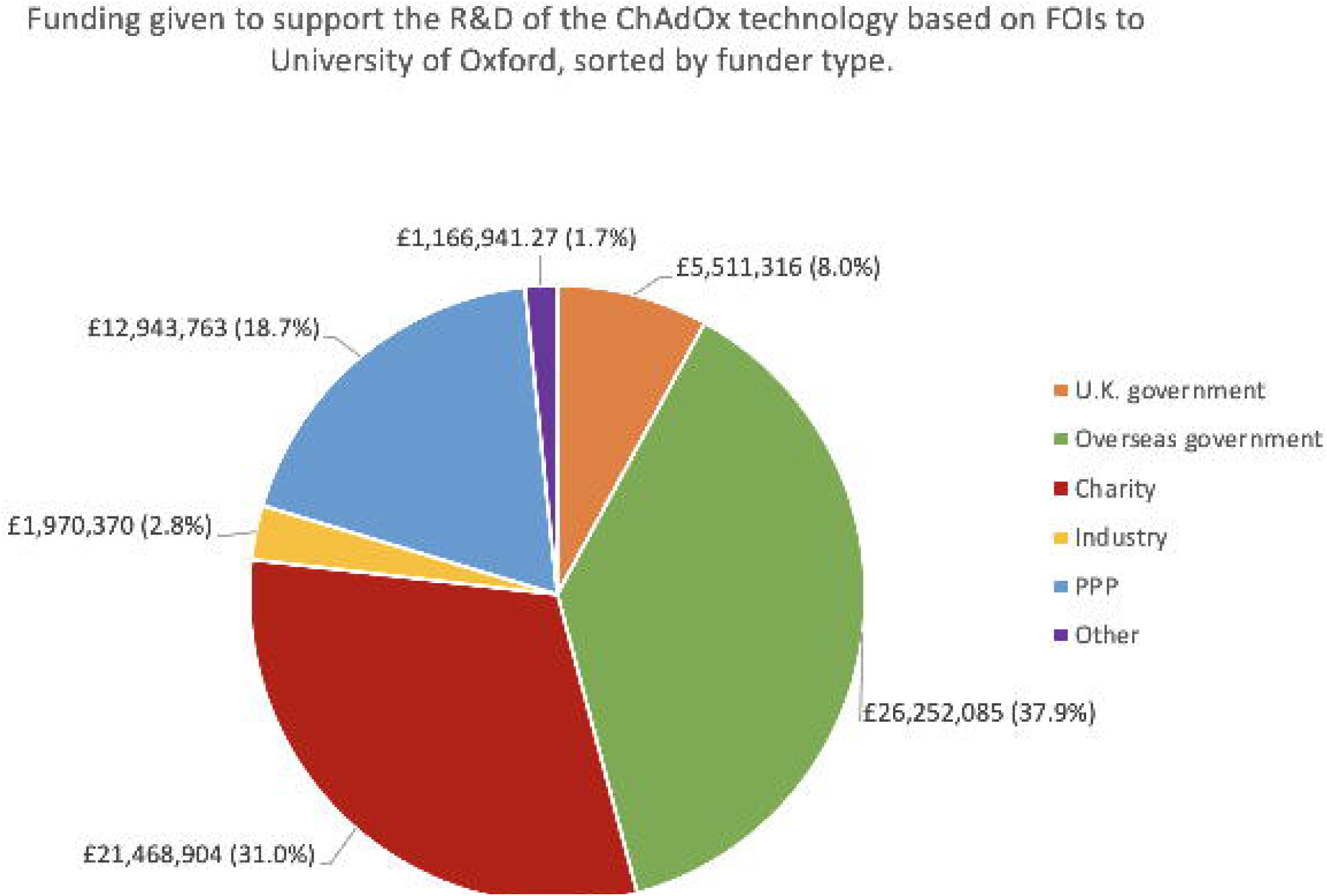

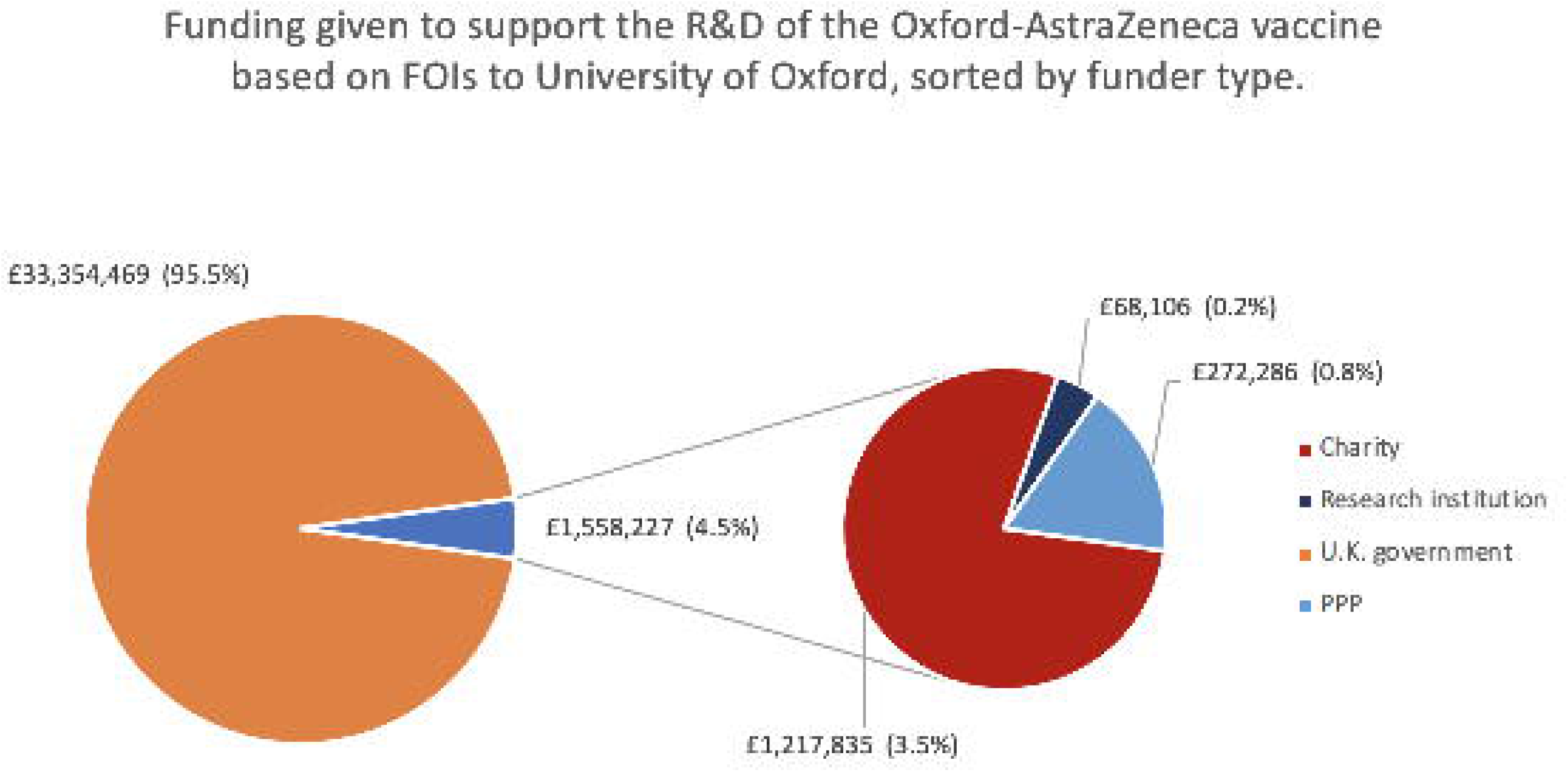

## References

1 Medicines & Healthcare products Regulatory Agency. Conditions of Authorisation for COVID-19 Vaccine AstraZeneca. 2021.https://www.gov.uk/government/publications/regulatory-approval-of-covid-19-vaccine-astrazeneca/conditions-of-authorisation-for-covid-19-vaccine-astrazeneca (accessed 29 Mar 2021).

2 Mallapaty S, Callaway E. What scientists do and don’t know about the Oxford-AstraZeneca COVID vaccine. Nature 2021;592:15–7.

3 Ramasamy MN, Minassian AM, Ewer KJ, et al. Safety and immunogenicity of ChAdOx1 nCoV-19 vaccine administered in a prime-boost regimen in young and old adults (COV002): a single-blind, randomised, controlled, phase 2/3 trial. Lancet 2020;396:1979–93.

4 Wouters OJ, Shadlen KC, Salcher-Konrad M, et al. Challenges in ensuring global access to COVID-19 vaccines: production, affordability, allocation, and deployment. Lancet 2021;397:1023–34.

5 University of Oxford. Oxford University breakthrough on global COVID-19 vaccine. 2020.https://www.ox.ac.uk/news/2020-11-23-oxford-university-breakthrough-global-covid-19-vaccine (accessed 6 Apr 2021).

6 Barnes E, Folgori A, Capone S, et al. Novel adenovirus-based vaccines induce broad and sustained T cell responses to HCV in man. Sci Transl Med 2012;4:115ra1.

7 Afolabi MO, Tiono AB, Adetifa UJ, et al. Safety and Immunogenicity of ChAd63 and MVA ME-TRAP in West African Children and Infants. Mol Ther 2016;24:1470–7.

8 van Doremalen N, Haddock E, Feldmann F, et al. A single dose of ChAdOx1 MERS provides protective immunity in rhesus macaques. Sci Adv 2020;6:eaba8399.

9 Folegatti PM, Bittaye M, Flaxman A, et al. Safety and immunogenicity of a candidate Middle East respiratory syndrome coronavirus viral-vectored vaccine: a dose-escalation, open-label, non-randomised, uncontrolled, phase 1 trial. Lancet Infect Dis 2020;20:816–26.

10 Cleary EG, Beierlein JM, Khanuja NS, et al. Contribution of NIH funding to new drug approvals 2010–2016. Proc Natl Acad Sci U S A 2018;115:2329–34.

11 Mazzucato M. Putting the Public Back in Public Health. Project Syndicate. 2018;:Online.

12 National Vaccine Advisory Committee. United States vaccine research: a delicate fabric of public and private collaboration. National Vaccine Advisory Committee. Pediatrics 1997;100:1015–20.

13 Kiszewski AE, Cleary EG, Jackson MJ, et al. NIH funding for vaccine readiness before the COVID-19 pandemic. Vaccine Published Online First: 8 March 2021. doi:10.1016/j.vaccine.2021.03.022

14 Ouzzani M, Hammady H, Fedorowicz Z, et al. Rayyan—a web and mobile app for systematic reviews. Syst Rev 2016;5:210.

15 Currency converter calculator. https://www.xe.com/currencyconverter/ (accessed 6 Apr 2021).

16 Universities Allied for Essential Medicines (UAEM), Student National Medical Association, American Medical Student Association. Tracking COVID-19 public investment in global COVID-19 research and development. 2020.https://www.publicmeds4covid.org/ (accessed 13 Mar 2021).

17 WhatDoTheyKnow. FOI/20201130/03. Freedom of Information Request to University of Oxford. 2020.https://www.whatdotheyknow.com/request/breakdown_of_funding_for_the_cha (accessed 6 Apr 2021).

18 WhatDoTheyKnow. FOI/20210205/02. Freedom of Information Request to University of Oxford. 2021.https://www.whatdotheyknow.com/request/funding_for_oxford_astrazeneca_v (accessed 6 Apr 2021).

19 University of Oxford, Nuffield Department of Medicine. Alexander (Sandy) Douglas. https://www.ndm.ox.ac.uk/team/alexander-sandy-douglas (accessed 6 Apr 2021).

20 The University of the Witwatersrand, Johannesburg. Oxford Covid-19 Vaccine Trial. https://www.wits.ac.za/covid19vaccine/oxford-covid-19-vaccine-trial/ (accessed 6 Apr 2021).

21 University of Oxford. Funding and manufacturing boost for UK vaccine programme. 2020.https://www.ox.ac.uk/news/2020-05-18-funding-and-manufacturing-boost-uk-vaccine-programme (accessed 6 Apr 2021).

22 UK Research and Innovation. Find COVID-19 research and innovation supported by UKRI. https://www.ukri.org/find-covid-19-research-and-innovation-supported-by-ukri/ (accessed 6 Apr 2021).

23 GovTribe. Definitive Contract 75A50120C00114. Federal Contract Award. 2020.https://govtribe.com/award/federal-contract-award/definitive-contract-75a50120c00114 (accessed 6 Apr 2021).

24 GovTribe. Other Transaction IDV W15QKN2191003. Federal Contract IDV Award. 2020.https://govtribe.com/award/federal-idv-award/other-transaction-idv-w15qkn2191003 (accessed 6 Apr 2021).

25 The United Nations Secretary-General’s High-Level Panel on Access to Medicines. The United Nations Secretary-General’s High-Level Panel on Access to Medicines Report - Promoting Innovation and Access to Health Technologies. 2016. https://static1.squarespace.com/static/562094dee4b0d00c1a3ef761/t/57d9c6ebf5e231b2f02cd3d4/1473890031320/UNSG+HLP+Report+FINAL+12+Sept+2016.pdf

26 Iqbal SA, Wallach JD, Khoury MJ, et al. Reproducible Research Practices and Transparency across the Biomedical Literature. PLOS Biology. 2016;14:e1002333. doi:10.1371/journal.pbio.1002333

27 Wallach JD, Boyack KW, Ioannidis JPA. Reproducible research practices, transparency, and open access data in the biomedical literature, 2015–2017. PLoS Biol 2018;16:e2006930.

28 Wimmer S, Keestra SM. Public Risk-Taking and Rewards During the COVID-19 Pandemic - A Case Study of Remdesivir in the Context of Global Health Equity. Int J Health Policy Manag Published Online First: 6 September 2020. doi:10.34172/ijhpm.2020.166

29 World Health Assembly, 72. Improving the transparency of markets for medicines, vaccines, and other health products. World Health Organization. 2020.https://apps.who.int/iris/handle/10665/329301 (accessed 6 Apr 2021).

30 Silverman E, editor. Pharma pushes back against setting international standards for drug-pricing transparency. Stat 2019. https://www.statnews.com/pharmalot/2019/05/08/pharma-transparency-resolution-drug-prices/

31 Pronker ES, Weenen TC, Commandeur H, et al. Risk in vaccine research and development quantified. PLoS One 2013;8:e57755.

32 Mehand MS, Al-Shorbaji F, Millett P, et al. The WHO R&D Blueprint: 2018 review of emerging infectious diseases requiring urgent research and development efforts. Antiviral Res 2018;159:63–7.

33 van Doremalen N, Lambe T, Sebastian S, et al. A single-dose ChAdOx1-vectored vaccine provides complete protection against Nipah Bangladesh and Malaysia in Syrian golden hamsters. PLoS Negl Trop Dis 2019;13:e0007462.

34 Ewer K, Rampling T, Venkatraman N, et al. A Monovalent Chimpanzee Adenovirus Ebola Vaccine Boosted with MVA. N Engl J Med 2016;374:1635–46.

35 Wheeler C, Berkley S. Initial lessons from public-private partnerships in drug and vaccine development. Bull World Health Organ 2001;79:728–34.

36 Young R, Bekele T, Gunn A, et al. Developing new health technologies for neglected diseases: a pipeline portfolio review and cost model. Gates Open Res 2018;2:23.

37 Coalition for Epidemic Preparedness Innovations. Our Mission. https://cepi.net/about/whyweexist/ (accessed 6 Apr 2021).

38 Coalition for Epidemic Preparedness Innovations. Board of Directors’ Report and Annual Accounts 2020. 2020. https://cepi.net/wp-content/uploads/2021/03/2020-Board-of-Directors-Report-and-Annual-A ccounts-incl-Auditors-report.pdf

39 PATH Malaria Vaccine Initiative. Donors. https://www.malariavaccine.org/about-us/donors (accessed 6 Apr 2021).

40 CGIAR. Funder Analysis. https://www.cgiar.org/food-security-impact/finance-reports/dashboard/funder-analysis/ (accessed 6 Apr 2021).

41 Nuffield Council on Bioethics. Fair and equitable access to COVID-19 treatments and vaccines. 2020. https://www.nuffieldbioethics.org/publications/fair-and-equitable-access-to-covid-19-treatments-and-vaccines/read-the-briefing-note/factors-affecting-fair-and-equitable-access

42 Oxford University Innovation, University of Oxford. Expedited access for COVID-19 related IP. Oxford University Innovation. https://innovation.ox.ac.uk/technologies-available/technology-licensing/expedited-access-covid-19-related-ip/ (accessed 13 Mar 2021).

43 Statement of Sir Menelas Pangalos, Ph. D. Executive Vice President Biopharmaceutical Research & Development AstraZeneca Before the Subcommittee on Oversight and Investigations Committee on Energy and Commerce U.S. House of Representatives. Pathway to a Vaccine: Efforts to Develop a Safe, Effective and Accessible COVID-19 Vaccine. 2020. https://docs.house.gov/meetings/IF/IF02/20200721/110926/HHRG-116-IF02-Wstate-PangalosM-20200721.pdf

44 WhatDoTheyKnow. FOI/20200603/06. Freedom of Information Request to University of Oxford. 2020.https://www.whatdotheyknow.com/request/agreements_between_oxford_univer#incoming-1755923 (accessed 6 Apr 2021).

45 AstraZeneca. AstraZeneca to supply Europe with up to 400 million doses of Oxford University’s vaccine at no profit. 2020.https://www.astrazeneca.com/media-centre/press-releases/2020/astrazeneca-to-supply-europe-with-up-to-400-million-doses-of-oxford-universitys-vaccine-at-no-profit.html (accessed 6 Apr 2021).

46 Mancini DP, Cookson C, editors. Vaccine deal allows AstraZeneca to take up to 20% on top of costs. Financial Times 2020. https://www.ft.com/content/e359159b-105c-407e-b1be-0c7a1ddb654b

47 Paun C, Furlong A, editors. Poorer countries hit with higher price tag for Oxford/AstraZeneca vaccine. Politico 2021. https://www.politico.eu/article/astrazeneca-vaccine-cost-higher-in-poorer-countries-coronavirus/

48 AstraZeneca. AstraZeneca advances mass global rollout of COVID-19 vaccine through COVAX. 2021.https://www.astrazeneca.com/media-centre/press-releases/2021/astrazeneca-advances-mass-global-rollout-of-covid-19-vaccine-through-covax.html (accessed 6 Apr 2021).

49 Menon S, editor. India coronavirus: Why have vaccine exports been suspended? BBC 2021. https://www.bbc.co.uk/news/world-asia-india-55571793

50 Herszenhorn DM, Von Der Burchard H, editors. EU moves toward six-week vaccine export cut. Politico 2021. https://www.politico.eu/article/commission-proposes-six-week-vaccine-export-ban-amid-fears-of-trade-war/

51 Callaway E. The unequal scramble for coronavirus vaccines — by the numbers. Nature. 2020;584:506–7. doi:10.1038/d41586-020-02450-x

52 Phillips N. The coronavirus is here to stay-here’s what that means. Nature 2021;590:382–4.

53 Vaccitech Limited. Vaccitech Technology. https://www.vaccitech.co.uk/technology/ (accessed 6 Apr 2021).

