## Supplementary File 1 for "Who funded the research behind the Oxford-AstraZeneca COVID-19 vaccine? Approximating the funding to the University of Oxford for the research and development of the ChAdOx vaccine technology"

**Appendix 1 – Search Strategy**

Embase Ovid (1974 to 26th October 2020) searched 26th October 2020

| No. | Search Terms | Results |
|---|---|---|
| 1 | Coronavir$ or "corona virus" or $coronavirus or covid19 or "covid 19" or nCoV or "CoV 2" or CoV2 or sarscov2 or 2019nCoV or "novel CoV" or "wuhan virus" or ((wuhan or hubei or huanan) and ("severe acute respiratory" or pneumonia) and outbreak) | |
| 2 | ChAdOx1 or ChAdOx2 or Chimpanzee adenovirus-vectored or AZD1222 or MN908947 or spike protein | |
| 3 | 1 OR 2 | |
| 4 | Vaccitech or AstraZeneca or AZC or Oxford or Jenner or JI | |
| 5 | 3 AND 4 | 166 |

Pubmed (1946 to 29th November 2020) searched on 29th November 2020

| No. | Search Terms | Results |
|---|---|---|
| 1 | Adrian Hill [Author] | 418 |

Pubmed (1946 to 29th November 2020) searched on 29th November 2020

| No. | Search Terms | Results |
|---|---|---|
| 1 | Sarah Gilbert [Author] | 199 |

**Figure 1 - PRISMA Flow Diagram scoping review of the academic literature on the ChAdOx technology**

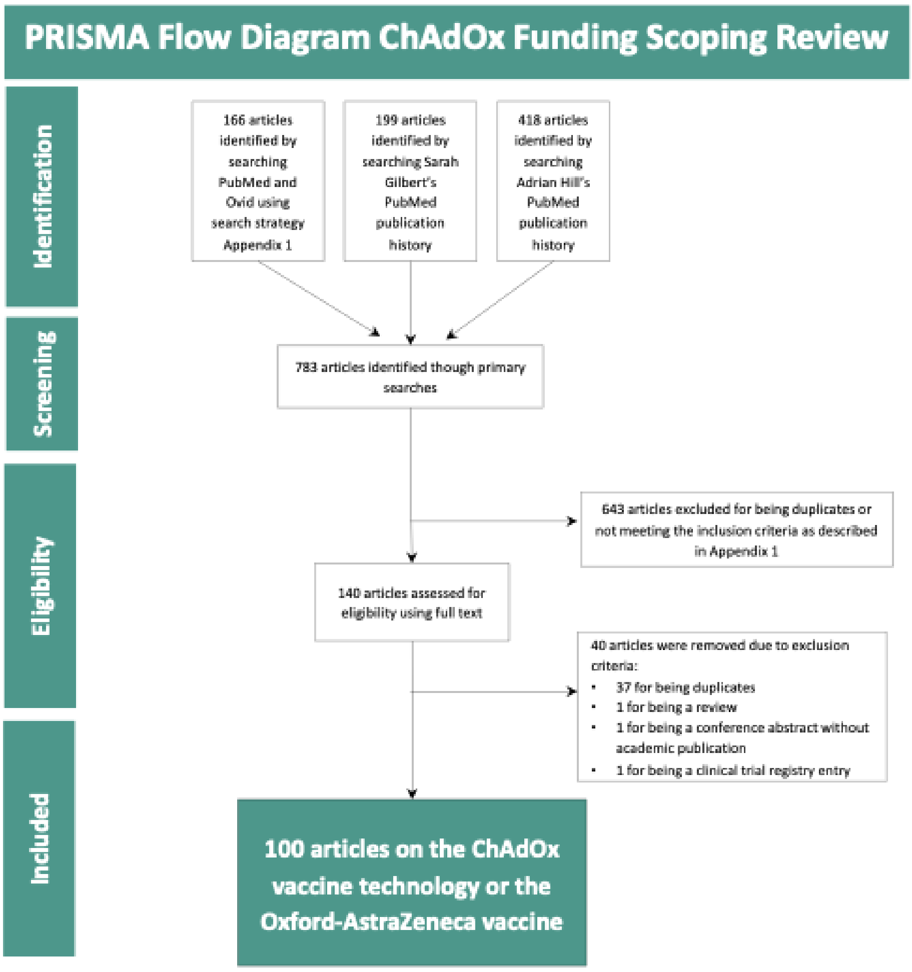
PRISMA Flow Diagram ChAdOx Funding Scoping Review
Identification
Screening
Eligibility
Included
166 articles identified by searching PubMed and Ovid using search strategy Appendix 1
199 articles identified by searching Sarah Gilbert's PubMed publication history
418 articles identified by searching Adrian Hill's PubMed publication history
783 articles identified though primary searches
643 articles excluded for being duplicates or not meeting the inclusion criteria as described in Appendix 1
140 articles assessed for eligibility using full text
40 articles were removed due to exclusion criteria:
37 for being duplicates
1 for being a review
1 for being a conference abstract without academic publication
1 for being a clinical trial registry entry
100 articles on the ChAdOx vaccine technology or the Oxford-AstraZeneca vaccine
