## Supplementary File 2 for "Who funded the research behind the Oxford-AstraZeneca COVID-19 vaccine? Approximating the funding to the University of Oxford for the research and development of the ChAdOx vaccine technology"

| Authors | Publication title | Date | Journal | Funding acknowledgement | Competing Interests | If relevant: clinical trial ID | Comments | Date of data extraction |
|---|---|---|---|---|---|---|---|---|
| Afolabi MO | Safety and Immunogenicity of ChAd63 and MVA ME-TRAP in West African Children and Infants | 2016 | Official journal of the American Society of Gene & Cell Therapy | Trials Partnership (EDCTP) and was performed by the Malaria Vectored Vaccines Consortium (MVVC), a four and a half year integrated project funded by the European and Developing Countries Clinical Trials Partnership (EDCTP, grant number IP.2008.31100.001). The work was also supported by the UK National Institute of Health Research (NIHR) through the NIHR Oxford Biomedical Research Centre (http://www.oxfordbrc.org/) [A91301 Adult Vaccine], The Wellcome Trust (http://www.wellcome.ac.uk/) (084113/Z/07/Z) and the Medical Research Council. Cofunding was also provided by the Swedish International Development Cooperation Agency (Sida) and Irish Aid. This research was supported by the UK Medical Research Council (MRC) and the UK Department for International Development (DFID) under the MRC/DFID Concordat agreement and MC_UP_A900/1122 | AVSH is a named inventor on patent applications on malaria vectored vaccines and immunization regimens. Authors from ReiThera are employees of and/or shareholders in ReiThera, which is developing vectored vaccines for malaria and other diseases. | NCT01373879, NCT01450293, NCT01635647. PACTR201204000362870, PACTR201401000363170, PACTR201208000404131 | N/A | 08/12/2020 |
| Alcock R | Long-term thermostabilization of live poxviral and adenoviral vaccine vectors at supraphysiological temperatures in carbohydrate glass | 2010 | Elselvier - Vaccine | No funding disclosure found | Conflict of interest: SCG is a co-founder of, consultant to and shareholder in Vac-citech plc which is developing vectored influenza and MERSvaccines. | N/A | No funding disclosure found | 16/12/20 |
| Alharbi NK | ChAdOx1 and MVA based vaccine candidates against MERS-CoV elicit neutralising antibodies and cellular immune responses in mice | 2017 | Vaccine | No funding statement found | SCG is a co-founder of, consultant to and shareholder in Vaccitech plc which is developing vectored influenza and MERS vaccines. | N/A | N/A | 21/12/2020 |
| Alharbi NK | Humoral Immunogenicity and Efficacy of a Single Dose of ChAdOx1 MERS Vaccine Candidate in Dromedary Camels | 2019 | Nature | This study is funded by KAIMRC, project RC16/093 granted to the PI: Naif Khalaf Alharbi; in addition, animals, research farm, and animal logistics were financially supported by MEWA, Saudi Arabia. SCG is a Jenner Investigator and supported the manufacturing of the vaccine batch. | SCG is a co-founder of and consultant to Vaccitech, a spin-out company from the University of Oxford which has commercial rights to ChAdOx1 MERS. ChAdOx1 MERS vaccine is registered as an IP, number: WO 2018/215766. The remaining authors declare no potential conflict of interest. | N/A | N/A | 18/12/20 |
| Alves E | Evaluation of Plasmodium vivax Cell-Traversal Protein for Ookinetes and Sporozoites as a Preerythrocytic P. vivax Vaccine | 2017 | Clinical and Vaccine Immunology | The work was funded by a Wellcome Trust Career Development Fellowship award(grantnumber 097395/Z/11/Z) to A.R.-S. | N/A | N/A | N/A | 08/12/2020 |
| Antrobus RD | Clinical assessment of a novel recombinant simian adenovirus ChAdOx1 as a vectored vaccine expressing conserved Influenza A antigens | 2014 | Molecular Therapy | The study was funded by grants from the UK MRC, the NIHR through the Oxford Biomedical Research Centre, and the Oxford Martin school. | T.L is an Oxford Martin fellow. S.C.G and A.V.S.H. are Jenner Investigators. S.C.G, M.D.D, and A.V.S.H are named inventors on a pat-ent application describing the ChAdOx1 vector (GB Patent Application No. 1108879.6). | NCT01623518 | N/A | 08/12/2020 |
| Asthagiri Arunkumar G | Vaccination with viral vectors expressing NP, M1 and chimeric hemagglutinin induces broad protection against influenza virus challenge in mice | 2019 | Elselvier Vaccine | The study was fundedby an MRC Biomedical Catalyst DPFS_DCS award (MR/N006372/1).In addition, this study was partially funded by the NIAID Centers ofExcellence for Influenza Research and Surveillance contract(CEIRS, HHSN272201400008C). Andriani Ioannou was supportedby an NIAID T32 Virus-Host Interactions training grant(5T32AI007647-17). | The Icahn School of Medicine at Mount Sinai has filed patentapplications regarding influenza virus vaccines with FlorianKrammer being an inventor. Sarah Gilbert is an inventor on patentscovering ChAdOx1 and MVA-NP+M1, filed and owned by theUniversity of Oxford, and is a co-founder of and consultant toVaccitech, a University of Oxford spin-out company which isundertaking advanced clinical development of viral vectoredinfluenza vaccines. | N/A | N/A | 18/12/20 |
| Atcheson E | Tailoring a Plasmodium vivax Vaccine To Enhance Efficacy through a Combination of a CSP Virus-Like Particle and TRAP Viral Vectors | 2018 | Infection and Immunity | The work was funded by a Wellcome Trust Career Development Fellowship award(grant 097395/Z/11/Z) to A.R.-S., who is also a Jenner Investigator and an Oxford MartinFellow and is supported by MRC-DPFS (grant MR/N019008/1). A.M.S. was funded byEVIMalaR's program funding (FP7/2007-2013) under grant agreement number 242095. E.A.was funded by CAPES from Science without Border program. A.V.S.H. is supported by aWellcome Trust grant (number 095540/Z/11/Z) and is a Jenner Investigator and an OxfordMartin Fellow. | N/A | N/A | N/A | 10/12/2020 |
| Barnes E | Novel adenovirus-based vaccines induce broad and sustained T cell responses to HCV in man | 2012 | Science Transitional Medicine | European Union (Framework VI;HEPACIVAC); Medical Research Council (UK); Wellcome Trust; Oxford NIHR Biomedical ResearchCentre; James Martin School for 21st Century, Oxford; Wellcome Trust Clinical Research Facility,Birmingham; National Institute for Health and Research Liver Biomedical Research Unit, Birmingham;and NIH grant 1U19AI082630-01. | S. Colloca, A.F., R.C., and A.N.are named inventors on patent applications covering HCV-vectored vaccines and chimpanzee ad-enovirus vectors [WO 2006133911 (A3) hepatitis Cvirus nucleic acid vaccine, WO 2005071093 (A3)chimpanzee adenovirus vaccine carriers, WO 03031588 (A2) hepatitis Cvirus vaccine]. P.K. has actedas a consultant to Tibotec and Pfizer on antiviraltherapy. Authors from Okairos are employees ofand/or shareholders in Okairos. The other authors declare that they have no competing interests. | NCT01070407, 2007-004259-12 | N/A | 10/12/2020 |
| Barnes E | ChAdOx1-HBV therapeutic vaccine: Phase 1 study results in healthy volunteers and patients with chronic hepatitis B | 2020 | N/A - only on clinicaltrials.gov (https://clinicaltrials.gov/ct2/show/NCT04297917) | N/A | N/A | NCT04297917 | Study still recruiting | 18/12/20 |
| Bauza K | Efficacy of a Plasmodium vivax malaria vaccine using ChAd63 and modified vaccinia Ankara expressing thrombospondin-related anonymous protein as assessed with transgenic Plasmodium berghei parasites | 2014 | Infection and Immunity | The work was funded by a Wellcome Trust Career Development Fel-lowship award, grant number 097395, to A.R-S. A.R.-S. and A.V.S.H. areJenner Investigators and Oxford Martin School Fellows. E.Y.J. and T.M.are funded by the Medical Research Council and Cancer Research UnitedKingdom. Work at the Wellcome Trust Sanger Institute was funded byWellcome Trust grant number WT098051. | N/A | N/A | N/A | 10/12/2020 |
| Betts G | Optimising immunogenicity with viral vectors: mixing MVA and HAdV-5 expressing the mycobacterial antigen Ag85A in a single injection | 2012 | Plos One | Funding was provided by NEWTBVAC (EC FP7). HM is a Wellcome Trust Senior Research Fellow (www.wellcome.ac.uk; WT076943MA). HM, AH and AR-Sare Jenner Institute Investigators. AR-S is a Wellcome Trust Career Development Fellow (097395). | The authors have read the journal's policy and have the following conflicts: AVSH, AAP, and HM are named inventors in a patent filingrelated to MVA85A and are shareholders in a joint venture, OETC, formed for the future development of this vaccine. AVSH and HM are named as co-inventorsonpatents related to heterologous prime-boost immunization. There are no other conflicts of interest. These conflicts of interest will not in any way interfere withthe authors' adherence to the journal's policies on sharing data and materials. | N/A | N/A | 10/12/2020 |
| Biswas S | Assessment of humoral immune responses to blood-stage malaria antigens following ChAd63-MVA immunization, controlled human malaria infection and natural exposure | 2014 | Plos One | This work was supported by the UK Medical Research Council (MRC; http://www.mrc.ac.uk) [grant number G0700735]; the EMVDA (European MalariaVaccine Development Association; http://www.emvda.org), a European Commission FP6-funded consortium [LSHP-CT-2007-037506]; the UK National Institute ofHealth Research through the Oxford Biomedical Research Centre (http://[084113/Z/07/Z]; and by EVIMalaR (http://www.evimalar.org) funded by the European Community'sSeventh Framework Programme (FP7/2007-2013)[Grant agreement No. 242095]. The GIA work was supported by the PATH Malaria Vaccine Initiative (http://www.malariavaccine.org) and the Intramural Program of the National Institutes of Health, National Institute of Allergy and Infectious Diseases (http://www.niaid.nih.gov). SHH holds a Wellcome Trust Research Training Fellowship (097940/Z/11/Z). AVSH and SJD are Jenner Investigators (http://www.jenner.ac.uk). SB is a NDM Leadership Fellow (http://www.ndm.ox.ac.uk) and Junior Research Fellow of St Catherine's College, Oxford University (http://www.stcatz.ox.ac.uk). SJD is a UK MRC Career Development Fellow [G1000527] and Lister Institute Research Prize Fellow (http://www.lister-institute.org.uk). The funders had no role in study design, data collection and analysis, decision to | SCdC, KAC, AVSH and SJD are named inventors on patent applications covering malaria vaccines and immunization regimes (Adenoviralvectors encoding a pathogen or tumour antigen, WO/2008/122811; Viral vector immunogenic compositions, GB1016471.3). This does not alter the authors'adherence to PLoS ONE policies on sharing data and materials. | NCT01095055, NCT01003314, NCT01142765, NCT00890760 | N/A | 10/12/2020 |
| Biswas S | Transgene optimization, immunogenicity and in vitro efficacy of viral vectored vaccines expressing two alleles of Plasmodium falciparum AMA1 | 2011 | Plos One | SB was funded by MalParTraining, an FP6-funded Marie Curie Action under contract number MEST-CT-2005-020492. The GIA work was supported inpart by the PATH-MVI Malaria Vaccine Initiative (MVI) and the Intramural Program of the National Institutes of Health, National Institute of Allergy and Infectious Diseases and in part by the EMVDA (European Malaria Vaccine Development Association, a European Commission FP6-funded consortium). AAH is funded bytheUK Medical Research Council (U117532067). AVSH and SCG are Jenner Investigators and are funded by the Wellcome Trust. SJD is a Junior Research Fellow ofMerton College, Oxford University. | SJD, SCG and AVSH are named inventors on patent applications covering malaria vectored vaccines and immunization regimes. Authorsfrom Okairo`s are employees of and/or shareholders in Okairo`s, which is developing vectored malaria vaccines. This does not alter the authors' adherence to all thePLoS ONE policies on sharing data and materials. | N/A | N/A | 10/12/2020 |
| Bliss CM | Assessment of novel vaccination regimens using viral vectored liver stage malaria vaccines encoding ME-TRAP | 2018 | Scientific Reports | This study was funded by the UK NIHR Biomedical Research Centre (BRC) with additional support from the Wellcome Trust. | Adrian Hill and Sarah Gilbert are named inventors on patent applications and patents relating to malaria vectored vaccines and immunization regimens. Stefano Colloca and Alfredo Nicosia are employees of and/or shareholders in ReiThera, which is developing vectored vaccines for malaria and other diseases. | NCT01364883, 2010-023824-26 | N/A | 10/12/2020 |
| Bliss CM | Targeting Antigen to the Surface of EVs Improves the In Vivo Immunogenicity of Human and Non-human Adenoviral Vaccines in Mice | 2020 | Molecular Therapy | This research project was supported in part by fundingfrom NIH/NIAID CEIRS (HHSN272201400008C), and by grantsawarded to L.C., including the US Graduate Women in Science(GWIS) 2017 Nell Mondy and Monique Braude Fellowship, a UKRoyal Society for Tropical Medicine and Hygiene small grant(GR000550), and by a Medical Research Fund pump-priming grantfrom the University of Oxford (MRF/TT2015/2150), UnitedKingdom. | A.V.S.H. is named as an inventor on a patent covering use ofChAdOx1-vectored vaccines and is a co-founder of, consultant to,and shareholder in Vaccitech plc, which is developing Ad-vectoredvaccines. The remaining authors declare no competing interests. | N/A | N/A | 10/12/2020 |
| Borthwick N | Vaccine-elicited human T cells recognizing conserved protein regions inhibit HIV-1 | 2014 | Molecular Therapy | The work was supported by Medical Research Council (MRC) UK and Department for International Development UK through an Experimental Medicine call II award G0701669 with contributions from the International AIDS Vaccine Initiative. HIV-1 infectious mo-lecular clones were obtained from Dr George Shaw, University of Pennsylvania. The FEC Control Peptide Pool was obtained through the NIH AIDS Reagent Program, Division of AIDS, NIAID, NIH (ref. no. 19626). | A.N. and S.C. who were employees and shareholders of Okairos and Advent during the conduct of the study, and are inventors on patents WO 2005071093 (A3), WO 2006133911 (A3) and WO 03031588 (A2), A.J.McM. who reports grants from MRC and NIH, per-sonal fees from International AIDS Vaccine Initiative SAB during the conduct of the study and is an inventor on patent WO 06123256, LC reports grants from MRC during the conduct of the study, and T.H. who reports grants from MRC and European and Developing Countries Clinical Trial Partnership obtained during the conduct of the study and is an inventor on patent WO 06123256. The other authors declare no conflict of interest. | No ID found | Tried searching clinicaltrials.gov but could not find registered trial and not in study | 16/12/20 |

| | | | | | | | | |
|---|---|---|---|---|---|---|---|---|
| Bowyer G | Activation-induced Markers Detect Vaccine-Specific CD4⁺ T Cell Responses Not Measured by Assays Conventionally Used in Clinical Trials | 2018 | Vaccines | The clinical trial was supported by funding from an Enhancement Award to a WellcomeTrust Strategic Award (to AVSH as PI) co-funded by the UK Medical Research Council, the UK Departmentfor International Development and the European and Developing Countries Clinical Trials Partnership, withadditional funding from the NIHR Oxford Biomedical Research Centre. | A.V.S.H. is a named inventor on patents relating to viral vectored vaccines. All other authorsdeclare no conflicts of interest. | NCT02451891, 2015-000593-3 | N/A | 21/12/2020 |
| Bowyer G | Reduced Ebola vaccine responses in CMV+ young adults is associated with expansion of CD57+KLRG1+ T cells | 2020 | JEM | The Oxford clinical trial was supported by funding from anEnhancement Award to a Wellcome Trust Strategic Award (toA.V.S. Hill as principal investigator) cofunded by the UK MedicalResearch Council, the UK Department for International Development, and the European and Developing Countries ClinicalTrials Parnership, with additional funding from the NationalInstitute for Health Research Oxford Biomedical Research Cen-tre.. GlaxoSmithKline Biologicals SA supplied the ChAd3–EBO-Zvaccine and had the opportunity to review this manuscript. TheMVA–EBO-Z vaccine was biomanufactured for these trials byEmergent Biosolutions under a contract from Oxford Universitywith funding from the same Enhancement Award. The Sene-galese trial was largely funded by a European Commission Ho-rizon 2020 program award, EbolaVac (http://www.ebolavac.eu), grant agreement no. 666085. | N/A | NCT02451891 | N/A | 18/12/20 |
| Boyd AC | Towards a universal vaccine for avian influenza: protective efficacy of modified Vaccinia virus Ankara and Adenovirus vaccines expressing conserved influenza antigens in chickens challenged with low pathogenic avian influenza virus | 2013 | Elselvier - Vaccine | The Biotechnology and Biological Sciences Research Council and Wellcome Trust are gratefully acknowledged for their financial support. | N/A | N/A | N/A | 16/12/20 |
| Bull TJ | Immunity, safety and protection of an Adenovirus 5 prime--Modified Vaccinia virus Ankara boost subunit vaccine against Mycobacterium avium subspecies paratuberculosis infection in calves | 2014 | Veterinary Research | This work was supported by Biotechnology and Biological Sciences ResearchCouncil (BBSRC) grants BB/H010556/1 and BB/H010718/1. Jayne Hope andIrene McGuinnes were supported by Institute Strategic Grant funding fromthe BBSRC | TJB is a minor shareholder in HAV Vaccines Ltd. | N/A | N/A | 20/12/2020 |
| Capone S | Immune responses against a liver-stage malaria antigen induced by simian adenoviral vector AdCh63 and MVA prime-boost immunisation in non-human primates | 2010 | Elselvier - Vaccine | This work was supported by the Wellcome Trust. AVSH is a Wellcome Trust Principal Research Fellow. | N/A | N/A | N/A | 16/12/20 |
| Cappuccini F | 5T4 oncofoetal glycoprotein: an old target for a novel prostate cancer immunotherapy | 2017 | Oncotarget | This work was supported by Oxford National Institutes for Health Research (NIHR) Biomedical Research Centre, UK (IR); the UK Medical Research Council CiC6 award (SS), the UK Wellcome Trust Senior Investigator's Award (AVSH) and the European Union's Seventh Framework Programme under grant agreement No. 602705 ( (FC, EP). | N/A | N/A | N/A | 21/12/2020 |
| Cappuccini F | Safety and immunogenicity of novel 5T4 viral vectored vaccination regimens in early stage prostate cancer: a phase I clinical trial | 2020 | Journal for ImmunoTherapy of Cancer | The VANCE clinical trial was supported by the European Union's Seventh Framework Programme under grant agreement no. 602705. | AVSH is a co- founder of and shareholder in Vaccitech Ltd which has supported the Oxford prostate cancer vaccine programme. | NCT02390063 | N/A | 18/12/20 |
| Cappuccini F | Immunogenicity and efficacy of the novel cancer vaccine based on simian adenovirus and MVA vectors alone and in combination with PD-1 mAb in a mouse model of prostate cancer | 2016 | Cancer Immunol Immunother | This work was supported by Oxford National Institutes for Health Research (NIHR) Biomedical Research Centre, UK (I. Redchenko); the UK Medical Research Council CiC6 award (S. Stribbling), the UK Wellcome Trust Senior Investigator's Award (A.V.S. Hill) and the European Union's Seventh Framework Programme under Grant Agreement No. 602705 (F. Cappuccini, E. Pollock) | N/A | N/A | N/A | 21/12/2020 |
| Carey JB | Microneedle-mediated immunization of an adenovirus-based malaria vaccine enhances antigen-specific antibody immunity and reduces anti-vector responses compared to the intradermal route | 2014 | Scientific Reports | This work wassupported by Enterprise Ireland (Commercialisation Fund, CFTD07/117) and ScienceFoundation Ireland (National Access Programme 70 and 170). AVSH and SJD are JennerInvestigators; and SJD is a UK MRC Career Development Fellow [G1000527] and ListerInstitute Research Prize Fellow. | The authors declare no competing financial interests.AVSH and SJD are named inventors on patent applications covering malaria vectoredvaccines and immunization regimes. JBC, AV, COM, AVSH, ACM are named inventors onpatent applications covering microneedle-mediated vaccine delivery. | N/A | N/A | 16/12/20 |
| Colloca S | Vaccine vectors derived from a large collection of simian adenoviruses induce potent cellular immunity across multiple species | 2012 | Science Transitional Medicine | This work was supported in part by Hepacivac (LSH-2005-1.2.4-2 project 037435)and the Wellcome Trust. A.V.S.H. was supported by a Wellcome Trust Principal Research Fel-lowship. E.B. was supported by Medical Research Council (UK).Author | N/A | HCV vaccine phase 1 trial (HCV001) was registered with the European Clinica lTrial database (EudraCT number: 2007-004259-12) and with the ClinicalTrials.gov database (ID: NCT01070407); malaria vaccine clinical trial (VAC043) was registered with the European Clinical Trial database (EudraCT number: 2010-023824-26) and with theClinicalTrials.gov database (ID: NCT01364883). | Clinical trials not this study but referenced studies | 16/12/2020 |
| Colston JM | Modification of Antigen Impacts on Memory Quality after Adenovirus Vaccination | 2016 | The Journal of Immunology | This work was supported by Wellcome Trust Grants 099897/Z/12/A and 091663MA. | N/A | N/A | N/A | 20/12/2020 |
| Cottingham MG | Preventing spontaneous genetic rearrangements in the transgene cassettes of adenovirus vectors | 2012 | Biotechnology and Bioengineering | This work was supported by the European Vaccine Initiative, theOxford Martin School, the Gates Foundation through the Foundationfor NIH, The Wellcome Trust, and the NIHR Oxford BiomedicalResearch Centre. We are grateful to Dr. Alexandra J. Spencer, JennerInstitute, University of Oxford, for assistance with immunology; toMr. Jake Matthews, Vector Core Facility, Jenner Institute, Universityof Oxford, for assistance with ChAd63-Pfs230; and to Dr. Nicola K.Green and Dr. Eleanor Berrie of the Clinical BiomanufacturingFacility, University of Oxford, for assistance and advice. Dr. DavidH. Wyllie, Jenner Institute, University of Oxford performed somecloning steps. | Conflict of interest: Okairo`s Srl and the University of Oxford hold intellectual propertyrelated to adenovirus vaccine vectors. | N/A | N/A | 16/12/20 |
| Coughlan L. | Heterologous Two-Dose Vaccination with Simian Adenovirus and Poxvirus Vectors Elicits Long-Lasting Cellular Immunity to Influenza Virus A in Healthy Adults | 2018 | Elselvier | Medical Research Council UK, NIHR BMRC Oxford. | SG and AVH are co-founders of Vaccitech, a company developingviral vectored vaccines including broadly cross-reactive influenza vac-cines. SS holds stock in SanofiPasteur which develops and markets in-fluenza vaccines. HdG received a travel grant from Abbvie. | NCT01818362 | N/A | 18/12/20 |
| de Barra E | A phase Ia study to assess the safety and immunogenicity of new malaria vaccine candidates ChAd63 CS administered alone and with MVA CS | 2014 | Plos One | The study was funded by a grant fromthe European Vaccine Initiative (EVI),http://www.euvaccine.eu/. Antibody assays were performed atWRAIR and were funded by the Malaria VaccineInitiative. This work was also supported by the UKNational Institute of Health Research through theOxford Biomedical Research Centre (A91301 AdultVaccine) and the Wellcome Trust (084113/Z/07/Z).SCG and AVSH are Jenner Investigators; AVSH issupported by a Wellcome Trust Principal ResearchFellowship (45488/Z/05); and SHH holds aWellcome Trust Research Training Fellowship(097940/Z/11/Z). The funders had no role in studydesign, data collection and analysis, decision topublish, or preparation of the manuscript. | The authors have read thejournal's policy and have the following conflicts:AVSH and SCG are named inventors on patentfilings related to immunisation with vectored malariavaccines, specifically WO2008/122769. None ofthese products have been commercialized. AN wasan employee of Okairos AG at the time of the study.Okairos AG has since been acquired by GSLVaccines, which now owns patents and patentapplications related to simian adenoviruses. None ofthe authors have had any consultancies relevant tothis paper. This conflict of interest does not alterthese authors' adherence to all PLOS ONE policieson sharing data and materials, as detailed online inthe guide for authors. | NCT01450280 | N/A | 16/12/20 |
| de Cassan SC | The requirement for potent adjuvants to enhance the immunogenicity and protective efficacy of protein vaccines can be overcome by prior immunization with a recombinant adenovirus | 2011 | The Journal of Immunology | S.C.d.C. is a Ph.D. student supported by the European Malaria Vaccine DevelopmentAssociation, a European Commission Framework Programme 6-funded consortium(Grant LSHP-CT-2007-037506). This work was also partly supported by the Well-come Trust (Grant 084113/Z/07/Z), the National Institute for Health Research Ox-ford Biomedical Research Centre, TRANSVAC, a European Commission FrameworkProgramme 7-funded consortium infrastructure grant, and grants to C.E.C. and V.S.C.from the Department of Biotechnology, Government of India, and European VaccineInitiative. C.E.C. is supported by a Tata Innovation Fellowship from the Department ofBiotechnology, Government of India. A.V.S.H. was supported by a Wellcome TrustPrincipal Research Fellowship. S.C.G., A.V.S.H., and S.J.D. are Jenner Investigators.S.J.D. is a Medical Research Council Career Development Fellow (Grant G1000527). | Disclosures: S.C.d.C., E.K.F., A.D.D., A.M., S.C.G., A.V.S.H., and S.J.D. are namedinventors on patent applications covering malaria-vectored vaccines andimmunization regimes. The other authors have no financial conflicts ofinterest. | N/A | N/A | 16/12/20 |
| Dicks MD | Differential immunogenicity between HAdV-5 and chimpanzee adenovirus vector ChAdOx1 is independent of fiber and penton RGD loop sequences in mice | 2015 | Scientific Reports | This work has been funded by a grant from the Wellcome Trust (095540/Z/11/Z). AVSH and SCG are Jenner Institute Investigators, AJS and MGC are James Martin Fellows. | MDJD, SCG, AVSH, and MGC are named inventors on a patent application describing the ChAdOx1 vector (US2015044766). | N/A | N/A | 20/12/2020 |
| Dicks MD | The relative magnitude of transgene-specific adaptive immune responses induced by human and chimpanzee adenovirus vectors differs between laboratory animals and a target species | 2015 | Vaccine | This work has been funded by the Wellcome Trust (095540) with addi-tional funding from the Foundation for the National Institute of Health through the Grand Challenges in Global Health Initiative (HILL05GCGH0). MDJD received additional funding from the Euro-pean Malaria Vaccine Development Association (EMVDA). AVSH is a Wellcome Trust Principal Research Fellow. EG and BC were funded by the Biotechnology and Biological Sciences Research Council BBS/E/I/00001373, UnitedKingdom. | MDJD,SCG,AVSH,andMGCarenamedinventorsonapatentapplicationdescribingtheChAdOx1vector(PCTApplicationNo.PCT/GB2012/000467). | N/A | N/A | 20/12/2020 |

| | | | | | | | | |
|---|---|---|---|---|---|---|---|---|
| Dicks MD | A novel chimpanzee adenovirus vector with low human seroprevalence: improved systems for vector derivation and comparative immunogenicity | 2012 | Plos One | This work has been funded by grants from the Foundation for the National Institute of Health through the Grand Challenges in Global Health Initiative,with additional funding from the Wellcome Trust. MDJD received additional funding from the European Malaria Vaccine Development Association (EMVDA). MGCis a fellow of the Oxford Martin School Institute for Vaccine Design. SCG is a Jenner Investigator. AVSH is Director of the Jenner Institute and a Wellcome TrustPrincipal Research Fellow. The funders had no role in study design, data collection and analysis, decision to publish, or preparation of the manuscript. | Competing interests: MDJD, SCG, AVSH, and MGC are named inventors on a patent application describing the ChAdY25/ChAdOx1 vector (GB PatentApplication No. 1108879.6). This does not alter the authors' adherence to all the PLoS ONE policies on sharing data and materials. | N/A | N/A | 16/12/20 |
| Draper SJ | Enhancing blood-stage malaria subunit vaccine immunogenicity in rhesus macaques by combining adenovirus, poxvirus, and protein-in-adjuvant vaccines | 2010 | The Journal of Immunology | This work was funded by the Wellcome Trust and the European Malaria Vaccine Devlopment Association, a European Commission FP6-funded consortium. S.J.D. is a Junior Research Fellow of Merton College, Oxford, United Kingdom. S.C.G. and A.V.S.H. are Jenner Investigators, and A.V.S.H is also a Wellcome Trust Principal Research Fellow. | Disclosures: S.J.D, S.C.G, and A.V.S.H. are named inventors on patent applications covering malaria vectored vaccines and immunization regiments. S.C., M.N., A.F., and A.N. are employees of and/or shareholders in Okairòs, which is developing vecotored vaccines for malarian and other diseases. | N/A | N/A | 16/12/20 |
| Dulal P | Potency of a thermostabilised chimpanzee adenovirus Rift Valley Fever vaccine in cattle | 2016 | Vaccine | This study was conducted with support from a grant from the Bill & Melinda Gates Foundation Grand Challenges ExplorationI nitiative to GMW (OPP1096893) and a Wellcome Trust fellowship to GMW (WT098635). B.C. and A.V.S.H. are Jenner Inves-tigators. | N/A | N/A | N/A | 20/12/2020 |
| Elias SC | Analysis of human B-cell responses following ChAd63-MVA MSP1 and AMA1 immunization and controlled malaria infection | 2013 | Immnunology | This work was supported by the UK Medical ResearchCouncil (grant number G0700735); the European MalariaVaccine Development Association, a European Commis-sion FP6-funded consortium (LSHP-CT-2007-037506);the UK National Institute of Health Research through theOxford Biomedical Research Centre; the Wellcome Trust(084113/Z/07/Z); and by EVIMalaR funded by the Euro-pean Community's Seventh Framework Programme (FP7/2007–2013) (Grant agreement No. 242095). AVSH andSJD are Jenner Investigators; and SJD is a UK MRCCareer Development Fellow (G1000527) and Lister Insti-tute Prize Research Fellow. | SCdC, KAC, AVSH and SJD are named inventors on pat-ent applications covering malaria vaccines and immuniza-tion regimens. | NCT01373879, NCT01142765, NCT01003314, NCT01095055 | N/A | 20/12/2020 |
| Ewer K | A Monovalent Chimpanzee Adenovirus Ebola Vaccine Boosted with MVA | 2016 | New England Journal of Medicine | Supported by the Wellcome Trust, the United Kingdom Medi-cal Research Council, the United Kingdom Department for Inter-national Development, and the United Kingdom National Insti-tute for Health Research Oxford Biomedical Research Centre. The National Health Service Blood and Transplant and Public Health England provided funding for the competition ELISA. The ChAd3 vaccine was provided by the Vaccine Research Center of the National Institute of Allergy and Infectious Diseases (NIAID) and GlaxoSmithKline. MVA-BN Filo was produced under a con-tract (FBS-004-009) between the NIAID and Fisher BioServices and a contract (HHSN272200800044C) between the National Institutes of Health and Fisher Bioservices. | Dr. Ballou, Dr. De Ryck report personal fees and other support from GlaxoSmithKline outside the submitted work. Dr. Colloca, Dr. Cortese, Dr. Nicosia reports a pending patent related to chimpanzee adenoviral-vector based filovirus vaccine (WO/2011/130627). Dr. Draper reports grant support from the UK Medical Research Council during the conduct of the study, and non-financial support from GlaxoSmithKline/Okairos outside the submitted work. In addition, Dr. Draper reports pending patents related to viral vector immunogenic compositions (WO 2012042279 A3) and adenoviral vectors encoding a pathogen or tumor antigen (WO 2008122811 A2). Dr. Gilbert reports patents related to prime boost immunization with viral vectors. Dr. Hill reports a patent related to heterologous prime-boost immunization, licensed to Oxford BioMedica. Ms. Lella reports other support from the NIHR during the conduct of the study. Dr. Levine reports grant support from Oxford University during the conduct of the study. Dr. Pollard reports grant support from the Wellcome Trust during the conduct of the study, and grant support from GlaxoSmithKline outside the submitted work. | N/A | N/A | 20/12/2020 |
| Ewer K | Protective CD8 þ T-cell immunity to human malaria induced by chimpanzee adenovirus-MVA immunisation | 2013 | Nature Communications | The study was funded by grants from the UK MRC,the NIHR through the Oxford Biomedical Research Centre, and the Wellcome Trust.AVSH was supported by a Wellcome Trust Principal Research Fellowship. A.L.G. wassupported by a grant from the MRC (G0600424). A.V.S.H., A.R.-S., S.J.D. and S.C.G.are Jenner Institute Investigators; A.V.S.H. is a Wellcome Trust and NIHR SeniorInvestigator. | Sarah Gilbert, Arturo Reyes-Sandoval, Anna Goodman,Geraldine O'Hara and Adrian Hill are named inventors on patent applications coveringmalaria vectored vaccines and immunization regimes including: WO/2008/122811-Adenoviral vectors encoding a pathogen or tumour antigen and WO/2008/122769-Adenoviral vector encoding malaria antigen. Authors from Okairos are employees ofand/or share holders in Okairos which is developing vectored malaria vaccines. All otherauthors declare no competing financial interests. | N/A | N/A | 16/12/20 |
| Fedosyuk S | Simian adenovirus vector production for early-phase clinical trials: A simple method applicable to multiple serotypes and using entirely disposable product-contact components | 2019 | Elselvier Vaccine | This work was supported by Merck KGaA, the UK MedicalResearch Council (grant MR/P017339/1), and the UK Engineeringand Physical Sciences Research Council (grant EP/R013756/1).ADD is supported by the Wellcome Trust [grants 201477/Z/16/Zand 204826/Z/16/Z] and is a Jenner Investigator. The study wasperformed in collaboration between the University of Oxford andMerck KGaA; both partners reviewed the manuscript prior to sub-mission. The other funders had no input to the design of the studyor decision to publish. | ADD, SJM, and SCG are named inventors on patent filings relat-ing to the use of simian adenoviruses, but not directly related tothe work described here. SCG is a founder of Vaccitech Ltd, whichdevelops adenovirus-vectored vaccines. | N/A | N/A | 10/12/2020 |
| Folegatti P.M. | Safety and immunogenicity of a candidate Middle East respiratory syndrome coronavirus viral-vectored vaccine: a dose-escalation, open-label, non-randomised, uncontrolled, phase 1 trial | 2020 | The Lancet | UK Department of Health and Social Care, using UK Aid funding, managed by the UK National Institute for Health Research. This project was funded by the UK Department of Health and Social Care (project number 16/107/01). The views expressed are those of the authors and not necessarily those of the Department of Health and Social Care. The work was supported by the UK National Institute for Health Research through the Oxford Biomedical Research Centre. The Coalition for Epidemic Preparedness Innovations provided funding for the extended 12 months of follow-up in this study. This study was also partially supported by the Coordenacao de Aperfeicoamento de Pessoal de Nivel Superior, Brazil (finance code 001). The pseudovirus neutralising antibody work was funded by a grant from the Korean Ministry of Health and Welfare (HI15C2971) | AH and SG are co-founders of, consultants for, and shareholders in Vaccitech, which is developing adenoviral vectored vaccines. PMF and TL are consultants for Vaccitech. All other authors declare no competing interests. | NCT03399578 | N/A | 18/12/20 |
| Folegatti P.M. | Safety and immunogenicity of the ChAdOx1 nCoV-19 vaccine against SARS-CoV-2: a preliminary report of a phase 1/2, single-blind, randomised controlled trial | 2020 | The Lancet | UK Research and Innovation, Coalition for Epidemic Preparedness Innovations, National Institute for Health Research (NIHR), NIHR Oxford Biomedical Research Centre, Thames Valley and South Midland's NIHR Clinical Research Network, and the German Center for Infection Research (DZIF), Partner site Gießen-Marburg-Langen. This work is funded by UK Research and Innovation (MC_PC_19055), Engineering and Physical Sciences Research Council (EP/R013756/1), Coalition for Epidemic Preparedness Innovations (CEPI), the National Institute for Health Research (NIHR), the NIHR Oxford Biomedical Research Centre, and the German Center for Infection Research (DZIF), Partner site Gießen-Marburg-Langen. Additional resources for study delivery were provided by NIHR Southampton Clinical Research Facility and NIHR Southampton Biomedical Research Centre, University Hospital Southampton NHS Foundation Trust; the NIHR Imperial Clinical Research Facility; and NIHR North West London, South London, Wessex, and West of England Local Clinical Research Networks and NIHR Oxford Health Biomedical Research Centre. PMF received funding from the Coordenacao de Aperfeicoamento de Pessoal de Nivel Superior, Brazil (finance code 001). Development of SARS-CoV-2 reagents was partially supported by the US National Institute of | SCG is co-founder and board member of Vaccitech (collaborators in the early development of this vaccine candidate) and named as an inventor on apatent covering use of ChAdOx1-vectored vaccines and a patent application covering this SARS-CoV-2 vaccine. TL is named as an inventor on a patent application covering this SARS-CoV-2 vaccine and consultant to Vaccitech. PMF is a consultant to Vaccitech. AJP is Chair of the UK Department of Health and Social Care's Joint Committee on Vaccination & Immunisation (JCVI), but does not participate in policy advice on coronavirus vaccines, and is a member of the WHO Strategic Advisory Group of Experts (SAGE). AVSH is a co-founder of and consultant to Vaccitech and is named as an inventor on a patent covering design and use of ChAdOx1-vectored vaccines. AF is a member of JCVI, Chair of the WHO European Technical Advisory Group of Experts on Immunisation, an ex-officiomember of WHO SAGE working group on COVID-19 vaccines, and acting director of National Institute for Health Research West of England Local Clinical Research Network. KMP reports grants from the NIHR Imperial Biomedical Research Centre and Gilead Sciences, and personal fees from Sanofi Pasteur, outside of the submitted work. MS reports grants from Janssen, GlaxoSmithKline, Medimmune, | NCT04324606. The study is ongoing, and was registered at ISRCTN, 15281137, and ClinicalTrials.gov, NCT04324606. | AZ mentioned in acknowledgements - facilitated and funded some assays and reviewed the data from the study and the final manuscript before submission. In declaration of interests says royalties paid to AZ for vectors. | 18/12/20 |
| Folegatti PM | Safety and Immunogenicity of a Novel Recombinant Simian Adenovirus ChAdOx2 as a Vectored Vaccine | 2019 | MDPI | This research was funded by HAV Vaccines Ltd. The research was supported by the National Institutefor Health Research (NIHR) Oxford Biomedical Research Centre (BRC). The views expressed are those of theauthor(s) and not necessarily those of the NHS, the NIHR or the Department of Health. | J.H.-T. is the Chief Scientific and Medical Officer for HAV Vaccines Ltd. S.C.G. andA.V.S.H. are co-founders of, consultants to and shareholders in Vaccitech plc which is developing adenoviralvectored vaccines. | NCT03027193 | N/A | 18/12/20 |
| Forbes EK | T cell responses induced by adenoviral vectored vaccines can be adjuvanted by fusion of antigen to the oligomerization domain of C4b-binding protein | 2012 | Plos One | This work was funded by the Wellcome Trust and the EMVDA (European Malaria Vaccine Development Association, a European Commission FP6-funded consortium). The research leading to these results has also received funding from the European Community's Seventh Framework Programme (FP7/2007–2013) under grant agreement No 242095. In addition, this work was supported in part by the Division of Intramural Research, National Institutes of Allergy andInfectious Diseases, National Institutes of Health, and also by the PATH Malaria Vaccine Initiative who support the GIA Reference Center. AVSH and SJD are JennerInvestigators. AVSH is a Wellcome Trust Principal Research Fellow. SJD is a MRC Career Development Fellow. The funders had no role in study design, datacollection and analysis, decision to publish, or preparation of the manuscript, except that the design of the rabbit study and the GIA analysis was performedfollowing discussion with PATH MVI. | Author contributions: . Contributed reagents/materials/analysis tools: SB ALGRJP FH. | N/A | N/A | 16/12/20 |
| Gilbert SC | Enhanced CD8 T cell immunogenicity and protective efficacy in a mouse malaria model using a recombinant adenoviral vaccine in heterologous prime-boost immunisation regimes | 2002 | Elselvier - Vaccine | Wellcome Trust and the European Commission(IC18-CT95-0019 TMR fellowship to J.S.) for support | N/A | N/A | N/A | 14/12/20 |
| Gola A | Prime and target immunization protects against liver-stage malaria in mice | 2018 | Science Transitional Medicine | A.G. is funded by the Wellcome Trust and by the Intramural Program of NIAID (NIH). B.R.H. is funded from the European Union Seventh Framework Programme FP7/2012-2106 under grant agreement 316655 (VACTRAIN). A.V.S.H. is a Wellcome Trust and National Institute of Health Research (NIHR) senior investigator. This work was in part funded by a Wellcome Trust Senior Investigator award (to A.V.S.H.) and a Wellcome Trust Enhancement award (to A.V.S.H.) for the clinical trial and also was supported in part by the Intramural Research Program of NIAID (NIH) (to R.N.G. and S.U.). The clinical trial was supported in part by funding from the UK NIHR Oxford Biomedical Research Centre | A.V.S.H., A.G., A.A.W., and A.M.S. are inventors on a patent application PCT/GB2017/051009 submitted by the Oxford University Innovation Limited that covers prime and target vaccination with viral vectors | N/A | N/A | 21/12/2020 |
| Goodman AL | A viral vectored prime-boost immunization regime targeting the malaria Pfs25 antigen induces transmission-blocking activity | 2011 | Plos One | This work was supported primarily by grant G0600424 from the Medical Research Council (ALG) and in addition by Transmolbloc (EU FP7) and BBSRC(award number LDAD_P15820). SJD and AVH are Jenner Investigators. SJD is a MRC Career Development Fellow. AVH is a Wellcome Trust Principal ResearchFellow. ALG was an MRC clinical training fellow whilst she undertook this research. The funders had no role in study design, data collection and analysis, decisionto publish, or preparation of the manuscript. http://www.mrc.ac.uk/index.htm. http://www.bbsrc.ac.uk/. http://www.wellcome.ac.uk/. | N/A | N/A | N/A | 16/12/20 |

| | | | | | | | | |
|---|---|---|---|---|---|---|---|---|
| Graham SP | Evaluation of the immunogenicity of prime-boost vaccination with the replication-deficient viral vectored COVID-19 vaccine candidate ChAdOx1 nCoV-19 | 2020 | Nature partner journals | This study was supported by UKRI Engineering and Physical Sciences ResearchCouncil (EPSRC) award EP/R013756/1 (VaxHub), UKRI Biotechnology and BiologicalSciences Research Council (BBSRC) Institute Strategic Programme and Core CapabilityGrants to The Pirbright Institute (BBS/E/I/00007031, BBS/E/I/00007034, BBS/E/I/00007037 and BBS/E/I/00007039), and the Bill and Melinda Gates Foundationsupported'Pirbright Livestock Antibody Hub'(Grant No. OPP1215550). Developmentof SARS-CoV-2 reagents was partially supported by the NIAID Centers of Excellencefor Influenza Research and Surveillance (CEIRS) contract HHSN272201400008C andEPSRC Grant No. EP/S025243/1 to the Rosalind Franklin Institute. A.L., G.W., C.B., A.B.,and V.M. are supported by the UK Department for Environment Food & Rural Affairs.We thank V. Clark, H. Gray, and R. Snaith for animal husbandry and the JennerInstitute Vector Core Facility for assistance, and The Pirbright Institute Animal ServicesTeam for animal care and provision of samples. | S.C.G. and T.L. are named on a patent application covering ChAdOx1 nCoV-19. Theremaining authors declare no competing interests. The funders played no role in theconceptualisation, design, data collection, analysis, decision to publish, or preparationof the manuscript. | N/A | N/A | 18/12/20 |
| Halbroth BR | Development of a Molecular Adjuvant to Enhance Antigen-Specific CD8(+) T Cell Responses | 2018 | Scientific Reports | BRH received funding from the European Union Seventh Framework Programme FP7/2012–2016 under grant agreement n° 316655 (VACTRAIN). Additional funding was provided by a Wellcome Trust Senior Investigator award to AVSH and a Wellcome Trust Strategic Award supporting the viral vector core facility. Further funding was provided by a Gates Grand Challenges in Global Health award through the Foundation for NIH (to AVSH). | A.V.S.H. is a named investigator on US 12/595 574 and UK PCT/GB2008/01262 novel adenovirus patent applications covering malaria vectored vaccines and immunisation regimens; A.V.S.H., A.J.S., M.J.C. and B.R.H. are named investigators on UK PCT/GB2014/053596, a novel molecular adjuvant application. | N/A | N/A | 10/12/2020 |
| Hodgson SH | Combining viral vectored and protein-in-adjuvant vaccines against the blood-stage malaria antigen AMA1: report on a phase 1a clinical trial | 2014 | Molecular Therapy | This work was supported by the EMVDA (European Malaria Vaccine Development Association), a European Commission (EC) FP6-funded consortium (LSHP-CT-2007–037506); the UK National Institute of Health Research through the Oxford Biomedical Research Centre (NIHR-BRC) (A91301 Adult Vaccine); the Wellcome Trust (084113/Z/07/Z); and EVIMalaR, an EC FP7-funded programme (Grant agreement No. 242095). The GIA work was supported by the PATH Malaria Vaccine Initiative and the Intramural Program of the National Institutes of Health, National Institute of Allergy and Infectious Diseases. | .D.D., J.J.I., S.C.dC., A.V.S.H., and S.J.D. are named inventors on patent applica-tions covering malaria vaccines and immunization regimens. A.N. is an employee of and/or shareholder in Okairòs, which is developing vectored vaccines for malaria and other diseases. | NCT01351948 | N/A | 20/12/2020 |
| Hodgson SH | Evaluation of the efficacy of ChAd63-MVA vectored vaccines expressing circumsporozoite protein and ME-TRAP against controlled human malaria infection in malaria-naive individuals | 2015 | The Journal of Infectious Diseases | This work was supported by the PATH MalariaVaccine Initiative, the United Kingdom National Institute of Health Re-search, through the Oxford Biomedical Research Centre (grant A91301,Adult Vaccine), and the Wellcome Trust (grants 084113/Z/07/Z and45488/Z/05 to A. V. S. H. and grant 097940/Z/11/Z to S. H. H.). | A. V. S. H. and S. C. G. are named inven-tors on patent applications covering malaria vectored vaccines and immu-nization regimens. S. C. and A. N. are employees of and/or shareholders inOkairos, whichis developing vectored vaccines formalariaandother diseases.All other authors report no potential conflicts. | NCT01623557 | N/A | 20/12/2020 |
| Kelly C | Chronic hepatitis C viral infection subverts vaccine-induced T-cell immunity in humans | 2016 | Hepatology | Supported by a Medical Research Council (MRC) UK Development Clinical Scheme award (G0701694); by the Wellcome Trust, the Oxford NHIRBRC, and the U19 grant (2U19AI082630-06, to C.K. and P.K.); by an MRC CASE studentship (to L.S.). E.B. is funded as an MRC Senior ClinicalScientist and is supported by the Oxford NHIR BRC, the Oxford Martin School, and the Jenner Institute. | Dr. Colloca, Dr. Folgori, Dr. Cortese, and Dr. Nicosia are named inventors on patent applications covering hepatitisCvirus-vectored vaccines and chimpanzee adenovirus vectors (WO 2006133911 [A3] hepatitis C virus nucleic acid vaccine, WO 2005071093 [A3]chimpanzee adenovirus vaccine carrier, WO 03031588 [A2] hepatitis C virus vaccine). Dr. Hill is a coinventor on patent filings and applicationsrelated to heterologous prime-boost immunizations. | NCT01094873, 2008-006127-32) | N/A | 20/12/2020 |
| Kimani D | Translating the immunogenicity of prime-boost immunization with ChAd63 and MVA ME-TRAP from malaria naive to malaria-endemic populations | 2014 | Molecular Therapy | This work was supported by an award from the European and Developing Countries Clinical Trials Partnership (EDCTP) and was performed by the Malaria Vectored Vaccines Consortium (MVVC), a four and a half year integrated project funded by the European and Developing Countries Clinical Trials Partnership (EDCTP, grant number IP.2008.31100.001). N.I.V. is an employee of the European Vaccine Initiative (EVI). E.V.I. is the coordinator of the EDCTP funded MVVC project (grant number IP.2008.31100.001). E.V.I. supports salaries of the MVVC project in kind. The work was also supported by the UK National Institute of Health Research through the Oxford Biomedical Research Centre (http://www.oxfordbrc.org/) [A91301 Adult Vaccine], the Wellcome Trust (http://www.wellcome.ac.uk/) (084113/Z/07/Z) and the Medical Research Council. S.H.H. holds a Wellcome Trust research training fellowship (097940/Z/11/Z). The funders had no role in study design, data collection and analysis, decision to publish, or preparation of the manuscript. | A.V.S.H. is a named inventor on patent applications on malaria vectored vaccines and immunization regimens. Authors from Okairòs are employees of and/or shareholders in Okairòs, which is devel-oping vectored vaccines for malaria and other diseases. | N/A | N/A | 16/12/20 |
| Lambe T | Immunity against heterosubtypic influenza virus induced by adenovirus and MVA expressing nucleoprotein and matrix protein-1 | 2013 | Scientific Reports | AvL was funded by a fellowship of theDr. Saal van Zwanenberg Stichting. Dr. Lambe is supported by the Oxford Martin School.SCG is a Jenner Investigator. | N/A | N/A | N/A | 16/12/20 |
| Longley RJ | Comparative assessment of vaccine vectors encoding ten malaria antigens identifies two protective liver-stage candidates | 2015 | Scientific Reports | This work has been funded by a grant from the Wellcome Trust (095540/Z/11/Z) with additional funding by the Rhodes Trust and a Nuffield Department of Medicine Studentship to support RJL and EVIMalR studentship to support AMS. AVSH is a Jenner Institute Investigator; AJS is a James Martin Fellow. CJJ was supported by a grant of the European Community's Seventh Framework Programme (FP7/2007-2013) under grant agreement number 242095. | AVSH is a named investigator on US 12/595 574 and UK PCT/GB2008/01262 novel adenovirus patent applications covering malaria vectored vaccines and immunization regimens; RJL, AMS, CJJ, SMK, AJS and AVSH are named investigators on filed patent (1318084.9) for novel malaria antigens | N/A | N/A | 20/12/2020 |
| Longley RJ | Assessment of the Plasmodium falciparum Preerythrocytic Antigen UIS3 as a Potential Candidate for a Malaria Vaccine | 2017 | Infection and Immunity | Funding for manufacture and quality control release and stability studies of Sanaria's PfSPZ challenge was provided by the National Institute of Allergy and Infectious Diseases (http://www.niaid.nih.gov ) under grant R44AI058375 (Universal Attenuated Malaria Sporozoite Vaccine and Challenge System). The present work was funded by a grant from the Wellcome Trust (grant 095540/Z/11/Z) to A.V.S.H., with additional funding by the Rhodes Trust and a Nuffield Department of Medicine Studentship to support R.J.L. The human CHMI study (VAC049) was supported by the UK National Institute of Health Research through the Oxford Biomedical Research Centre (http://www.oxfordbrc.org/ ) (A91301 adult vaccine program) and the Wellcome Trust (grant 084113/Z/07/Z). B.R.H. receives funding from the European Union Seventh Framework Programme FP7/2012-2016 under grant agreement 316655 (VACTRAIN). S.H.H. is a Wellcome Trust Clinical Research Fellow (grant 097940/Z/11/Z). A.V.S.H. is a Jenner Institute Investigator, and A.J.S. is a James Martin Fellow. | A.V.S.H. is a named investigator on novel adenovirus patent applications U.S. 12/595 574 and UK PCT/GB2008/01262, covering malaria vectored vaccines and immunization regimens. R.J.L., A.J.S., and A.V.S.H. are named investigators on patent PCT/GB2014/053077, identifying novel malaria vaccine antigens. | NCT01465048 | N/A | 21/12/2020 |
| López-Camacho C | Rational Zika vaccine design via the modulation of antigen membrane anchors in chimpanzee adenoviral vectors | 2018 | Nature Communications | This report is independent research funded by the UKDepartment of Health and Social Care through Innovate UK"New vaccines for globalepidemics: development and manufacture"grant No. 972216 (A.R.-S.), and also fundedfrom an ODA budget (Global Health (ODA), 16/107/05—Design, development andGMP manufacture of a Zika vaccine) (A.H.P. and A.R-S). The views expressed in thispublication are those of the author(s) and not necessarily those of the Department ofHealth and Social Care. We also acknowledge funding by the UK Medical ResearchCouncil [MC_UU_12014 (A.H.P. and A.K.) and MR/N017552/1 (A.K.)]. JuthathipMongkolsapaya is supported by an MRC-Newton Fund grant, Gavin Screaton is aWellcome Trust Senior Investigator. | A.R.-S. and C.L.-C. are co-inventors of the Zika vaccines describedin this manuscript,filed by Oxford University Innovation Limited in the InternationalPatent Application No. PCT/GB2017/052220 Zika Vaccine; A.V.S.H. and S.C.G. are co-inventors on a patent application (WO/2012/172277) on the ChAdOx1 viral vectorfiledby Oxford University Innovation. The remaining authors declare no competing interests. | N/A | N/A | 21/12/2020 |
| McMahon M | Vaccination With Viral Vectors Expressing Chimeric Hemagglutinin, NP and M1 Antigens Protects Ferrets Against Influenza Virus Challenge | 2019 | Vaccine | The study was fundedby an MRC Biomedical Catalyst DPFS_DCS award (MR/N006372/1).In addition, this study was partially funded by the NIAID Centers ofExcellence for Influenza Research and Surveillance contract(CEIRS, HHSN272201400008C, grant AI109946 and grant AI142046-01) | The Icahn School of Medicine at Mount Sinaihas filed patent applications regarding influenza virus vaccines withFK being aninventor. SG is an inventor on patents covering ChAdOx1 and MVA-NP + M1,filed and owned by the University of Oxford, and is a co-founder ofand consultantto Vaccitech, a University of Oxford spin-out company which is undertakingadvanced clinical development of viral vectored influenza vaccines. | N/A | N/A | 10/12/2020 |
| Mensah VA | Safety and Immunogenicity of Malaria Vectored Vaccines Given with Routine Expanded Program on Immunization Vaccines in Gambian Infants and Neonates: A Randomized Controlled Trial | 2017 | Frontiers in Immunology | This work was supported by a Strategic Primer grant award from the European and Developing Countries Clinical Trials Partnership (EDCTP, grant number SP.2011.41304.025); with co-funding from Swedish International Development Cooperation Agency (Sida); UK Medical Research Council; Irish Aid, Department of Foreign Affairs and Trade, Ireland; and Bundesministerium für Bildung und Forschung (BMBF), Germany. European Vaccine Initiative (EVI) coordinated the pro-ject under phase two of Malaria Vectored Vaccines Consortium (MVVC 2). Additional funding for the Oxford collaborators was provided by the Wellcome Trust and the UK National Institute of Health Research | The following authors have declared that no conflict of interest exists: VM, SR, EK, AmN, FO, CB, GB, YJ, RR, NV, FD, OL, AL, BF, BK, BC, SG, EC, KE, EI, and MA. AH is a named inventor on patent applications on malaria vectored vaccines and immunization regimens. RC and AlN are employees and/or shareholders in ReiThera, which develops vectored vaccines for malaria and other diseases. The authors declare that the research was conducted in the absence of any commercial or financial relationships that could be construed as a potential conflict of interest. The reviewer AL declared a past collaboration with four of the authors, OL, NV, FD, and AH, to the handling Editor. | NCT02083887 | N/A | 21/12/2020 |
| Mensah VA | Safety, Immunogenicity and Efficacy of Prime-Boost Vaccination with ChAd63 and MVA Encoding ME-TRAP against Plasmodium falciparum Infection in Adults in Senegal | 2016 | Plos One | This study was supported by an award from the European and Developing Countries Clinical Trials Partnership (EDCTP) and was performed by the Malaria Vectored Vaccines Consortium (MVVC), an integrated project funded by EDCTP (grantnumberIP.2008.31100.001).Co-funding was also provided by the Medical Research Council UK, the Swedish International Development Cooperation Agency (Sida) and Irish Aid. The work was also supported by the Dakar University Cheikh Anta Diop. | We have the following interests. Adrian V.S. Hill is a named inventor on patent applications and patents on malaria vectored vaccines and immunisation regimes including the following (WO2008/122769, Adenoviral vector encoding malaria antigen; and WO 2008/122811 Novel adenovirus vectors).Egeruan Imoukhuede and Ines Petersen were employees of EVI at the time of the study,which supports the development and testing of malaria vaccines.Nicola Viebigis an employee of EVI and Odile Leroyis executive director of EVI. Authors from Rei Thera (formerly Okairos) are employees of and/or shareholders in Rei Thera, which is developing vectored vaccines for malaria and other diseases. Alfredo Nicosia was employed by Rei Thera (formerly Okairos) at the time of thestudy. | PACTR201-303-000-499-409. (African Pan Trial Registry) | N/A | 21/12/2020 |
| Munster VJ | Protective efficacy of a novel simian adenovirus vaccine against lethal MERS-CoV challenge in a transgenic human DPP4 mouse model | 2017 | Npj Vaccines | This work is published withthe permission of the Director of the Kenya Medical Research Institute, and wassupported by the Intramural Research Program of the National Institute of Allergyand Infectious Diseases (NIAID), National Institutes of Health (NIH) and a grant fromthe UK Medical Research Council Confidence in Concept scheme to GMW throughthe LSTM Tropical Infectious Disease Consortium | S.C.G. is a co-founder of, consultant to and shareholder inVaccitech plc, which is developing a vectored MERS vaccine. Remaining authorsdeclares that they have no competingfinancial interests. | N/A | N/A | 21/12/2020 |

| | | | | | | | | |
|---|---|---|---|---|---|---|---|---|
| Nébié I | Assessment of chimpanzee adenovirus serotype 63 neutralizing antibodies prior to evaluation of a candidate malaria vaccine regimen based on viral vectors | 2014 | Clinical and Vaccine Immunology | This work was supported by an award from the European and Developing Countries Clinical Trials Partnership (EDCTP) and wasperformed by the Malaria Vectored Vaccines Consortium (MVVC), an integrated project funded by EDCTP (grant numberIP.2008.31100.001). | N/A | N/A | N/A | 16/12/20 |
| O'Hara GA | Clinical assessment of a recombinant simian adenovirus ChAd63: a potent new vaccine vector | 2012 | The Journal of Infectious Diseases | Financial support. This work was supported by an Experimental Medicine grant from the UK Medical Research Council (grant number G0502018) with additional support from the UK National Institute for Health Research Oxford Biomedical Research Centre and the Wellcome Trust. No funding bodies had any role in study design, data collection andanalysis, decision to publish, or preparation of the manuscript. | Potential conflicts of interest: S. G., A. R.-S., A. G., G. O. H., and A. H.are named inventors on patent applications covering malaria-vectoredvaccines and immunization regimes. Authors from Okairos are employeesof and/or shareholders in Okairos, which is developing vectored malariavaccines. All other authors report no potential conflicts.All authors have submitted the ICMJE Form for Disclosure of PotentialConflicts of Interest. Conflicts that the editors consider relevant to thecontent of the manuscript have been disclosed. | NCT00890019 | N/A | 16/12/20 |
| Ogwang C | Prime-boost vaccination with chimpanzee adenovirus and modified vaccinia Ankara encoding TRAP provides partial protection against Plasmodium falciparum infection in Kenyan adults | 2015 | Science Transitional Medicine | This work was funded by the European and Developing Countries Clinical TrialsPartnership, grant number IP.2008.31100.001, to the Malaria Vectored Vaccines Consortium(MVVC), and coordinated by the European Vaccine Initiative (EVI). P.B. is jointly funded by theU.K. Medical Research Council (MRC) and the U.K. Department for International Development(DFID) under the MRC/DFID Concordat agreement. | A.V.S.H., A.N., and S.G. are listed as inventors on patent filings related to heterologous prime-boost immunization and specific malaria vaccines. E.B.I. was an employee of EVI at the time ofthis study, which supports the development and testing of malaria vaccines. N.K.V. is an em-ployee of EVI, and O.L. is executive director of EVI. A.N. is an employee of Okairos and consultsfor GlaxoSmithKline. The other authors declare no competing interests. | NCT01666925, PACTR 201202000356208 | N/A | 20/12/2020 |
| Ogwang C | Safety and immunogenicity of heterologous prime-boost immunisation with Plasmodium falciparum malaria candidate vaccines, ChAd63 ME-TRAP and MVA ME-TRAP, in healthy Gambian and Kenyan adults | 2013 | Plos One | This work was performed by the Malaria Vectored Vaccines Consortium (MVVC), a four year integrated project funded by the European and DevelopingCountries Clinical Trials Partnership (EDCTP). The work was also supported by the UK National Institute of Health Research through the Oxford BiomedicalResearch Centre (http://www.oxfordbrc.org/) [A91301 Adult Vaccine], the Wellcome Trust (http://www.wellcome.ac.uk/) [084113/Z/07/Z] and theMedicalResearch Council. The funders had no role in study design, data collection and analysis, decision to publish, or preparation of the manuscript. | Competing Interests:AH is a named inventor on the following patent applications on malaria vectored vaccines and immunization regimes (WO2008/122769,Adenoviral vector encoding malaria antigen; and WO 2008/122811 Novel adenovirus vectors). Authors from Okairo`s are employees of and/or shareholders inOkairo`s, which is developing vectored vaccines for malaria and other diseases. This does not alter the authors' adherence to all the PLOS ONE policies on sharingdata and materials. | Pactr.org PACTR201002000177182 8 Pactr.org PACTR201008000221638 ClinicalTrials.gov NCT01373879ClinicalTrials.gov NCT01379430 | N/A | 16/12/20 |
| Payne RO | Human vaccination against Plasmodium vivax Duffy-binding protein induces strain-transcending antibodies | 2017 | JCI Insight | This work was supported by a UK Medical Research Council (MRC) grant (number G1100086). The study was also supported in part by UK National Institute of Health Research (NIHR) infrastructure through the NIHR Oxford Biomedical Research Centre and the Wellcome Trust (084113/Z/07/Z). DL was supported by the Rhodes Trust. TAR holds a Wellcome Trust Research Training Fellowship (108734/Z/15/Z). SCdC was a PhD student supported by the European Malaria Vaccine Development Association, a European Commission Framework Programme 6–funded consortium (grant LSHP-CT-2007-037506). TDO is sup-ported by the Wellcome Trust (WT 098051). JSM is supported by an NHMRC Practitioner Fellowship (number 1041802). AVSH and SJD are Jenner Investigators. SJD is a Lister Institute Research Prize Fellow and a Wellcome Trust Senior Fellow (grant number 106917/Z/15/Z). | S.C. de Cassan, M.K. Higgins, A.V.S. Hill, and S.J. Draper are named inventors on patent applications (patent nos. GB1413530.5, GB1016471.3, and WO/2008/122811) covering malaria vaccines and immunization regimens. A. Nicosia was an employee of and shareholder in Okairòs (since acquired by GlaxoSmithKline), which is developing vectored vaccines for a number of diseases. T. Jørgensen and W.A. de Jongh are employees of, and W.A. de Jongh is a shareholder in, ExpreS2ion Biotechnologies, which has developed and is marketing the ExpreS2 cell expression platform. C.E. Chitnis is a named inventor on a patent covering PvDBP_RII (patent no. WO/1996/040766). | NCT01816113 | N/A | 21/12/2020 |
| Payne RO | Human vaccination against RH5 induces neutralizing antimalarial antibodies that inhibit RH5 invasion complex interactions | 2017 | JCI Insight | This work was supported by funding from the European Union Seventh Framework Programme (FP7/2007-2013) under the grant agreement for MultiMalVax (number 305282). The study was also supported in part by UK NIHR infrastructure through the NIHR Oxford Biomedical Research Centre; the MAVARE-CA program funded by Danida (the Consultative Committee for Development Research, Denmark); and the Wellcome Trust (grant numbers 084113/Z/07/Z and 206194). The GIA work was supported by the United States Agency for International Development (USAID) and the Intramural Program of the NIH, National Institute of Allergy and Infectious Diseases. DGWA holds a UK MRC iCASE PhD Studentship (MR/K017632/1); JSM is supported by a National Health and Medical Research Council (NHMRC) Practitioner Fellowship (10418020); ADD held a Wellcome Trust Training Fellowship for Clinicians in Basic Sciences (089455/2/09/z); SB, AVSH, and SJD are Jenner Investigators; and SJD is a Lister Institute Research Prize Fellow and a Wellcome Trust Senior Fellow (106917/Z/15/Z) | S.J. Draper is a named inventor on patent applications relating to RH5 and/or other malaria vaccines and immunization regimens; is a cofounder of, shareholder in, and consultant for SpyBiotech; and declares research funding support from Pfizer and GSK BioPharm. A.D. Douglas, G.J. Wright, and A.V.S. Hill are named inventors on patent applications relating to RH5 and/or other malaria vaccines and immunization regimens. L. Siani and S. Di Marco are employees of ReiThera (formerly Okairos), which is currently developing vectored vaccines for a number of diseases. J. Vekemans was an employee of GSK, which has acquired the ChAd63 vector from Okairos. R. Ashfield is a director of Ducentis and holds shares in the company, which is developing a therapy for autoimmune disease. A.M. Minassian has an immediate family member who is an inventor on patents relating to RH5 and/or other malaria vaccines and immunization regimens and who is a cofounder of, shareholder in, and consultant for SpyBiotech. S. Biswas is a cofounder and CEO of, and shareholder in, SpyBiotech and is a contributor in a patent application relating to multimerisation technology. J. Jin is a cofounder of and shareholder in SpyBiotech. | NCT02181088 | N/A | 21/12/2020 |
| Pearson FE | Dry-coated live viral vector vaccines delivered by nanopatch microprojections retain long-term thermostability and induce transgene-specific T cell responses in mice | 2013 | Plos One | This work has been supported by a UK Medical Research Council Capacity Building Studentship (G0600311, www.mrc.ac.uk) and by the Bill andMelinda Gates Foundation (003436, www.gatesfoundation.org). The funders had no role in study design, data collection and analysis, decision to publish, orpreparation of the manuscript. | Competing Interests:Authors MLC, GJPF, XC and MAFK are either inventors or contributors to a patent filings related to the NanopatchTM technology, that isnow licensed to Vaxxas Pty Ltd. These are detailed in Supporting Information - Table S2. Authors MLC, SRY, GJPF and MAFK have employment with Vaxxas. MAFKis a member of the Vaxxas board. There are no further patents, products in development or marketed products to declare. This does not alter the authors'adherence to all the PLOS ONE policies on sharing data and materials. | N/A | N/A | 16/12/20 |
| Pearson FE | Induction of CD8(+) T cell responses and protective efficacy following microneedle-mediated delivery of a live adenovirus-vectored malaria vaccine | 2015 | Vaccine | This study was funded by Enterprise Ireland (CFTD07/117,http://www.enterprise-ireland.com), Science Foundation Ireland (NAP156andNAP170,www.sfi.ie) and the Medical Research Coun-cil,United Kingdom (G0600311,www.mrc.ac.uk). | None | N/A | N/A | 20/12/2020 |
| Pérez de Val B | A multi-antigenic adenoviral-vectored vaccine improves BCG-induced protection of goats against pulmonary tuberculosis infection and prevents disease progression | 2013 | Plos One | The study was funded by the European Community's 7th Framework Programme (FP7-KBBE-2007-1-3-04: TB-STEP project under grant agreement212414). The funders had no role in study design, data collection and analysis, decision to publish, or preparation of the manuscript. | N/A | N/A | N/A | 16/12/20 |
| Rampling T | Safety and High Level Efficacy of the Combination Malaria Vaccine Regimen of RTS,S/AS01B With Chimpanzee Adenovirus 63 and Modified Vaccinia Ankara Vectored Vaccines Expressing ME-TRAP | 2016 | The Journal of Infectious Diseases | This work was supported by the PATH MalariaVaccine Initiative and by the United Kingdom NIHR, through the NIHROxford Biomedical Research Centre, the Southampton NIHR WellcomeTrust Clinical Research Facility, and the Imperial College NIHR WellcomeTrust Clinical Research Facility. | . V. S. H. and S. C. G. are named inven-tors on patent applications covering malaria vectored vaccines and immu-nization regimens. D. M., M. L., and R. W. B. are employees of GSK, which isdeveloping vectored vaccines for malaria and other diseases. S. N. F. acts onbehalf of the University of Southampton/University Hospital SouthamptonNational Health Service Foundation trust as chief and principal investigatorfor clinical trials sponsored by vaccine manufacturers, including GSK, butreceives no personal payments for the work. | NCT01883609 | N/A | 20/12/2020 |
| Rampling T | Safety and efficacy of novel malaria vaccine regimens of RTS,S/AS01B alone, or with concomitant ChAd63-MVA-vectored vaccines expressing ME-TRAP | 2018 | Nature partner journals | This work was funded primarily by the PATH Malaria VaccineInitiative (MVI); in addition, the work was supported by the United Kingdom NationalInstitute of Health Research (NIHR) infrastructure, through the NIHR OxfordBiomedical Research Centre, the Southampton NIHR Wellcome Trust Clinical ResearchFacility, and the Imperial College NIHR Wellcome Trust Clinical Research Facility; thisarticle/paper/report presents independent research funded by PATH MVI andsupported by the NIHR CRF and BRC at Imperial College Healthcare NHS Trust. Theviews expressed are those of the author(s) and not necessarily those of PATH MVI, theNHS, the NIHR, or the Department of Health. | A.V.S.H. and S.C.G. are named inventors on patent applicationsand patents relating to malaria vectored vaccines and immunization regimens. D.M.,M.L., and R.W.B are employees of GSK, which is developing vaccines for malaria andother diseases. S.N.F. acts on behalf of the University of Southampton/UniversityHospital Southampton NHS Foundation trust as chief and principal investigator forclinical trials Sponsored by vaccine manufacturers including GSK, but receives nopersonal payments for the work. The other authors declare no competing interests. | NCT02252640 | N/A | 18/12/20 |
| Reyes-Sandoval A | Prime-boost immunization with adenoviral and modified vaccinia virus Ankara vectors enhances the durability and polyfunctionality of protective malaria CD8+ T-cell responses | 2010 | Human Vaccines | Work in the Oxford malaria vaccine program is supported by the Wellcome Trust, the UK Medical Research Council, the UK National Institute for Health Research through the Oxford Biomedical Research Centre, the European Commission, the Gates Foundation through a Grand Challenges in Global Health award from the Foundation for NIH, the European Malaria Vaccine Initiative, the Jenner Vaccine Foundation and the European and Developing Countries Clinical Trials Partnership. | N/A | N/A | N/A | 10/12/2020 |
| Reyes-Sandoval A | Mixed vector immunization with recombinant adenovirus and MVA can improve vaccine efficacy while decreasing antivector immunity | 2010 | The American Society of Gene & Cell Therapy | We thank the Jenner Institute's vector core facility for providing the viral-vectored vaccines and Dr Helen McShane for providing the ad-enoviral and MVA vectors expressing antigen 85A. We are also grateful to Andrew Williams for providing the P. berghei parasites and the NIH tetramer facility (MHC tetramer core facility, Emory University Vaccine Center, Atlanta, GA) for preparing the Pb9 tetramer. The transgenic parasites were kindly provided by Dr Oliver Billker from Wellcome Trust Sanger Institute, Hinxton, UK. This work was funded by Wellcome Trust Principal Research Fellowship award; Grant Number: 076438; The National Institute for Health Research Oxford Biomedical Research Centre Program and Grand Challenges in Global Health. A.R.-S. is a Scientific Leadership Fellow of the Nuffield Department of Medicine and a Wellcome Trust Fellow. C.S.R. is supported by Oxford Biomedical Research Center, The Oxford Martin School and Meningitis UK. | N/A | N/A | N/A | 16/12/20 |
| Reyes-Sandoval A | Single-dose immunogenicity and protective efficacy of simian adenoviral vectors against Plasmodium berghei | 2008 | European Jounral of Immunology | The work was supported by a Wellcome Trust Principal Research Fellowship award grant number 076438 toA.V.S.H. | N/A | N/A | N/A | 16/12/20 |

| | | | | | | | | |
|---|---|---|---|---|---|---|---|---|
| Rollier CS | Modification of Adenovirus vaccine vector-induced immune responses by expression of a signalling molecule | 2020 | Nature | This work was funded by a grant from the Foundation for the National Institutes of Health through the Grand Challenges in Global Health Initiative of the Gates Foundation, with additional support from the Wellcome Trust. Non-human primate studies were supported by National Center for Research Resources (NCRR) grant # P51 RR000167, and were conducted at a facility constructed with support from grants RR15459 and RR020141. The authors wish to acknowledge the expert contribution of Dr Matthew G. Cottingham, and the expert help provided by the Animal Services Unit, and the Immunology Services Unit of the Wisconsin National Primate Research Center, in particular D. Watkins and E. Rakasz. CR is supported by the NIHR Biomedical Research Centre, Oxford. CR, SGC and AVSH are Jenner Institute Investigators and AVSH is a Wellcome Trust and NIHR Senior Investigator. The funding sources had no involvement in study design; collection, analysis and interpretation of data, in the writing of the report and in the decision to submit the article for publication. | A.V.S.H. and S.G.C. are names as co-inventors on patents related to recombinant viral vectors for malaria and other indications. | N/A | N/A | 18/12/20 |
| Salman AM | Rational development of a protective P. vivax vaccine evaluated with transgenic rodent parasite challenge models | 2017 | Scientific Reports | he work was funded by a Wellcome Trust Career Development Fellowship award, grant number 097395/Z/11/Z, to A.R.-S, who is also a Jenner Investigator and an Oxford Martin Fellow. Funding was also provided by the Medical Research Council, through a DPFS grant (MR/N019008/1) to A.R.-S. Ahmed M. Salman was funded by EVIMalaR's Program funding (FP7/2007–2013) under grant agreement N° 242095. Adrian Hill is supported by a Wellcome Trust grant number 095540/Z/11/Z and is a Jenner Investigator and an Oxford Martin Fellow. | N/A | N/A | N/A | 21/12/2020 |
| Sebastian S | A Multi-Filovirus Vaccine Candidate: Co-Expression<br>of Ebola, Sudan, and Marburg Antigens in a<br>Single Vector | 2020 | MDPI | This research was funded by Innovate UK (Novel multivalent vaccines against haemorrhagic fevers,971510) and MRC (Confidence in Concept CiC 2015-16, MC_PC_15040, Liverpool School of Tropical Medicine). | Teresa Lambe and Sarah Gilbert are Jenner Investigators. All other authors declare noconflicts of interest. Views expressed are those of the authors and do not necessarily reflect those of the employinginstitutions. The funders had no role in the design of the study; in the collection, analyses, or interpretation ofdata; in the writing of the manuscript, or in the decision to publish the results. | N/A | N/A | 18/12/20 |
| Sheehy SH | Phase Ia clinical evaluation of the safety and immunogenicity of the Plasmodium falciparum blood-stage antigen AMA1 in ChAd63 and MVA vaccine vectors | 2012 | Plos One | This work was supported by the European Malaria Vaccine Development Association (EMVDA), a European Commission FP6-funded consortium (http://www.emvda.org/) [LSHP-CT-2007-037506]; the UK National Institute of Health Research through the Oxford Biomedical Research Centre (http://www.oxfordbrc.org/) [A91301 Adult Vaccine]; and the Wellcome Trust (http://www.wellcome.ac.uk/) [084113/Z/07/Z]. The GIA work was supported by the PATH MalariaVaccineInitiative (MVI; http://www.malariavaccine.org/) and the Intramural Program of the National Institutes of Health, National Institute of Allergyand InfectiousDiseases (http://www.niaid.nih.gov/Pages/default.aspx). CJAD holds a Wellcome Trust Research Training Fellowship [RTEI0]; SCG, AVSH and SJD are JennerInvestigators; AVSH was supported by a Wellcome Trust Principal Research Fellowship [45488/Z/05]; and SJD is a UK Medical Research Council CareerDevelopment Fellow [G1000527]. The funders had no role in study design, data collection and analysis, decision to publish, or preparation of the manuscript. | Competing Interests: AJS, MDJD, SCG, AVSH and SJD are named inventors on US 12/595 574 and UK PCT/GB2008/01262, US 12/595 351 and UK PCT/GB2008/01271 novel adenovirus patent applications covering malaria vectored vaccines and immunization regimes. This does not alter our adherence to all the PLoS ONEpolicies on sharing data and materials. Authors from Okairo`s are employees of and/or shareholders in Okairos, which is developing vectored vaccines for malaria and other diseases. This does not alter our adherence to all the PLoS ONE policies on sharing data and materials. | NCT01095055 | N/A | 16/12/20 |
| Sheehy SH | ChAd63-MVA-vectored blood-stage malaria vaccines targeting MSP1 and AMA1: assessment of efficacy against mosquito bite challenge in humans | 2012 | The American Society of Gene & Cell Therapy | This work was supported by the European Malaria Vaccine Development Association, a European Commission FP6-funded consortium (LSHP-CT-2007-037506); the UK Medical Research Council (grant no. G0700735); the UK National Institute of Health Research through the Oxford Biomedical Research Centre (A91301 Adult Vaccine), and the Southampton NIHR Wellcome Trust Clinical Research Facility; the Wellcome Trust (084113/Z/07/Z); and EVIMalaR, an European Commission FP7-funded programme (grant agreement no. 242095). The growth inhibitory activity work was sup-ported by the PATH Malaria Vaccine Initiative and the Intramural Program of the National Institutes of Health, National Institute of Allergy and Infectious Diseases. S.C.G., A.V.S.H., and S.J.D. are Jenner Investigators; A.V.S.H. was supported by a Wellcome Trust Principal Research Fellowship (45488/Z/05); C.J.A.D. holds a Wellcome Trust Research Training Fellowship (094449/Z/10/Z); and S.J.D. is a UK Medical Research Council Career Development Fellow (G1000527). The funders had no role in study design, data collection and analysis, decision to publish, or prepa-ration of the manuscript. A.D.D., S.C.G., A.V.S.H., and S.J.D. are named inventors on patent applications covering malaria-vectored vaccines and immunization [illegible] | N/A | N/A | N/A | 16/12/20 |
| Sheehy SH | Phase Ia clinical evaluation of the Plasmodium falciparum blood-stage antigen MSP1 in ChAd63 and MVA vaccine vectors | 2011 | The Journal of Immunology | This work was supported by the UK Medical Research Council (MRC) (grant number G0700735), the European Malaria Vaccine Development Association (EMVDA), a European Commission FP6-funded consortium (LSHP-CT-2007-037506), the UK National Institute of Health Research through Oxford Biomedical Research Centre (A91301 Adult Vaccine) and the Wellcome Trust (084113/Z/07/Z). The GIA work was supported by the PATH Malaria Vaccine Initiative (MVI) and the Intramural Program of the National Institues of Health, National Institue of Allergy and Infectious Diseases. S.C.G., A.V.S.H, and S.J.D. are Jenner Investigators; A.V.S.H. was supported by a Wellcome Trust Principal Research Fellowship (45488/Z/05) and S.J.D. is a MRC Career Development Fellow (G1000157). A.R.W., S.C.G., A.V.S.H., and S.J.D. are named inventors on patent applications coverin malaria vectored vaccines and immunization regimes. Authors from Okairòs are employees of and/or shareholders in Okairòs which is developing vecotred vaccines for malaria and other diseases. | N/A | N/A | N/A | 16/12/20 |
| Spencer AJ | Enhanced vaccine-induced CD8+ T cell responses to malaria antigen ME-TRAP by fusion to MHC class ii invariant chain | 2014 | Plos One | This work has been funded by grants from the Foundation for the National Institute of Health through the Grand Challenges in Global Health Initiative(HILL05GCGH0) with additional funding from the Wellcome Trust (095540/Z/11/Z). Non-human primate studies were supported by National Center for ResearchResources (NCRR) grant#P51 RR000167, and was conducted at a facility constructed with support from grants RR15459 and RR020141. The funding bodies didnot have a role in the study design, data collectionand analysis, decision to publish or preparation of the manuscript. Co-authors S. Capone, S. Colloca, AF, RC andAN are employees of Okairos. Okairos provided support in the form of salaries for the authors, but did not have any additional role in the study design, datacollection and analysis, decision to publish, or preparation of the manuscript. The specific roles of all authors are articulated in the 'author contributions' section. | AVSH is a named inventor on WO/2008/122811-Adenoviral vectors encoding a pathogen or tumour antigen and WO/2008/122769-Adenoviral vector encoding malaria antigen. S. Colloca, AF, RC, and AN are named inventors on patent application WO 2005071093 (A3)-Chimpanzee adenovirusvaccine carriers. Authors from Okairos were employees of and/or share holders in Okairos, this does not alter the authors' adherence to all the PLOS ONE policieson sharing data and materials. | N/A | N/A | 16/12/20 |
| Stedman A | Safety and efficacy of ChAdOx1 RVF vaccine against Rift Valley fever in pregnant sheep and goats | 2019 | Nature partner journals | This study was funded by the UK Department of Health and Social Care (UK DHSC,Project 16/107/03). G.M.W. is supported by an Oak foundation fellowship and aWellcome Trust grant (203077_Z_16_Z). S.C.G., B.C. and A.V.S.H. are JennerInvestigators. The funders had no role in study design, data collection and analysis,decision to publish, or preparation of the manuscript. The views expressed in thispublication are those of the authors and do not necessarily reflect those of the UKDHSC. We thank Dr. Connie Schmaljohn (USAMRIID, Fort Detrick, MD) for providingthe 4-D4 antibody. This paper is published with the permission of the PirbrightInstitute and the Director of the Kenya Medical Research Institute. | S.C.G. and A.V.S.H. are co-founders of, consultants to and shareholders in VaccitechLtd., which is developing ChAdOx1-vectored vaccines. The remaining authors declareno competing interests. | N/A | N/A | 18/12/20 |
| Svitek N | An Ad/MVA vectored Theileria parva antigen induces schizont-specific CD8(+) central memory T cells and confers partial protection against a lethal challenge | 2018 | Nature partner journals | This work was funded by the Bill and Melinda Gates Foundation (BMGF) and theDepartment for International Development (DFID) of the United Kingdom[OPP1078791], the Norman Borlaug Commemorative Research Initiative, an initiativebetween the Feed the Future program of United States Agency for InternationalDevelopment (USAID), USA and United States Department of Agriculture (USDA)-Agricultural Research Service, USA (58-5348-2-117F) and through funding from theCGIAR Research Program on Livestock and Fish [CRP 3.7] | N/A | N/A | N/A | 18/12/20 |
| Swadling L | A human vaccine strategy based on chimpanzee adenoviral and MVA vectors that primes, boosts, and sustains functional HCV-specific T cell memory | 2014 | Science Transitional Medicine | Funding:Supported by the Medical ResearchCouncil (MRC) UK and the European Union (Framework VI; HEPACIVAC) for funding the studyand the manufacture of MVA-NSmut through an MRC UK DCS (Developmental Clinical Studies)award. E.B. is supported by the MRC as a Senior Clinical Fellow, the Oxford Martin Schools, andNational Institute for Health Research Oxford Biomedical Research Centre. L. Swadling is supported by an MRC CASE studentship. | Competing interests:S. Colloca, A.F., R.C., and A.N.are named inventors on patent applications covering HCV-vectored vaccines and chimpanzeeadenovirus vectors [WO 2006133911 (A3) hepatitis C virus nucleic acid vaccine, WO 2005071093(A3) chimpanzee adenovirus vaccine carriers, WO 03031588 (A2) hepatitis C virus vaccine]. P.K. hasacted as a consultant to Tibotec and Pfizer on antiviral therapy. The other authors declare thatthey have no competing interests | N/A | N/A | 16/12/20 |
| Swadling L | | 2016 | Vaccines | Supported by the Medical Research Council (MRC) UK and the European Union (FrameworkVI; HEPACIVAC) for funding the study and the manufacture of MVA-NSmut through an MRC UK DCS(Developmental Clinical Studies) award (G0701694). Christabel Kelly and Paul Klenerman are supported by theWellcome Trust, the Oxford NHIR BRC and the U19 grant (2U19AI082630-06). Leo Swadling is supported by anMRC CASE studentship. Elleanor Barnes is funded as an MRC Senior Clinical Fellow, and is supported by theOxford NIHR BRC, the Oxford Martin School and the Jenner Institute. | Stefano Colloca, Antonella Folgori, Riccardo Cortese and Alfredo Nicosia are namedinventors on patent applications covering HCV-vectored vaccines and chimpanzee adenovirus vectors[WO 2006133911 (A3) hepatitis C virus nucleic acid vaccine, WO 2005071093 (A3) chimpanzee adenovirusvaccine carriers, WO 03031588 (A2) hepatitis C virus vaccine]. Adrian Hill is a co-inventor on patent filings andapplications related to heterologous prime-boost immunisations. | NCT01296451 | N/A | 20/12/2020 |
| Tapia MD | Use of ChAd3-EBO-Z Ebola virus vaccine in Malian and US adults, and boosting of Malian adults with MVA-BN-Filo: a phase 1, single-blind, randomised trial, a phase 1b, open-label and double-blind, dose-escalation trial, and a nested, randomised, double-blind, placebo-controlled trial | 2016 | The Lancet Infectious Diseases | This study was funded by a Wellcome Trust Strategic Translation Award, with funding contributions from the Medical Research Council UK, and the Department for International Development UK supported the primary vaccination of 80 Malian participants with ChAd3-EBO-Z. A Wellcome Trust Enhancement Award funded the boosting of 52 Malians with MVA-BN-Filo or saline. Funding for priming immunisation of the Malian participants with the ChAd3-EBO-Z vaccine and for data management was provided by the National Cancer Institute, the Frederick National Laboratory for Cancer Research, Federal Funds from the National Institute of Allergy and Infectious Diseases, and a contract (number HHSN261200800001E) awarded to Leidos Biomedical Research. The US trial was funded by the Vaccine Research Center, National Institute of Allergy and Infectious Diseases, through a contract to the EMMES Corporation. | FR, IDR, and WRB are employees of GlaxoSmithKline and manufacture non-replicating chimpanzee adenovirus 3-based vaccines. NS is a named inventor on patents related to ChAd3-EBO-Z. AVSH is named as an inventor on patents using heterologous prime boost immunisation with viral vectors, such as those assessed in this report. MML serves as a member of the Scientifi c Advisory Working Group to the Vaccine Research Center, National Institute of Allergy and Infectious Diseases, and of the Vaccine Research Center Board of Scientifi c Counselors. | NCT02231866, NCT02267109 | N/A | 20/12/2020 |

| | | | | | | | | |
|---|---|---|---|---|---|---|---|---|
| Tiono AB | First field efficacy trial of the ChAd63 MVA ME-TRAP vectored malaria vaccine candidate in 5-17 months old infants and children | 2018 | Plos One | This work was supported by an award from the European and Developing Countries Clinical Trials Partnership (EDCTP) and was performed by the Malaria Vectored Vaccines Consortium (MVVC), a 5-year integrated project (Grant number IP.2008.31100.001). The European Vaccine Initiative (EVI) was the coordinator of the EDCTP-funded MVVC project. Co-funding was also provided by the Swedish International Development Cooperation Agency (SIDA), the Austrian Federal Ministry of Science and Research, and Irish Aid. Additional support for the Oxford clinical trials team was provided by the UK NIHR through the Oxford Biomedical Research Centre | : AVSH is a named inventor on patent applications and issued patents relating to malaria vectored vaccines and immunization regimes. This does not alter the author's adherence to all the PLOS ONE policies on sharing data and materials. | NCT01635647, PACTR201208000404131. | N/A | 21/12/2020 |
| Tuthill M. | Results from ADVANCE: A phase I/II open-label non-randomised safety and efficacy study of the viral vectored ChAdOx1-MVA 5T4 (VTP-800) vaccine in combination with PD-1 checkpoint blockade in metastatic prostate cancer | 2020 | Annals of Oncology (Abstracts) | European Union Seventh Framework Programme under grant AgreementNo. 602705 (Project IMPROVE) and Vaccitech Ltd. | M. Tuthill: Advisory/Consultancy: Vaccitech Limited; Advisory/Consultancy, SpeakerBureau/Expert testimony: BMS; Advisory/Consultancy, Speaker Bureau/Expert testimony: Pfizer;Advisory/Consultancy, Speaker Bureau/Expert testimony: Novartis, ; Advisory/Consultancy, SpeakerBureau/Expert testimony: Janssen; Advisory/Consultancy, Speaker Bureau/Expert testimony: Roche;Advisory/Consultancy: Lilly; Advisory/Consultancy: Oxford Vacmedix, ; Speaker Bureau/Expert tes-timony: Astellas; Speaker Bureau/Expert testimony: Genomic Health; Speaker Bureau/Expert testi-mony: Esai; Speaker Bureau/Expert testimony: Everything Genetic; Travel/Accommodation/Expenses: EUSA Pharma. T. Evans: Leadership role, Shareholder/Stockholder/Stock options: Vacci-tech Limited. A. Protheroe: Speaker Bureau/Expert testimony: BMS. A.V.S. Hill: Advisory/Consultancy,Founder Vaccitech: Vaccitech Limited. All other authors have declared no conflicts of interest. | NCT03815942 | N/A | 18/12/20 |
| Utrilla-Trigo S | Heterologous Combination of ChAdOx1 and MVA Vectors Expressing Protein NS1 as Vaccination Strategy to Induce Durable and Cross-Protective CD8+ T Cell Immunity to Bluetongue Virus | 2020 | MDPI | This work was supported by grants AGL2017-82570-R from the Spanish Ministry of Science and EUHorizon 2020 Program (European Comission Grant Agreement NO.727393-PALE-Blu). SUT was a recipient of apredoctoral fellowship from the Instituto Nacional de Investigación y Tecnología Agraria y Alimentaria, Centro deInvestigación en Sanidad Animal (program FPI-SGIT-201). | N/A | N/A | N/A | 18/12/20 |
| van Doremalen N | A single dose of ChAdOx1 MERS provides protective immunity in rhesus macaques | 2020 | Science Advances | This work was supported by the Intramural Research Program of the National Institute of Allergy and Infectious Diseases (NIAID), NIH (1ZIAAI001179-01) and the Department of Health and Social Care using UK Aid funding managed by the NIHR.S.C.G. is a Jenner investigator. The views expressed in this publication are those of the author(s) and not necessarily those of the Department of Health and Social Care. | S.C.G. is a board member of Vaccitech and named as an inventor on a patent covering use of ChAdOx1-vectored vaccines. The other authors declare that they have no competing interests. | N/A | N/A | 18/12/20 |
| van Doremalen N | A single-dose ChAdOx1-vectored vaccine provides complete protection against Nipah Bangladesh and Malaysia in Syrian golden hamsters | 2019 | PLOS | This work was supported by the Intramural Research Program of the National Institute of Allergy and Infectious Diseases(NIAID),National Institutes of Health(NIH).TAB issupportedby the Medical Research Council(MR/L009528/1). SCG is a Jenner investigator.The sponsors did not play a role in study design. | I have read the journal's policy and the authorsof this manuscript have the following competing interests:SCG is named as an inventor on a patent covering use of ChAdOx1-vectored vaccines.The remaining authors declare no conflict of interest. | N/A | N/A | 18/12/20 |
| van Doremalen N | ChAdOx1 nCoV-19 vaccine prevents SARS-CoV-2 pneumonia in rhesus macaques | 2020 | Nature | This work was supported by the Intramural Research Program of the National Institute of Allergy and Infectious Diseases (NIAID), National Institutes of Health (NIH) (1ZIAAI001179-01) and the Department of Health and Social Care using UK Aid funding managed by the NIHR. | S.C.G. is a board member of Vaccitech and named as an inventor on a patent covering the use of ChAdOx1-vector-based vaccines and a patent application covering a SARS-CoV-2 (nCoV-19) vaccine (UK patent application no. 2003670.3). T.L. is named as an inventor on a patent application covering a SARS-CoV-2 (nCoV-19) vaccine (UK patent application no. 2003670.3). The University of Oxford and Vaccitech, having joint rights in the vaccine, entered into a partnership with AstraZeneca in April 2020 for further development, large-scale manufacture and global supply of the vaccine. Equitable access to the vaccine is a key component of the partnership. Neither Oxford University nor Vaccitech will receive any royalties during the pandemic period or from any sales of the vaccine in developing countries. The other authors declare no competing interests. | N/A | AZ in competing interests | 18/12/20 |
| Venkatraman N | Safety and Immunogenicity of a Heterologous Prime-Boost Ebola Virus Vaccine Regimen in Healthy Adults in the United Kingdom and Senegal | 2019 | The Journal of Infectious Diseases | This work was supported by the Wellcome Trust, the UK Medical Research Council, the UK Department for International Development, and the UK NIHR Oxford Biomedical Research Centre. The Good Manufacturing Practice biomanufacture of MVA-EBO-Z and the UK clinical trial was supported by an Enhancement Award to a Wellcome Trust Strategic Award (to A. V. S. H. as principal investigator) co-funded by the Medical Research Council, the Department for International Development, and the European and Developing Countries Clinical Trials Partnership (grant number 106325/Z/14/A), with additional support from the NIHR Oxford Biomedical Research Centre. The Senegal trial was largely funded by a European Commission Horizon 2020 program award EbolaVac (www.ebolavac.eu; grant agreement 666085), which provided additional resources for the MVA-EBO-Z biomanufacture. This study was also supported by GlaxoSmithKline Biologicals SA. | F. R. and W. R. B. are employ-ees of GSK and own restricted shares of the company. S. C. G., K. E., and A. V. S. H. are named inventors on patents relating to viral vector vaccines for malaria and other diseases. F. R. and W. R. B. are employees of GSK, which is developing vectored vaccines for Ebola and other diseases. All other authors report no potential conflicts of interest. All authors have submitted the ICMJE Form for Disclosure of Potential Conflicts of Interest. Conflicts that the editors consider relevant to the content of the manuscript have been disclosed. | NCT02451891; NCT02485912 | N/A | 18/12/20 |
| Walker AS | Modeling Combinations of Pre-erythrocytic Plasmodium falciparum Malaria Vaccines | 2015 | American Journal of Tropical Medicine and Hygiene | Andrew S. Walker, José Lourenço andSunetra Gupta are funded by the European Research Council (ERCAdvanced Grant—Diversity). AVSH is a Jenner Investigator andWellcome Trust and NIHR Senior Investigator. | N/A | N/A | N/A | 20/12/2020 |
| Wang C | A simian-adenovirus-vectored rabies vaccine suitable for thermostabilisation and clinical development for low-cost single-dose pre-exposure prophylaxis | 2018 | Plos Neglected Tropical Diseases | This work has been supported by the UK Medical Research Council including Confidence in Concept (grantsMC_PC_13073and MR/P017339/1). SJD, AVSH and ADD are Jenner Investigators; SJD is also a Lister Institute Research Prize Fellow and a Wellcome Trust Senior Fellow (grant106917/Z/15/Z). ADD is supported by the Wellcome Trust (grant201477/Z/16/Z). The funders had no role in study design, data collection and analysis, decision to publish, or preparation of the manuscript. | AVSH and ADD are named inventors on a patent application relating to the development of the ChAdOx2 vector. AVSH and SJD are named inventors on a patent relating to the use of the intron-containing promoter used in ChAdOx2 RabG. The Universityof Oxford and the Wistar Institute have entered into a partnership to share any future revenue from development of the ChAdOx2 RabG vaccine. | N/A | N/A | 21/12/2020 |
| Warimwe GM | Chimpanzee Adenovirus Vaccine Provides Multispecies Protection against Rift Valley Fever | 2016 | Scientific Reports | This work was conducted with the support from the University of Oxford, a Wellcome Trust fellowship to GMW (WT098635) and grant from the Bill & Melinda Gates Foundation through the Grand Challenges Exploration Initiative to GMW (OPP1096893). B.C., S.C.G. and A.V.S.H. are Jenner Investigators. | N/A | N/A | N/A | 20/12/2020 |
| Warimwe GMLorenzo | Immunogenicity and efficacy of a chimpanzee adenovirus-vectored Rift Valley fever vaccine in mice | 2013 | Virology Journal | This work was supported by a Wellcome Trust fellowship in PublicHealth and Tropical Medicine to GMW (grant no. 098635/B/12/Z) and by aSpanish Ministry of Science grant (no. AGL2011-22485) to AB. ELG is a recipi-ent of a pre-doctoral fellowship program from the Spanish Ministry of Sci-ence. SCG and AVSH are Jenner Investigators. | MDJD, MGC, SCG and AVSH are named inventors on a patent applicationdescribing the ChAdOx1 vector (GB Patent application number 1108879.6).All other authors declare that they have no competing interests. | N/A | N/A | 16/12/20 |
| Yoshida K | Adenovirus-prime and baculovirus-boost heterologous immunization achieves sterile protection against malaria sporozoite challenge in a murine model | 2018 | Scientific Reports | This work was supported, in part, by a Grant-in-Aid for Young Scientists (B) (JSPS KAKENHI grant number 26860278), a grant from the Ohyama Health Foundation, and Cooperative Research Grants from NEKKEN, 2014 (grant number 26-6) and 2015 (grant number 27-5) to M.I.; by Grants-in-Aid for Scientific Research (B) (JSPS KAKENHI grant numbers 21390126 and 25305007) and a Grant-in-Aid for Challenging Exploratory Research (JSPS KAKENHI grant number 24659460) to S.Y.; and by the UK Medical Research Council (award number MR/N00227X/1) to A.M.B. | The authors have read the journal's policy and declare the following conflicts of interest: S.Y. and A.H. are named inventors on filed patents related to immunization with the B.D.E.S. (WO/2007/091624) and ChAd63 (WO/2008/122769) anti-malaria vaccines, respectively. Neither of these products has been commercialized. None of the authors have undertaken any consultancies relevant to this study. These conflicts of interest do not alter the authors' adherence to all the policies of Scientific Reports on sharing data and materials, as detailed online in the guide for authors. | N/A | N/A | 21/12/2020 |

| Award number | Funder name | Awardee (to whom was the grant?) | Date | Amount | Exchange rate | Amount in GBP | Relevant publications (author and date) | Direct citation from articles |
|---|---|---|---|---|---|---|---|---|
| - | AIDS Vaccine Initiative | - | - | - | | | Borthwick et al (2014) | The work was supported by |
| - | Austrian Federal Ministry of Science and | - | - | - | | | Tiono (2018) | This work was supported by a |
| BBS/E/I/00001373 | BBRSC | Bryan Charleston | Jan 09 - Jan 13 | £790,209 | | £ 790,209.00 | Dicks et al (2015 - 2) | This work has been funded by |
| BB/H010556/1 | BBSRC | Tim Bull | Mar 10 - May 13 | £351,371 | | £ 351,371.00 | Bull et al (2014) | This work was supported by B |
| BB/H010718/1 | BBSRC | Jayne Hope | Sep 11 - Aug 13 | £235,928 | | £ 235,928.00 | Bull et al (2014) | This work was supported by B |
| - | BBSRC | Hope J | - | - | | | Bull et al (2014) | This work was supported by B |
| - | BBSRC | McGuines I | - | - | | | Bull et al (2014) | This work was supported by B |
| LDAD_P15820 | BBSRC | - | - | - | | | Goodman et al (2011) | This work was supported prim |
| LDAD_P15820 | BBSRC | - | - | - | | | Goodman et al (2011) | This work was supported prim |
| OPP1096893 | Bill & Melinda Gates Foundation | Warimwe GM | - | - | | | Dulal et al (2016) | This study was conducted witl |
| OPP1096893 | Bill & Melinda Gates Foundation | Warimwe GM | - | - | | | Warimwe et al (2016) | This work was conducted with |
| OPP1078791 | Bill and Melinda Gate Foundation | Research Institute | Oct-13 | $ 10,999,924 | 0.72 | £ 7,919,945.28 | Svitek (2018) | This work was funded by the I |
| OPP1215550 | Bill and Melinda Gates Foundation | The Pirbright Institute | Nov-19 | $ 5,530,900 | 0.72 | £ 3,982,248.00 | S Graham (2020) | and the Bill and Melinda Gate |
| 3436 | Bill and Melinda Gates Foundation | - | - | - | | | Pearson et al (2013) | This work has been supported |
| - | Biotechnology and Biological Sciences R | - | - | - | | | Boyd et al (2013) | The Biotechnology and Biolog |
| - | Bundesministerium für Bildung und Fors | - | - | - | | | Mensah (2017) | This work was supported by a |
| - | Cancer Research | Malinauskas T | - | - | | | Bauza et al (2014) | The work was funded by a We |
| - | Cancer Research | Jones EY | - | - | | | Bauza et al (2014) | The work was funded by a We |
| - | CAPES | Atcheson E | | - | | | Atcheson (2018) | The work was funded by a We |
| CRP 3.7 | CGIAR Research Program on Livestock a | - | - | - | | | Svitek (2018) | This work was funded by the I |
| - | Coalition for Epidemic Preparedness Inn | - | - | - | | | Folegatti P.M.(2020) | UK Department of Health and |
| finance code 001 | Coordenacao de Aperfeicoamento de Pe | Folegatti P.M | - | - | | | Folegatti P.M.(2020) | UK Department of Health and |
| finance code 001 | Coordenacao de Aperfeicoamento de Pe | - | - | - | | | Folegatti P.M.(2020) | UK Department of Health and |
| - | Dakar University Cheikh Anta Diop | - | - | - | | | Mensah et al (2016) | This study was supported by a |
| - | Danida | - | - | - | | | Payne et al (2017) | This work was supported by fu |
| | Department for Business, Energy and Industrial Strategy | | | £65,500,000.00 | | £ 65,500,000.00 | https://www.imperial.ac.uk/news/197573/covid-19-vacc | |
| | Department for Business, Energy and Industrial Strategy | | | £20,000,000.00 | | £ 20,000,000.00 | https://www.imperial.ac.uk/news/197017/imperial-covic | |
| - | Department for International Developme | - | - | - | | | Bowyer (2018) | The clinical trial was supporte |
| - | Department for International Developme | - | - | - | | | Venkatraman N (2019) | This work was supported |
| - | Department of Biotechnology, Governm | Chauhan VS | - | - | | | de Cassan et al (2011) | S.C.d.C. is a Ph.D. student sup |
| - | Department of Biotechnology, Governm | Chitnis CE | - | - | | | de Cassan et al (2011) | S.C.d.C. is a Ph.D. student sup |
| - | Department of Foreign Affairs and Trade | - | - | - | | | Mensah (2017) | This work was supported by a |
| - | DFID | - | - | - | | | Tapia et al (2016) | This study was funded by a W |
| - | DFID | - | - | - | | | Ewer et al (2016) | Supported by the Wellcome T |
| - | Division of Intramural Research | - | - | - | | | Forbes et al (2012) | This work was funded by the |
| - | Dr. Saal van Zwanenberg Stichting | van Laarhoven A | - | - | | | Lambe et al (2013) | AvL was funded by a fellowsh |
| IP.2008.31100.00 | EDCTP | - | - | - | | | Kimani et al (2014) | This work was supported by |
| IP.2008.31100.001 | EDCTP | - | - | - | | | Afolabi et al (2016) | Trials Partnership (EDCTP) and |
| IP.2008.31100.001 | EDCTP | - | - | - | | | Mensah et al (2016) | This study was supported by a |
| IP.2008.31100.001 | EDCTP | - | - | - | | | Nébié et al (2014) | This work was supported by a |
| - | EMVDA | Dicks MDJ | - | - | | | Dicks et al (2012) | This work has been funded by |
| - | EMVDA | Dicks MDJ | - | - | | | Dicks et al (2015 - 2) | This work has been funded by |
| - | EMVDA | - | - | - | | | Forbes et al (2012) | This work was funded by the |
| LSHP-CT-2007-03750 | EMVDA | - | - | - | | | Biswas et al (2014) | This work was supported by tl |
| LSHP-CT-2007-037506 | EMVDA | - | - | - | | | Sheehy et al (2011) | This work was supported by tl |
| LSHP-CT-2007-037506 | EMVDA | - | - | - | | | Sheehy et al (2012 - 2) | This work was supported by tl |
| LSHP-CT-2007-037506 | EMVDA | - | - | - | | | Sheehy et al (2012) | This work was supported by |
| LSHP-CT-<br>2007–037506 | EMVDA | - | - | - | | | Hodgson et al (2014) | This work was supported by tl |
| | EMVDA | | - | - | | | Draper et al (2011) | SB was funded by MalParTrai |

| | | | | | | | | | |
|---|---|---|---|---|---|---|---|---|---|
| EP/R013756/1 | Engineering and Physical Sciences Resea | Tarit K Mukhopadhyay | Apr 18 - Sep 21 | £6,968,179 | | £ | 6,968,179.00 | Folegatti P.M.(2020) | UK Department of Health and |
| CFTD07/117 | Enterprise Ireland | - | - | - | | | | Pearson et al (2015) | This study was funded by Ente |
| CFTD07/117 | Enterprise Ireland | - | - | - | | | | Carey et al (2013) | This work wassupported by Er |
| - | EU HEPACIVAC | - | - | - | | | | Swadling (2016) | Supported by the Medical Res |
| - | European and Developing Countries Clini | - | - | - | | | | Bowyer G (2020) | The Oxford clinical trial was s |
| - | European and Developing Countries Clini | - | - | - | | | | Reyes-Sandoval (2010) | Work in the Oxford malaria va |
| - | European and Developing Countries Clini | - | - | - | | | | Bowyer (2018) | The clinical trial was supporte |
| - | European and Developing Countries Clini | - | - | - | | | | Venkatraman N (2019) | This work was supported |
| IP.2008.31100.001 | European and Developing Countries Clini | - | - | - | | | | Ogwang et al (2015) | This work was funded by the I |
| SP.2011.41304.025 | European and Developing Countries Clini | - | - | - | | | | Mensah (2017) | This work was supported by a |
| IP.2008.31100.001 | European and Developing Countries Clini | - | - | - | | | | Tiono (2018) | This work was supported by a |
| 666085 | European Comission | GLAXOSMITHKLINE BIOLOGICALS SA | 07-Oct-14 | € 15,153,216 | 0.87 | £ | 13,183,297.92 | Venkatraman N (2019) | This work was supported |
| 666085 | European Commission | GLAXOSMITHKLINE BIOLOGICALS SA | 07-Oct-14 | € 15,153,216 | 0.87 | £ | 13,183,297.92 | Bowyer G (2020) | The Oxford clinical trial was s |
| 242095 | European Commission | Salman AM | 01-Oct-09 | € 12,000,000 | 0.87 | £ | 10,440,000.00 | Atcheson (2018) | The work was funded by a We |
| 727393-PALE-Blu | European Commission | THE UNIVERSITY OF NOTTINGHAM | 01-Jun-17 | € 6,039,301.50 | 0.87 | £ | 5,254,192.31 | Utrilla-Trigo S (2020) | This work was supported by g |
| 602705 | European Commission | - | - | € 6,000,000 | 0.87 | £ | 5,220,000.00 | Cappuccini F (2020) | The VANCE clinical trial was s |
| 602705 | European Commission | THE CHANCELLOR, MASTERS AND SCHOLARS OF THE UNIVERSITY OF OXFORD | 01-Apr-14 | € 6,000,000 | 0.87 | £ | 5,220,000.00 | Tuthill M. (2020) | European Union Seventh Fram |
| 316655 | European Commission | Halbroth BR | 01-Nov-12 | € 3,060,467.67 | 0.87 | £ | 2,662,606.87 | Halbroth (2018) | BRH received funding from th |
| 316655 | European Commission | Halbroth BR | - | - | | | | Gola (2018) | A.G. is funded by the Wellcom |
| - | European Commission | - | - | - | | | | Reyes-Sandoval (2010) | Work in the Oxford malaria va |
| IC18-CT95-0019 | European Commission | Fellowship to author to J.S. | - | - | | | | Gilbert et al (2002) | Wellcome Trust and the Eu |
| 212414 | European Community's 7th Framework F | UNIVERSIDAD COMPLUTENS | 01-Oct-08 | € 2,894,759 | 0.87 | £ | 2,518,440.33 | Pérez et al (2013) | The study was funded by th |
| 242095 | European Community's Seventh Framew | Janse CJ | - | - | | | | Longley et al (2015) | This work has been funded by |
| 242095 | European Community's Seventh Framew | - | - | € 12,000,000 | 0.87 | £ | 10,440,000.00 | Forbes et al (2012) | This work was funded by the \ |
| - | European Malaria Vaccine Development | - | - | - | | | | Draper et al (2010) | This work was funded by the \ |
| LSHP-CT-2007-037506 | European Malaria Vaccine Development | - | - | - | | | | Elias et al (2013) | This work was supported by tl |
| LSHP-CT-2007-037506 | European Malaria Vaccine Development | de Cassan SC | - | - | | | | Payne (2017) | This work was supported by a |
| LSHP-CT-2007-037506 | European Malaria Vaccine Development | de Cassan SC | - | - | | | | de Cassan et al (2011) | S.C.d.C. is a Ph.D. student sup |
| - | European Malaria Vaccine Devlopment A | - | - | - | | | | Capone et al (2010) | This work was supported by tl |
| - | European Malaria Vaccine Initiative | - | - | - | | | | Reyes-Sandoval (2010) | Work in the Oxford malaria va |
| - | European Research Council | Walker A | - | - | | | | Walker et al (2015) | Andrew S. Walker, José Loure |
| - | European Research Council | Lourenço J | - | - | | | | Walker et al (2015) | Andrew S. Walker, José Loure |
| - | European Research Council | Gupta S | - | - | | | | Walker et al (2015) | Andrew S. Walker, José Loure |
| 305282 | European Union | MultiMalVax | 01-Oct-12 | € 6,000,000 | 0.87 | £ | 5,220,000.00 | Payne et al (2017) | This work was supported by fu |
| 316655 | European Union | Halbroth BR | - | - | | | | Longley et al (2017) | Funding for manufacture and |
| 602705 | European Union | Capuccini F | - | - | | | | Capuccini et al (2017) | This work was supported by C |
| 602705 | European Union | Pollock E | - | - | | | | Capuccini et al (2017) | This work was supported by C |
| Framework VI;HEPACI\ | European union | - | - | - | | | | Barnes et al (2012) | European Union (Framework \ |
| 602705 | European Union's Seventh Framework Pı | Capuccini F | 01-Apr-14 | € 6,000,000 | 0.87 | £ | 5,220,000.00 | Capuccini et al (2016) | This work was supported by C |
| 602705 | European Union's Seventh Framework Pı | Pollock E | | - | | | | Capuccini et al (2016) | This work was supported by C |
| - | European Vaccine Initiative | - | - | - | | | | Cottingham et al (2012) | This work was supported by tl |
| - | European Vaccine Initiative | - | - | - | | | | de Barra et al (2014) | The study was funded by a gra |
| - | European Vaccine Initiative | Chauhan VS | - | - | | | | de Cassan et al (2011) | S.C.d.C. is a Ph.D. student sup |
| - | European Vaccine Initiative | Chitnis CE | - | - | | | | de Cassan et al (2011) | S.C.d.C. is a Ph.D. student sup |
| - | European Vaccine Initiative | Viebig NK | - | - | | | | Kimani et al (2014) | This work was supported by |
| 242095 | EVIMalaR | - | - | - | | | | Biswas et al (2014) | This work was supported by tl |

| | | | | | | | | |
|---|---|---|---|---|---|---|---|---|
| 242095 | EVIMalaR | - | - | - | | | Elias et al (2013) | This work was supported by tl |
| 242095 | EVIMalaR | - | - | - | | | Hodgson et al (2014) | This work was supported by tl |
| 242095 | EVIMalaR | Salman M | - | - | | | Salman et al (2017) | The work was funded by a We |
| 242095 | EVIMalaR | - | - | - | | | Sheehy et al (2012) | This work was supported by |
| - | EVIMalR | Salman AM | - | - | | | Longley et al (2015) | This work has been funded by |
| - | Foundation to NIH | Hill A.V.S | - | - | | | Halbroth (2018) | BRH received funding from th |
| - | Frederick National Laboratory for Cancer | - | - | - | | | Tapia et al (2016) | This study was funded by a W |
| - | Gates Foundation | - | - | - | | | Reyes-Sandoval (2010) | Work in the Oxford malaria va |
| - | Gates Foundation (through the foundati | - | - | - | | | Cottingham et al (2012) | This work was supported by tl |
| - | German Center for Infection Research | - | - | - | | | Folegatti P.M.(2020) | UK Department of Health and |
| - | Graduate Women in Science | Coughlan L | - | - | | | Bliss (2020) | This research project was sup |
| - | Grand Challenges in Global Health | - | - | - | | | Reyes-Sandoval et al (2010 | We thank the Jenner Institu |
| 24659460 | Grant-in-Aid for Challenging Exploratory | Yoshida S | - | - | | | Yoshida Klyori (2018) | This work was supported, in p |
| 26860278 | Grant-in-Aid for Young Scientists | - | - | - | | | Yoshida Klyori (2018) | This work was supported, in p |
| 21390126 | Grants-in-Aid for Scientific Research | - | - | - | | | Yoshida Klyori (2018) | This work was supported, in p |
| 25305007 | Grants-in-Aid for Scientific Research | - | - | - | | | Yoshida Klyori (2018) | This work was supported, in p |
| - | GSK | - | | - | | | Venkatraman N (2019) | This work was supported |
| - | HAV Vaccines Ltd | - | - | - | | | Folegatti P.M. (2019) | This research was funded by H |
| - | HEPACIVAC | - | - | - | | | Swadling et al (2014) | Funding:Supported by the Me |
| LSH-2005-1.2.4-2 proje | Hepacivac | - | - | - | | | Colloca et al (2012) | This work was supported in pa |
| - | Imperial College NIHR Wellcome Trust C | - | - | - | | | Rampling et al (2016) | This work was supported by tl |
| 971510 | Innovate UK | The Jenner Institute, Universi | Apr 17 - May 18 | £483,455 | £ | 483,455.00 | Sebastian S (2020) | This research was funded by I |
| FPI-SGIT-201 | Instituto Nacional de Investigación y Tec | Utrilla-Trigo S | - | - | | | Utrilla-Trigo S (2020) | This work was supported by g |
| - | Irish Aid | - | - | - | | | Afolabi et al (2016) | Trials Partnership (EDCTP) and |
| - | Irish Aid | - | - | - | | | Mensah (2017) | This work was supported by a |
| - | Irish Aid | - | - | - | | | Mensah et al (2016) | This study was supported by a |
| - | Irish Aid | - | - | - | | | Tiono (2018) | This work was supported by a |
| - | James Martin School for 21st Century, O | - | - | - | | | Barnes et al (2012) | European Union (Framework ' |
| - | Jenner Institue | Gilbert SC | - | - | | | de Cassan et al (2011) | S.C.d.C. is a Ph.D. student sup |
| - | Jenner Institue | Hill AVS | - | - | | | Dicks et al (2012) | This work has been funded by |
| - | Jenner Institue | Hill AVS | - | - | | | Sheehy et al (2012) | This work was supported by |
| - | Jenner Institue | Gilbert SC | - | - | | | Sheehy et al (2012) | This work was supported by |
| - | Jenner Institute | Reyes-Sandoval A | - | - | | | Atcheson (2018) | The work was funded by a We |
| - | Jenner Institute | Hill A.V.S | - | - | | | Atcheson (2018) | The work was funded by a We |
| - | Jenner Institute | Hill AVS | - | - | | | Bauza et al (2014) | The work was funded by a We |
| - | Jenner Institute | Reyes-Sandoval A | - | - | | | Bauza et al (2014) | The work was funded by a We |
| - | Jenner Institute | McShane H | - | - | | | Betts et al (2012) | Funding was provided by NEW |
| - | Jenner Institute | Hill AVS | - | - | | | Betts et al (2012) | Funding was provided by NEW |
| - | Jenner Institute | Reyes-Sandoval A | - | - | | | Betts et al (2012) | Funding was provided by NEW |
| - | Jenner Institute | Hill AVS | - | - | | | Biswas et al (2014) | This work was supported by tl |
| - | Jenner Institute | Draper SJ | - | - | | | Biswas et al (2014) | This work was supported by tl |
| - | Jenner Institute | Hill AVS | - | - | | | Carey et al (2013) | This work wassupported by Er |
| - | Jenner Institute | Gilbert SC | - | - | | | Carey et al (2013) | This work wassupported by Er |
| - | Jenner Institute | Hill AVS | - | - | | | de Barra et al (2014) | The study was funded by a gra |
| - | Jenner Institute | Gilbert SC | - | - | | | de Barra et al (2014) | The study was funded by a gra |
| - | Jenner Institute | Hill AVS | - | - | | | de Cassan et al (2011) | S.C.d.C. is a Ph.D. student sup |
| - | Jenner Institute | Draper SJ | - | - | | | de Cassan et al (2011) | S.C.d.C. is a Ph.D. student sup |
| - | Jenner Institute | Gilbert SC | - | - | | | Dicks et al (2012) | This work has been funded by |
| - | Jenner Institute | Hill AVS | - | - | | | Dicks et al (2015) | This work has been funded by |
| - | Jenner Institute | Gilbert SC | - | - | | | Dicks et al (2015) | This work has been funded by |
| - | Jenner Institute | Gilbert SC | - | - | | | Draper et al (2010) | This work was funded by the ' |
| - | Jenner Institute | Hill A.V.S. | - | - | | | Draper et al (2010) | This work was funded by the ' |
| - | Jenner Institute | Hill AVS | - | - | | | Dulal et al (2016) | This study was conducted witl |
| - | Jenner Institute | Charleston B | - | - | | | Dulal et al (2016) | This study was conducted witl |

| | | | | | | |
|---|---|---|---|---|---|---|
| - | Jenner Institute | Hill AVS | - | - | Elias et al (2013) | This work was supported by th |
| - | Jenner Institute | Draper SJ | - | - | Elias et al (2013) | This work was supported by th |
| - | Jenner Institute | Hill AVS | - | - | Ewer et al (2013) | The study was funded by gran |
| - | Jenner Institute | Draper SJ | - | - | Ewer et al (2013) | The study was funded by gran |
| - | Jenner Institute | Gilbert SC | - | - | Ewer et al (2013) | The study was funded by gran |
| - | Jenner Institute | Reyes-Sandoval A | - | - | Ewer et al (2013) | The study was funded by gran |
| - | Jenner Institute | Hill AVS | - | - | Forbes et al (2012) | This work was funded by the W |
| - | Jenner Institute | Draper SJ | - | - | Forbes et al (2012) | This work was funded by the W |
| - | Jenner Institute | Draper SJ | - | - | Goodman et al (2011) | This work was supported prim |
| - | Jenner Institute | Hill A.V.S | - | - | Goodman et al (2011) | This work was supported prim |
| - | Jenner Institute | Barnes E | - | - | Kelly et al (2016) | Supported by a Medical Resea |
| - | Jenner Institute | Gilbert SC | - | - | Lambe et al (2013) | AvL was funded by a fellowsh |
| - | Jenner Institute | Hill AVS | - | - | Longley et al (2015) | This work has been funded by |
| - | Jenner Institute | Hill AVS | - | - | Longley et al (2017) | Funding for manufacture and |
| - | Jenner Institute | Hill A.V.S | - | - | Payne (2017) | This work was supported by a |
| - | Jenner Institute | Draper SJ | - | - | Payne (2017) | This work was supported by a |
| - | Jenner Institute | Hill AVS | - | - | Payne et al (2017) | This work was supported by fu |
| - | Jenner Institute | Draper SJ | - | - | Payne et al (2017) | This work was supported by fu |
| - | Jenner Institute | Biswas S | - | - | Payne et al (2017) | This work was supported by fu |
| - | Jenner Institute | Reyes-Sandoval A | - | - | Salman et al (2017) | The work was funded by a We |
| - | Jenner Institute | Hill AVS | - | - | Salman et al (2017) | The work was funded by a We |
| - | Jenner Institute | Gilbert SC | - | - | Sheehy et al (2011) | This work was supported by th |
| - | Jenner Institute | Hill AVS | - | - | Sheehy et al (2011) | This work was supported by th |
| - | Jenner Institute | Draper SJ | - | - | Sheehy et al (2011) | This work was supported by th |
| - | Jenner Institute | Gilbert SC | - | - | Sheehy et al (2012 - 2) | This work was supported by th |
| - | Jenner Institute | Draper SJ | - | - | Sheehy et al (2012 - 2) | This work was supported by th |
| - | Jenner Institute | Draper SJ | - | - | Sheehy et al (2012) | This work was supported by |
| - | Jenner Institute | Gilbert S.C. | - | - | Stedman (2019) | This study was funded by the |
| - | Jenner Institute | Charleston B | - | - | Stedman (2019) | This study was funded by the |
| - | Jenner Institute | A.V.S Hill | - | - | Stedman (2019) | This study was funded by the |
| - | Jenner Institute | Barnes E | - | - | Swadling (2016) | Supported by the Medical Res |
| - | Jenner Institute | Hill AVS | - | - | Walker et al (2015) | Andrew S. Walker, José Loure |
| - | Jenner Institute | Hill A.V.S | - | - | Wang (2018) | This work has been supported |
| - | Jenner Institute | Draper AD | - | - | Wang (2018) | This work has been supported |
| - | Jenner Institute | Douglas SJ | - | - | Wang (2018) | This work has been supported |
| - | Jenner Institute | Hill AVS | - | - | Warimwe et al (2013) | This work was supported by a |
| - | Jenner Institute | Gilbert SC | - | - | Warimwe et al (2013) | This work was supported by a |
| - | Jenner Institute | Hill AVS | - | - | Warimwe et al (2016) | This work was conducted with |
| - | Jenner Institute | Gilbert SC | - | - | Warimwe et al (2016) | This work was conducted with |
| - | Jenner Institute | Charleston B | - | - | Warimwe et al (2016) | This work was conducted with |
| - | Jenner Institute | Hill AVS | - | - | Sheehy et al (2012 - 2) | This work was supported by th |
| - | Jenner Institute | Gilbert S.C. | - | - | Alharbi (2019) | This study is funded by KAIMR |
| - | Jenner Institute | Douglas AD | - | - | Fedosyuk S (2019) | This work was supported by M |
| - | Jenner Institute | Rollier C | - | - | Rollier C (2020) | This work was funded by a gra |
| - | Jenner Institute | Hill A.V.S. | - | - | Rollier C (2020) | This work was funded by a gra |
| - | Jenner Institute | Gilbert S.C. | - | - | Rollier C (2020) | This work was funded by a gra |
| - | Jenner Institute | Gilbert S.C. | - | - | van Doremalen N (2019) | This work was supported by th |
| - | Jenner Vaccine Foundation | - | - | - | Reyes-Sandoval (2010) | Work in the Oxford malaria va |
| RC16/093 | KAIMRC | Naif Khalaf Alharbi | - | - | Alharbi (2019) | This study is funded by KAIMR |
| HI15C2971 | Korean Ministry of Health and Welfare | - | - | - | Folegatti P.M.(2020) | UK Department of Health and |
| - | Lister Institute | Draper SJ | - | - | Biswas et al (2014) | This work was supported by th |
| - | Lister Institute | Draper SJ | - | - | Elias et al (2013) | This work was supported by th |
| - | Lister Institute | Draper SJ | - | - | Payne (2017) | This work was supported by a |
| - | Lister Institute | Draper SJ | - | - | Payne et al (2017) | This work was supported by fu |
| - | Lister Institute | Draper SJ | - | - | Carey et al (2013) | This work wassupported by Er |

| | | | | | | | | | |
|---|---|---|---|---|---|---|---|---|---|
| - | Lister Institute | Douglas SJ | - | - | | | | Wang (2018) | This work has been supported |
| - | Malaria Vaccine Initiative | - | - | - | | | | de Barra et al (2014) | The study was funded by a gra |
| - | Malaria Vectored Vaccines Consortium ( | - | - | - | | | | Ogwang et al (2015) | This work was funded by the I |
| MEST-CT-2005-020492 | MalParTraining | Biswas S | - | - | | | | Draper et al (2011) | SB was funded by MalParTrai |
| G0600424 | Medical Research Council | Anna Louise Goodman | Sep 06 - Oct 09 | | £159,968 | £ | 159,968.00 | Goodman et al (2011) | This work was supported prim |
| - | Meningitis UK | Rollier CS | - | - | | | | Reyes-Sandoval et al (2010 | We thank the Jenner Institu |
| - | Merck KGaA | - | - | - | | | | Fedosyuk S (2019) | This work was supported by M |
| - | Merton College, Oxford | Draper SJ | - | - | | | | Draper et al (2010) | This work was funded by the V |
| - | Merton College, Oxford | Draper SJ | - | - | | | | Draper et al (2011) | SB was funded by MalParTrai |
| - | MEWA, Saudi Arabia | - | - | - | | | | Alharbi (2019) | This study is funded by KAIMR |
| - | MRC | - | - | - | | - | | Bowyer (2018) | The clinical trial was supporte |
| - | MRC | - | - | - | | - | | Coughlan L (2018) | Medical Research Council UK, |
| - | MRC | Juthathip Mongkolsapaya | - | - | | - | | Lopez-Camacho (2018) | This report is independent res |
| - | MRC | - | - | - | | - | | Mensah (2017) | This work was supported by a |
| - | MRC | Warimwe GM | - | - | | - | | Munster (2017) | This work is published withthe |
| - | MRC | - | - | - | | - | | Venkatraman N (2019) | This work was supported |
| MC_UU_12014 | MRC | Patel AH, Kohl A | - | - | | - | | Lopez-Camacho (2018) | This report is independent res |
| MR/P017339/1 | MRC | Alexander Donald Douglas | Apr 17 - Mar 22 | | £2,228,194 | £ | 2,228,194.00 | Fedosyuk S (2019) | This work was supported by M |
| MR/P017339/1 | MRC | Alexander Donald Douglas | Apr 17 - Mar 22 | | £2,228,194 | £ | 2,228,194.00 | Wang (2018) | This work has been supported |
| MR/N019008/1 | MRC | Reyes-Sandoval A | Aug 16 - Feb 22 | | £1,792,688 | £ | 1,792,688.00 | Atcheson (2018) | The work was funded by a We |
| MR/N019008/1 | MRC | Reyes-Sandoval A | Aug 16 - Feb 22 | | £1,792,688 | £ | 1,792,688.00 | Salman et al (2017) | The work was funded by a We |
| G0701669 | MRC | Tomas Hanke | Oct 08 - Oct 12 | | £1,300,519 | £ | 1,300,519.00 | Borthwick et al (2014) | The work was supported by |
| MR/L009528/1 | MRC | Thomas Alexander Bowden | Jan 14 - Dec 18 | | £1,144,287 | £ | 1,144,287.00 | van Doremalen N (2019) | This work was supported by th |
| G1000527 | MRC | Draper SJ | Aug 10 - Jul 15 | | £1,104,645 | £ | 1,104,645.00 | Biswas et al (2014) | This work was supported by th |
| G1000527 | MRC | Draper SJ | Aug 10 - Jul 15 | | £1,104,645 | £ | 1,104,645.00 | Carey et al (2013) | This work wassupported by En |
| G1000527 | MRC | Draper SJ | Aug 10 - Jul 15 | | £1,104,645 | £ | 1,104,645.00 | Elias et al (2013) | This work was supported by th |
| G1000527 | MRC | Draper SJ | Aug 10 - Jul 15 | | £1,104,645 | £ | 1,104,645.00 | Sheehy et al (2012 - 2) | This work was supported by th |
| G1100086 | MRC | Simon Draper | Jul 11 - Jun 14 | | £895,438 | £ | 895,438.00 | Payne (2017) | This work was supported by a |
| G0700735 | MRC | Adrian Hill | Jan 08 - Dec 10 | | £748,840 | £ | 748,840.00 | Biswas et al (2014) | This work was supported by th |
| G0700735 | MRC | Adrian Hill | Jan 08 - Dec 10 | | £748,840 | £ | 748,840.00 | Elias et al (2013) | This work was supported by th |
| MR/N006372/1 | MRC | Sarah Catherine Gilbert | Mar 16 - May 18 | | £679,559 | £ | 679,559.00 | Asthagiri Arunkumar (2019 | The study was fundedby an M |
| MC_PC_13073 | MRC | Chas Bountra | Mar 14 - Sep 15 | | £650,000 | £ | 650,000.00 | Wang (2018) | This work has been supported |
| G0502018 | MRC | Adrian Hill | Oct 06 - Jan 09 | | £647,586 | £ | 647,586.00 | O'Hara et al (2012) | Financial support. This work w |
| MR/N00227X/1 | MRC | Andrew Michael Blagborough | Jan 16 - Apr 19 | | £549,297 | £ | 549,297.00 | Yoshida Klyori (2018) | This work was supported, in p |
| MC_PC_15040 | MRC | Stephen Ward | Mar 16 - Feb 18 | | £500,000 | £ | 500,000.00 | Sebastian S (2020) | This research was funded by I |
| G0701694 | MRC | Eleanor Barnes | Aug 09 - Jul 12 | | £250,000 | £ | 250,000.00 | Kelly et al (2016) | Supported by a Medical Resea |
| G0701694 | MRC | Eleanor Barnes | Aug 09 - Jul 12 | | £250,000 | £ | 250,000.00 | Swadling (2016) | Supported by the Medical Res |
| MR/N017552/1 | MRC | Kohl A | Jan 16 - Jan 19 | | £221,947 | £ | 221,947.00 | Lopez-Camacho (2018) | This report is independent res |
| G0600424 | MRC | SJD | - | - | | £ | 159,968.00 | Goodman et al (2011) | This work was supported prim |
| G0600424 | MRC | Goodman A | Sep 06 - Oct 09 | | £159,968 | £ | 159,968.00 | Ewer et al (2013) | The study was funded by gran |
| - | MRC | - | - | - | | | | Afolabi et al (2016) | Trials Partnership (EDCTP) and |
| - | MRC | - | - | - | | | | Antrobus et al (2014) | The study was funded by gran |
| - | MRC | - | - | - | | | | Barnes et al (2012) | European Union (Framework V |
| - | MRC | Jones EY | - | - | | | | Bauza et al (2014) | The work was funded by a We |
| - | MRC | Malinauskas T | - | - | | | | Bauza et al (2014) | The work was funded by a We |
| - | MRC | - | - | - | | | | Bowyer G (2020) | The Oxford clinical trial was s |
| - | MRC | Stribbling S | - | - | | | | Capuccini et al (2016) | This work was supported by C |
| - | MRC | Stribbling S | - | - | | | | Capuccini et al (2017) | This work was supported by C |
| - | MRC | Bartiromo M | - | - | | | | Colloca et al (2012) | This work was supported in pa |
| - | MRC | - | - | - | | | | Ewer et al (2013) | The study was funded by gran |
| - | MRC | Draper SJ | - | - | | | | Forbes et al (2012) | This work was funded by the V |

| | | | | | | | | |
|---|---|---|---|---|---|---|---|---|
| - | MRC | Draper SJ | - | - | | | Goodman et al (2011) | This work was supported prim |
| - | MRC | Goodman AL | - | - | | | Goodman et al (2011) | This work was supported prim |
| - | MRC | Barnes E | - | - | | | Kelly et al (2016) | Supported by a Medical Resea |
| - | MRC | - | - | - | | | Kimani et al (2014) | This work was supported by |
| - | MRC | - | - | - | | | Mensah et al (2016) | This study was supported by a |
| - | MRC | - | - | - | | | Ogwang et al (2013) | This work was performed by t |
| - | MRC | - | - | - | | | Swadling (2016) | Supported by the Medical Res |
| - | MRC | Swadling L | - | - | | | Swadling (2016) | Supported by the Medical Res |
| - | MRC | Barnes E | - | - | | | Swadling (2016) | Supported by the Medical Res |
| - | MRC | - | - | - | | | Swadling et al (2014) | Funding:Supported by the Me |
| - | MRC | Barnes E | - | - | | | Swadling et al (2014) | Funding:Supported by the Me |
| - | MRC | Swadling L | - | - | | | Swadling et al (2014) | Funding:Supported by the Me |
| - | MRC | - | - | - | | | Tapia et al (2016) | This study was funded by a W |
| - | MRC | - | - | - | | | Ewer et al (2016) | Supported by the Wellcome T |
| - | MRC | Swadling L | - | - | | | Kelly et al (2016) | Supported by a Medical Resea |
| G0600311 | MRC | - | - | - | | | Pearson et al (2013) | This work has been supported |
| G0600311 | MRC | - | - | - | | | Pearson et al (2015) | This study was funded by Ente |
| G0600424 | MRC | ALG | Sep 06 - Oct 09 | £159,968 | | | Goodman et al (2011) | This work was supported prim |
| G0700735 | MRC | - | - | - | | | Sheehy et al (2011) | This work was supported by th |
| G1000157 | MRC | Draper SJ | - | - | | | Sheehy et al (2011) | This work was supported by th |
| G1000527 | MRC | Draper SJ | - | - | | | de Cassan et al (2011) | S.C.d.C. is a Ph.D. student sup |
| MR/K017632/1) | MRC | Alanine DGW | - | - | | | Payne et al (2017) | This work was supported by fu |
| U117532067 | MRC | Holder AA | - | - | | | Draper et al (2011) | SB was funded by MalParTrai |
| - | MRC and DFID | - | - | - | | | Afolabi et al (2016) | Trials Partnership (EDCTP) an |
| - | MRC and DFID | - | - | - | | | Afolabi et al (2016) | Trials Partnership (EDCTP) an |
| - | MRC and DFID | Bejon P | - | - | | | Ogwang et al (2015) | This work was funded by the |
| - | National Cancer Institute | - | - | - | | | Tapia et al (2016) | This study was funded by a W |
| P51 RR000167 | National Center for Research Resources | - | - | - | | | Rollier C (2020) | This work was funded by a gra |
| RR020141 | National Center for Research Resources | - | - | - | | | Rollier C (2020) | This work was funded by a gra |
| RR15459 | National Center for Research Resources | - | - | - | | | Rollier C (2020) | This work was funded by a gra |
| P51 RR000167 | National Centre for Research Resources | - | - | - | | | Spencer et al (2014) | This work has been funded by |
| RR020141 | National Centre for Research Resources | - | - | - | | | Spencer et al (2014) | This work has been funded by |
| RR15459 | National Centre for Research Resources | - | - | - | | | Spencer et al (2014) | This work has been funded by |
| - | National Health Service Blood and Trans | - | - | - | | | Ewer et al (2016) | Supported by the Wellcome T |
| - | National Insitute of Health and Research | - | - | - | | | Tiono (2018) | This work was supported by a |
| - | National Institue of Allergy and Infectiou | - | - | - | | | Sheehy et al (2012 - 2) | This work was supported by th |
| - | National Institute for Health and Resear | - | - | - | | | Barnes et al (2012) | European Union (Framework |
| - | National Institute for Health Research | - | - | - | | | Bowyer G (2020) | The Oxford clinical trial was s |
| - | National Institute for Health Research | - | - | - | | | de Cassan et al (2011) | S.C.d.C. is a Ph.D. student sup |
| - | National Institute for Health Research | - | - | - | | | Folegatti P.M.(2020) | UK Department of Health and |
| - | National Institute for Health Research | Rollier C | - | - | | | Rollier C (2020) | This work was funded by a gra |
| - | National Institute for Health Research | Hill A.V.S. | - | - | | | Rollier C (2020) | This work was funded by a gra |
| - | National Institute for Health Research | | - | - | | | van Doremalen N (2020) | This work was supported by th |
| - | National Institute for Health Research | | - | - | | | van Doremalen N (2020) | This work was supported by th |
| - | National Institute of Allergy and Infectio | - | - | - | | | Sheehy et al (2012) | This work was supported by |
| HHSN272201400008C | National Institute of Allergy and Infectio | Icahn School Of Medicine At | 09/01/2014 | $78,100,000 | 0.72 | £ 56,232,000.00 | S Graham (2020) | Developmentof SARS-CoV-2 |
| - | National Institute of Allergy and Infectio | - | - | - | | | Biswas et al (2014) | This work was supported by th |
| - | National Institute of Allergy and Infectious Diseases | | - | - | | | Sheehy et al (2011) | This work was supported by th |
| - | National Institute of Allergy and Infectio | - | - | - | | | Tapia et al (2016) | This study was funded by a W |
| - | National Institute of Allergy and Infectio | EMMES Corporation | - | - | | | Tapia et al (2016) | This study was funded by a W |
| HHSN261200800001E | National Institute of Allergy and Infectio | Leidos Biomedical Research | - | - | | | Tapia et al (2016) | This study was funded by a W |
| R44AI058375 | National Institute of Allergy and Infectio | - | - | - | | | Longley et al (2017) | Funding for manufacture and |
| | National Institute of Allergy and Infectious Diseases | | - | - | | | Draper et al (2011) | SB was funded by MalParTrai |

| | | | | | | | | | |
|---|---|---|---|---|---|---|---|---|---|
| | National Institute of Allergy and Infectio | - | - | - | | | | Payne et al (2017) | This work was supported by fu |
| | National Institute of Allergy and Infectio | - | - | - | | | | van Doremalen N (2019) | This work was supported by th |
| - | National Institute of Allergy and Infectio | - | - | - | | | | van Doremalen N (2020) | This work was supported by th |
| - | National Institute of Allergy and Infectio | - | - | - | | | | van Doremalen N (2020) | This work was supported by th |
| HHSN272201400008C | National Institute of Allergy and Infectio | - | - | - | | | | van Doremalen N (2020) | This work was supported by th |
| - | National Institute of Health | - | - | - | | | | Biswas et al (2014) | This work was supported by th |
| - | National Institute of Health | - | - | - | | | | Folegatti P.M. (2019) | This research was funded by H |
| - | National Institute of Health | Hill A.V.S | - | - | | | | Gola (2018) | A.G. is funded by the Wellcom |
| - | National Institute of Health | - | - | - | | | | Reyes-Sandoval (2010) | Work in the Oxford malaria va |
| - | National Institute of Health | - | - | - | | | | Rollier C (2020) | This work was funded by a gra |
| - | National Institute of Health | - | - | - | | | | Sheehy et al (2012) | This work was supported by |
| - | National Institute of Health | - | - | - | | | | Sheehy et al (2012 - 2) | This work was supported by th |
| HHSN272201400008C | National Institute of Health | - | - | - | | | | Bliss (2020) | This research project was sup |
| | National Institute of Health | | - | - | | | | Draper et al (2011) | SB was funded by MalParTrai |
| HILL05GCGH0 | National Institute of Health | | - | - | | | | Spencer et al (2014) | This work has been funded by |
| - | National Institute of Health and Researc | - | - | - | | | | Bowyer (2018) | The clinical trial was supporte |
| - | National Institute of Health and Researc | - | - | - | | | | Coughlan L (2018) | Medical Research Council UK, |
| - | National Institute of Health and Researc | - | - | - | | | | Venkatraman N (2019) | This work was supported |
| - | National Institute of Health and Researc | - | - | - | | | | Bliss (2018) | This study was funded by the |
| - | National Institutes of Allergy and Infecti | - | - | - | | | | Forbes et al (2012) | This work was funded by the V |
| 1ZIAAI001179-01 | National Institutes of Health | MUNSTER, VINCENT | 2013 | $877,861 | 0.72 | £ | 632,059.92 | van Doremalen N (2020) | This work was supported by th |
| - | National Institutes of Health | - | - | - | | | | Forbes et al (2012) | This work was funded by the V |
| - | National Institutes of Health | - | - | - | | | | Sheehy et al (2011) | This work was supported by th |
| - | National Institutes of Health, National Ir | - | - | - | | | | Hodgson et al (2014) | This work was supported by th |
| - | NDM | Biswas S | - | - | | | | Biswas et al (2014) | This work was supported by th |
| 26-6. | NEKKEN | Iyori M | 2014 | - | | | | Yoshida Klyori (2018) | This work was supported, in p |
| 27-5. | NEKKEN | Iyori M | 2015 | - | | | | Yoshida Klyori (2018) | This work was supported, in p |
| EC FP7 | NEWTBVAC | - | - | - | | | | Betts et al (2012) | Funding was provided by NEW |
| 1041802 | NHMRC | McCarthy JS | - | - | | | | Payne (2017) | This work was supported by a |
| 10418020 | NHMRC | McCarthey JS | - | - | | | | Payne et al (2017) | This work was supported by fu |
| 5T32AI007647-17 | NIAID | PALESE, PETER | 04-May-16 | $523,884 | 0.72 | £ | 377,196.48 | Asthagiri Arunkumar (2019 | The study was fundedby an M |
| 1R03AI142046-01 | NIAID | ALBRECHT, RANDY A. | 01-Dec-18 | $84,750 | 0.72 | £ | 61,020.00 | McHanon M (2019) | The study was fundedby an M |
| - | NIAID | Gola A | - | - | | | | Gola (2018) | A.G. is funded by the Wellcom |
| - | NIAID | Uderhardt S | - | - | | | | Gola (2018) | A.G. is funded by the Wellcom |
| - | NIAID | Germain RN | - | - | | | | Gola (2018) | A.G. is funded by the Wellcom |
| - | NIAID | - | - | - | | | | Munster (2017) | This work is published withthe |
| AI109946 | NIAID | - | - | - | | | | McHanon M (2019) | The study was fundedby an M |
| HHSN272201400008C | NIAID | - | - | - | | | | Asthagiri Arunkumar (2019 | The study was fundedby an M |
| HHSN272201400008C | NIAID | - | - | - | | | | McHanon M (2019) | The study was fundedby an M |
| 1U19AI082630-01 | NIH | CHUNG, RAYMOND T | 07-Jun-09 | $3,086,377 | 0.72 | £ | 2,222,191.44 | Barnes et al (2012) | European Union (Framework V |
| 2U19AI082630-06 | NIH | CHUNG, RAYMOND T | 30-May-14 | $2,351,111 | 0.72 | £ | 1,692,799.92 | Kelly et al (2016) | Supported by a Medical Resea |
| - | NIH | - | - | - | | | | Munster (2017) | This work is published withthe |
| 2U19AI082630-06 | NIH | Klenerman P | - | - | | | | Kelly et al (2016) | Supported by a Medical Resea |
| 2U19AI082630-06 | NIH | Kelly C | - | - | | | | Swadling (2016) | Supported by the Medical Res |
| 2U19AI082630-06 | NIH | Klenerman P | - | - | | | | Swadling (2016) | Supported by the Medical Res |
| HILL05GCGH0 | NIH | - | - | - | | | | Dicks et al (2015 - 2) | This work has been funded by |
| - | NIH foundation | - | - | - | | | | Dicks et al (2012) | This work has been funded by |
| - | NIHR | - | - | - | | | | Mensah (2017) | This work was supported by a |
| - | NIHR | - | - | - | | | | Payne (2017) | This work was supported by a |
| - | NIHR | Hill AVS | - | - | | | | Walker et al (2015) | Andrew S. Walker, José Loure |
| - | NIHR | Hill AVS | - | - | | | | Ewer et al (2013) | The study was funded by gran |
| - | NIHR | | - | - | | | | Reyes-Sandoval et al (2010 | We thank the Jenner Institu |
| - | NIHR (Oxford Biomedical Research Cent | - | - | - | | | | O'Hara et al (2012) | Financial support. This work w |

| | | | | | | |
|---|---|---|---|---|---|---|
| A91301 Adult Vaccine | NIHR (through Oxford Biomedical Resea | - | - | - | Sheehy et al (2011) | This work was supported by th |
| - | NIHR Oxford Biomedical Research Centr | - | - | - | Antrobus et al (2014) | The study was funded by gran |
| - | NIHR Oxford Biomedical Research Centr | - | - | - | Cottingham et al (2012) | This work was supported by th |
| - | NIHR Oxford Biomedical Research Centr | - | - | - | Elias et al (2013) | This work was supported by th |
| - | NIHR Oxford Biomedical Research Centr | Barnes E | - | - | Swadling et al (2014) | Funding:Supported by the Me |
| - | NIHR Oxford Biomedical Research Centr | - | - | - | Ewer et al (2013) | The study was funded by gran |
| 084113/Z/07/Z | NIHR Oxford Biomedical Research Centr | - | - | - | Biswas et al (2014) | This work was supported by th |
| A91301 Adult Vaccine | NIHR Oxford Biomedical Research Centr | - | - | - | de Barra et al (2014) | The study was funded by a gra |
| A91301 Adult Vaccine | NIHR Oxford Biomedical Research Centr | - | - | - | Kimani et al (2014) | This work was supported by |
| A91301 Adult Vaccine | NIHR Oxford Biomedical Research Centr | - | | - | Ogwang et al (2013) | This work was performed by t |
| A91301 Adult Vaccine | NIHR Oxford Biomedical Research Centr | - | - | - | Sheehy et al (2012 - 2) | This work was supported by th |
| A91301 Adult Vaccine | NIHR Oxford Biomedical Research Centr | - | - | - | Sheehy et al (2012) | This work was supported by |
| A91301 Adult Vaccine | NIHR Oxford Biomedical Research Centr | - | - | - | Hodgson et al (2014) | This work was supported by th |
| A91301,Adult Vaccine | NIHR Oxford Biomedical Research Centr | - | - | - | Hodgson et al (2015) | This work was supported by th |
| - | NIHR Oxford BRC | - | - | - | Capuccini et al (2017) | This work was supported by O |
| - | NIHR Oxford BRC | - | - | - | Ewer et al (2016) | Supported by the Wellcome T |
| - | NIHR Oxford BRC | - | - | - | Longley et al (2017) | Funding for manufacture and |
| - | NIHR Oxford BRC | - | | - | Payne et al (2017) | This work was supported by fu |
| - | NIHR Oxford BRC | - | - | - | Rampling et al (2016) | This work was supported by th |
| - | NIHR Oxford BRC | Kelly C | - | - | Swadling (2016) | Supported by the Medical Res |
| - | NIHR Oxford BRC | Klenerman P | - | - | Swadling (2016) | Supported by the Medical Res |
| - | NIHR Oxford BRC | Barnes E | - | - | Swadling (2016) | Supported by the Medical Res |
| 91301 Adult Vaccine | NIHR Oxford BRC | - | - | - | Afolabi et al (2016) | Trials Partnership (EDCTP) and |
| 58-5348-2-117F | Norman Borlaug Commemorative Resea | - | - | - | Svitek (2018) | This work was funded by the |
| - | Nuffield Department of Medicine | Reyes-Sandoval A | - | - | Reyes-Sandoval et al (2010 | We thank the Jenner Institu |
| - | Nuffield Department of Medicine | Longley RJ | - | - | Longley et al (2015) | This work has been funded by |
| - | Nuffield Department of Medicine | Longley RJ | - | - | Longley et al (2017) | Funding for manufacture and |
| - | Oak Foundation | - | - | - | Stedman (2019) | This study was funded by the |
| 16/107/05 | ODA budget | Reyes-Sandoval A, Patel AH | - | - | Lopez-Camacho (2018) | This report is independent res |
| | Ohyama Health Foundation | - | - | - | Yoshida Klyori (2018) | This work was supported, in p |
| - | Oxford Biomedical Research Centre | - | - | - | de Cassan et al (2011) | S.C.d.C. is a Ph.D. student sup |
| - | Oxford Biomedical Research Centre | Rollier CS | - | - | Reyes-Sandoval et al (2010 | We thank the Jenner Institu |
| - | Oxford Martin Institute | Spencer AJ | - | - | Longley et al (2017) | Funding for manufacture and |
| - | Oxford Martin Institute | Hill AVS | - | - | Salman et al (2017) | The work was funded by a We |
| - | Oxford Martin School | Reyes-Sandoval A | - | - | Atcheson (2018) | The work was funded by a We |
| - | Oxford Martin School | Hill A.V.S | - | - | Atcheson (2018) | The work was funded by a We |
| - | Oxford Martin School | Reyes-Sandoval A | - | - | Bauza et al (2014) | The work was funded by a We |
| - | Oxford Martin School | - | - | - | Cottingham et al (2012) | This work was supported by th |
| - | Oxford Martin School | Barnes E | - | - | Kelly et al (2016) | Supported by a Medical Resea |
| - | Oxford Martin School | Lambe T | - | - | Lambe et al (2013) | AvL was funded by a fellowsh |
| - | Oxford Martin School | Spencer AJ | - | - | Longley et al (2015) | This work has been funded by |
| - | Oxford Martin School | Reyes-Sandoval A | - | - | Salman et al (2017) | The work was funded by a We |
| - | Oxford Martin School | Barnes E | - | - | Swadling (2016) | Supported by the Medical Res |
| - | Oxford Martin School | Hill AVS | - | - | Bauza et al (2014) | The work was funded by a We |
| - | Oxford Martin School | - | - | - | Antrobus et al (2014) | The study was funded by gran |
| - | Oxford Martin School | Spencer AJ | - | - | Dicks et al (2015) | This work has been funded by |
| - | Oxford Martin School | Cottingham MG | - | - | Dicks et al (2015) | This work has been funded by |
| - | Oxford Martin School | Cottingham MG | - | - | Dicks et al (2012) | This work has been funded by |
| - | Oxford Martin Schools | Barnes E | - | - | Swadling et al (2014) | Funding:Supported by the Me |

| | | | | | | | |
|---|---|---|---|---|---|---|---|
| - | Oxford NHIRBRC | - | - | - | | Kelly et al (2016) | Supported by a Medical Resea |
| - | Oxford NHIRBRC | Barnes E | - | - | | Kelly et al (2016) | Supported by a Medical Resea |
| - | Oxford NIHR Biomedical Research Centr | - | - | - | | Barnes et al (2012) | European Union (Framework ' |
| - | Oxford NIHR Biomedical Research Centr | Redchenko I | - | - | | Capuccini et al (2016) | This work was supported by C |
| - | PATH Malaria Vaccine Initiative | - | - | - | | Forbes et al (2012) | This work was funded by the ' |
| - | PATH Malaria Vaccine Initiative | - | - | - | | Hodgson et al (2014) | This work was supported by tl |
| - | PATH Malaria Vaccine Initiative | - | - | - | | Rampling T (2018) | This work was funded primari |
| - | PATH Malaria Vaccine Initiative | - | - | - | | Sheehy et al (2012) | This work was supported by |
| - | PATH Malaria Vaccine Initiative | - | - | - | | Biswas et al (2014) | This work was supported by tl |
| - | PATH Malaria Vaccine Initiative | - | - | - | | Rampling et al (2016) | This work was supported by tl |
| - | PATH Malaria Vaccine Initiative (MVI) | - | - | - | | Sheehy et al (2011) | This work was supported by tl |
| - | PATH MalariaVaccine Initiative | - | - | - | | Hodgson et al (2015) | This work was supported by tl |
| - | PATH MalariaVaccineInitiative | - | - | - | | Sheehy et al (2012 - 2) | This work was supported by tl |
| | PATH-MVI Malaria Vaccine Initiative | - | - | - | | Draper et al (2011) | SB was funded by MalParTrai |
| - | Public Health England | - | - | - | | Ewer et al (2016) | Supported by the Wellcome T |
| - | ReiThera (formerly Okairos) | Nicosia A | - | - | | Mensah et al (2016) | This study was supported by a |
| - | Rhodes Trust | - | - | - | | Longley et al (2015) | This work has been funded by |
| - | Rhodes Trust | Longley RJ | - | - | | Longley et al (2017) | Funding for manufacture and |
| - | Rhodes Trust | Llewellyn D | - | - | | Payne (2017) | This work was supported by a |
| - | Science Foundation Ireland | - | - | - | | Carey et al (2013) | This work wassupported by Er |
| NAP156 | Science Foundation Ireland | - | - | - | | Pearson et al (2015) | This study was funded by Ente |
| NAP170 | Science Foundation Ireland | - | - | - | | Pearson et al (2015) | This study was funded by Ente |
| - | Southampton NIHR Wellcome Trust Clini | - | - | - | | Sheehy et al (2012) | This work was supported by |
| - | Southampton NIHR Wellcome Trust Clini | - | - | - | | Rampling et al (2016) | This work was supported by tl |
| AGL2017-82570-R | Spanish Ministry of Science | - | | - | | Utrilla-Trigo S (2020) | This work was supported by g |
| - | Spanish Ministry of Science | Lopez-Gil E | - | - | | Warimwe et al (2013) | This work was supported by a |
| AGL2011-22485 | Spanish Ministry of Science | Brun AV | - | - | | Warimwe et al (2013) | This work was supported by a |
| - | St Catherine's College, Oxford | Biswas S | - | - | | Biswas et al (2014) | This work was supported by tl |
| - | Swedish International Development Coo | - | - | - | | Afolabi et al (2016) | Trials Partnership (EDCTP) an |
| - | Swedish International Development Coo | - | - | - | | Mensah (2017) | This work was supported by a |
| - | Swedish International Development Coo | - | - | - | | Mensah et al (2016) | This study was supported by a |
| - | Swedish International Development Coo | - | - | - | | Tiono (2018) | This work was supported by a |
| | The Coalition for Epidemic Preparedness | - | - | - | | Folegatti P.M.(2020) | UK Department of Health and |
| - | The Oxford Martin School | Rollier CS | - | - | | Reyes-Sandoval et al (2010 | We thank the Jenner Institu |
| EU FP7 | Transmolbloc | - | - | - | | Goodman et al (2011) | This work was supported prim |
| EU FP7 | Transmolbloc | - | - | - | | Goodman et al (2011) | This work was supported prim |
| - | TRANSVAC | - | - | - | | de Cassan et al (2011) | S.C.d.C. is a Ph.D. student sup |
| - | UK Department for International Develo | - | - | - | | Bowyer G (2020) | The Oxford clinical trial was s |
| - | UK Department for International Develo | - | - | - | | Venkatraman N (2019) | This work was supported |
| Project 16/107/03 | UK Department of Health and Social Car | - | - | - | | Stedman (2019) | This study was funded by the |
| 16/107/01 | UK Department of Health and Social Car | - | - | - | | Folegatti P.M.(2020) | UK Department of Health and |
| EP/R013756/1 | UK Engineeringand Physical Sciences Re | - | - | £6,968,179 | £ 6,968,179.00 | Fedosyuk S (2019) | This work was supported by M |
| - | UK Medical Research Council | - | - | - | - | Reyes-Sandoval (2010) | Work in the Oxford malaria va |
| - | UK National Institute for Health Researc | - | - | - | - | Folegatti P.M.(2020) | UK Department of Health and |
| MC_PC_19055 | UK Research and Innovation | Sarah Catherine Gilbert | Apr 20 - Sep 21 | £2,174,847 | £ 2,174,847.00 | Folegatti P.M.(2020) | UK Research and Innovation, ( |
| GR000550 | UK Royal Society for Tropical Medicine a | Coughlan L | - | - | - | Bliss (2020) | This research project was sup |
| 972216 | UKRI | Reyes-Sandoval A | Oct 16 - Sep 17 | £498,870 | £ 498,870.00 | Lopez-Camacho (2018) | This report is independent res |
| | UKRI | Alexandar Douglas | | £411,388.00 | £ 411,388.00 | https://www.ukri.org/research/c | This COVID-19 Rapid Respons |
| | UKRI | Sandy Douglas | | £400,000.00 | £ 400,000.00 | https://www.ox.ac.uk/news/202 | Working with Professor Sarah Gilbert |
| | UKRI | Graham Ogg | | £246,000.00 | £ 246,000.00 | https://www.ukri.org/resea | This £246k award is to procur |
| BBS/E/I/00007037 | UKRI Biotechnology and BiologicalScienc | Michael Johnson | Apr 17 - Mar 20 | £17,455,044 | £ 17,455,044.00 | S Graham (2020) | UKRI Biotechnology and Biolo |
| BBS/E/I/00007039 | UKRI Biotechnology and BiologicalScienc | Simon Thomas Carpenter | Apr 17 - Mar 20 | £6,662,753 | £ 6,662,753.00 | S Graham (2020) | UKRI Biotechnology and Biolo |
| BBS/E/I/00007031 | UKRI Biotechnology and BiologicalScienc | Philippa Beard | Apr 17 - Mar 20 | £2,965,523 | £ 2,965,523.00 | S Graham (2020) | UKRI Biotechnology and Biolo |
| BBS/E/I/00007034 | UKRI Biotechnology and BiologicalScienc | Simon Thomas Carpenter | Apr 17 - Mar 20 | £2,728,186 | £ 2,728,186.00 | S Graham (2020) | UKRI Biotechnology and Biolo |

| | | | | | | | | | |
|---|---|---|---|---|---|---|---|---|---|
| EP/R013756/1 | UKRI Engineering and Physical Sciences | Tarit K Mukhopadhyay | Apr 18 - Sep 21 | | £6,968,179 | £ | 6,968,179.00 | S Graham (2020) | This study was supported by U |
| EP/S025243/1 | UKRI Engineering and Physical Sciences | James Henderson Naismith | Nov 18 - May 20 | | £1,649,512 | £ | 1,649,512.00 | S Graham (2020) | andEPSRC Grant No. EP/S025 |
| - | University of Oxford | - | - | - | | | | Warimwe et al (2016) | This work was conducted with |
| MRF/TT2015/2150 | University of Oxford | Coughlan L | - | - | | | | Bliss (2020) | This research project was sup |
| | USAID | - | | - | | | | Payne et al (2017) | This work was supported by fu |
| - | Vaccine Research Center | - | - | - | | | | Tapia et al (2016) | This study was funded by a W |
| - | Vaccitech Ltd | - | | - | | | | Tuthill M. (2020) | European Union Seventh Fram |
| - | Wellcome Trust | - | - | - | | | | Ewer et al (2013) | The study was funded by gran |
| 76438 | Wellcome Trust | - | - | - | | - | | Reyes-Sandoval et al (2010 | We thank the Jenner Institu |
| 206194 | Wellcome Trust | - | - | - | | - | | Payne et al (2017) | This work was supported by fu |
| - | Wellcome Trust | - | - | - | | - | | Bowyer G (2020) | The Oxford clinical trial was s |
| - | Wellcome Trust | AVSH | - | - | | - | | Capone et al (2010) | This work was supported by th |
| - | Wellcome Trust | - | - | - | | - | | Capone et al (2010) | This work was supported by th |
| - | Wellcome Trust | Hill AVS | - | - | | - | | Capuccini et al (2017) | This work was supported by O |
| - | Wellcome Trust | - | - | - | | - | | Reyes-Sandoval (2010) | Work in the Oxford malaria va |
| - | Wellcome Trust | Reyes-Sandoval A | - | - | | - | | Reyes-Sandoval et al (2010 | We thank the Jenner Institu |
| - | Wellcome Trust | - | - | - | | - | | Rollier C (2020) | This work was funded by a gra |
| - | Wellcome Trust | Hill A.V.S. | - | - | | - | | Rollier C (2020) | This work was funded by a gra |
| - | Wellcome Trust | - | - | - | | - | | Venkatraman N (2019) | This work was supported |
| 084113/Z/07/Z | Wellcome Trust | - | - | - | | - | | Afolabi et al (2016) | Trials Partnership (EDCTP) and |
| 084113/Z/07/Z | Wellcome Trust | - | - | - | | - | | Longley et al (2017) | Funding for manufacture and |
| 084113/Z/07/Z | Wellcome Trust | - | - | - | | - | | Payne et al (2017) | This work was supported by fu |
| 095540/Z/11/Z | Wellcome Trust | Hill AVS | - | - | | - | | Longley et al (2017) | Funding for manufacture and |
| 095540/Z/11/Z | Wellcome Trust | Hill AVS | - | - | | - | | Salman et al (2017) | The work was funded by a We |
| 097395/Z/11/Z | Wellcome Trust | Reyes-Sandoval A | - | - | | - | | Alves et al (2017) | The work was funded by a We |
| 097395/Z/11/Z | Wellcome Trust | Reyes-Sandoval A | - | - | | - | | Salman et al (2017) | The work was funded by a We |
| 097940/Z/11/Z | Wellcome Trust | Hodgson SH | - | - | | - | | Longley et al (2017) | Funding for manufacture and |
| 106917/Z/15/Z | Wellcome Trust | Draper SJ | - | - | | - | | Payne et al (2017) | This work was supported by fu |
| 204826/Z/16/Z | Wellcome Trust | Prof Matthew Freeman | 07/09/2016 | | £3,000,000.00 | £ | 3,000,000.00 | Fedosyuk S (2019) | This work was supported by M |
| 106917/Z/15/Z | Wellcome Trust | Prof Simon Draper | 01/04/2015 | | £1,901,424.00 | £ | 1,901,424.00 | Wang (2018) | This work has been supported |
| 095540/Z/11/Z | Wellcome Trust | Hill A.V.S | 10/05/2011 | | £1,372,456.00 | £ | 1,372,456.00 | Atcheson (2018) | The work was funded by a We |
| 95540/Z/11/Z | Wellcome Trust | Prof Adrian Hill | 10/05/2011 | | £1,372,456.00 | £ | 1,372,456.00 | Dicks et al (2015 - 2) | This work has been funded by |
| 97395/Z/11/Z | Wellcome Trust | Reyes-Sandoval A | 12/12/2011 | | £1,081,461.00 | £ | 1,081,461.00 | Bauza et al (2014) | The work was funded by a We |
| 201477/Z/16/Z | Wellcome Trust | Dr Alexander Douglas | 18/05/2016 | | £414,492.00 | £ | 414,492.00 | Wang (2018) | This work has been supported |
| 201477/Z/16/Z | Wellcome Trust | Douglas AD | 18/05/2016 | | £414,492.00 | £ | 414,492.00 | Fedosyuk S (2019) | This work was supported by M |
| 098635/B/12/Z | Wellcome Trust | Warimwe GM | 21/06/2012 | | £253,778.00 | £ | 253,778.00 | Dulal et al (2016) | This study was conducted with |
| 098635/B/12/Z | Wellcome Trust | Warimwe GM | 21/06/2012 | | £253,778.00 | £ | 253,778.00 | Warimwe et al (2013) | This work was supported by a |
| 094449/Z/10/Z | Wellcome Trust | Duncan CJ | 31/08/2010 | | £218,216.00 | £ | 218,216.00 | Sheehy et al (2012) | This work was supported by |
| 089455/Z/09/Z | Wellcome Trust | Douglas AD | 29/05/2009 | | £217,651 | £ | 217,651.00 | Payne et al (2017) | This work was supported by fu |
| 97395/Z/11/A | Wellcome Trust | Reyes-Sandoval A | 01/08/2013 | | £203,200.00 | £ | 203,200.00 | | |
| 097940/Z/11/Z | Wellcome Trust | Hodgson SH | 29/03/2012 | | £195,304.00 | £ | 195,304.00 | Kimani et al (2014) | This work was supported by |
| 76438 | Wellcome Trust | Prof Adrian Hill | 16/09/2008 | | £84,023.00 | £ | 84,023.00 | Reyes-Sandoval et al (2008 | The work was supported by a |
| 098635/Z/12/Z | Wellcome Trust | Warimwe GM | 21/06/2012 | | £74,551.00 | £ | 74,551.00 | Dulal et al (2016) | This study was conducted with |
| - | Wellcome Trust | - | - | - | | | | Barnes et al (2012) | European Union (Framework V |

| | | | | | | |
|---|---|---|---|---|---|---|
| - | Wellcome Trust | - | - | - | Bliss (2018) | This study was funded by the |
| - | Wellcome Trust | - | - | - | Boyd et al (2013) | The Biotechnology and Biolog |
| - | Wellcome Trust | Hill AVS | - | - | Capuccini et al (2016) | This work was supported by C |
| - | Wellcome Trust | - | - | - | Colloca et al (2012) | This work was supported in pa |
| - | Wellcome Trust | - | - | - | Cottingham et al (2012) | This work was supported by th |
| - | Wellcome Trust | Hill AVS | - | - | de Cassan et al (2011) | S.C.d.C. is a Ph.D. student sup |
| - | Wellcome Trust | - | - | - | Dicks et al (2012) | This work has been funded by |
| - | Wellcome Trust | Hill AVS | - | - | Dicks et al (2015 - 2) | This work has been funded by |
| - | Wellcome Trust | Hill A.V.S | - | - | Draper et al (2010) | This work was funded by the |
| - | Wellcome Trust | Hill A.V.S. | - | - | Draper et al (2011) | SB was funded by MalParTrai |
| - | Wellcome Trust | Gilbert SC | - | - | Draper et al (2011) | SB was funded by MalParTrai |
| - | Wellcome Trust | - | - | - | Ewer et al (2013) | The study was funded by gran |
| - | Wellcome Trust | Hill AVS | - | - | Ewer et al (2013) | The study was funded by gran |
| - | Wellcome Trust | - | - | - | Forbes et al (2012) | This work was funded by the |
| - | Wellcome Trust | Hill AVS | - | - | Forbes et al (2012) | This work was funded by the |
| - | Wellcome Trust | Hill A.V.S | - | - | Gola (2018) | A.G. is funded by the Wellcom |
| - | Wellcome Trust | Hill A.V.S | - | - | Gola (2018) | A.G. is funded by the Wellcom |
| - | Wellcome Trust | Hill A.V.S. | - | - | Goodman et al (2011) | This work was supported prim |
| - | Wellcome Trust | - | - | - | Halbroth (2018) | BRH received funding from th |
| - | Wellcome Trust | - | - | - | Kelly et al (2016) | Supported by a Medical Resea |
| - | Wellcome Trust | Gavin Screaton | - | - | Lopez-Camacho (2018) | This report is independent res |
| - | Wellcome Trust | - | - | - | Tapia et al (2016) | This study was funded by a W |
| - | Wellcome Trust | - | - | - | Tapia et al (2016) | This study was funded by a W |
| - | Wellcome Trust | Hill AVS | - | - | Walker et al (2015) | Andrew S. Walker, José Loure |
| - | Wellcome Trust | Hill A.V.S | | | Halbroth (2018) | BRH received funding from th |
| - | Wellcome Trust | Hill AVS | - | - | Colloca et al (2012) | This work was supported in pa |
| - | Wellcome Trust | - | - | - | Ewer et al (2016) | Supported by the Wellcome T |
| - | Wellcome Trust | Kelly C | - | - | Swadling (2016) | Supported by the Medical Res |
| - | Wellcome Trust | Klenerman P | - | - | Swadling (2016) | Supported by the Medical Res |
| 084113/Z/07/Z | Wellcome Trust | - | - | - | de Barra et al (2014) | The study was funded by a gra |
| 084113/Z/07/Z | Wellcome Trust | - | - | - | Elias et al (2013) | This work was supported by th |
| 084113/Z/07/Z | Wellcome Trust | - | - | - | Hodgson et al (2014) | This work was supported by th |
| 084113/Z/07/Z | Wellcome Trust | Hill AVS | - | - | Hodgson et al (2015) | This work was supported by th |
| 084113/Z/07/Z | Wellcome Trust | - | - | - | Kimani et al (2014) | This work was supported by |
| 084113/Z/07/Z | Wellcome Trust | - | - | - | Sheehy et al (2011) | This work was supported by th |
| 084113/Z/07/Z | Wellcome Trust | - | - | - | Sheehy et al (2012 - 2) | This work was supported by th |
| 084113/Z/07/Z | Wellcome Trust | Prof Adrian Hill | - | - | Sheehy et al (2012) | This work was supported by |
| 091663MA | Wellcome Trust | - | - | - | Colston et al (2016) | This work was supported by W |
| 095540/Z/11/Z | Wellcome Trust | - | - | - | Dicks et al (2015) | This work has been funded by |
| 095540/Z/11/Z | Wellcome Trust | - | - | - | Longley et al (2015) | This work has been funded by |
| 095540/Z/11/Z | Wellcome Trust | - | - | - | Spencer et al (2014) | This work has been funded by |
| 097940/Z/11/Z | Wellcome Trust | Hodgson SH | - | - | de Barra et al (2014) | The study was funded by a gra |
| 097940/Z/11/Z | Wellcome Trust | Hodgson SH | - | - | Hodgson et al (2015) | This work was supported by th |
| 097940/Z/11/Z) | Wellcome Trust | Hodgson SH | - | - | Biswas et al (2014) | This work was supported by th |
| 099897/Z/12/A | Wellcome Trust | - | - | - | Colston et al (2016) | This work was supported by W |
| 45488/Z/05 | Wellcome Trust | Hill AVS | - | - | de Barra et al (2014) | The study was funded by a gra |
| 45488/Z/05 | Wellcome Trust | Hill AVS | - | - | Hodgson et al (2015) | This work was supported by th |
| 45488/Z/05 | Wellcome Trust | Hill AVS | - | - | Sheehy et al (2011) | This work was supported by th |
| 45488/Z/05 | Wellcome Trust | Hill AVS | - | - | Sheehy et al (2012 - 2) | This work was supported by th |
| 45488/Z/05 | Wellcome Trust | Hill AVS | - | - | Sheehy et al (2012) | This work was supported by |
| RTEI0 | Wellcome Trust | Duncan CJ | - | - | Sheehy et al (2012 - 2) | This work was supported by th |
| WT076943MA | Wellcome Trust | McShane H | - | - | Betts et al (2012) | Funding was provided by NEW |
| WT098051 | Wellcome Trust | - | - | - | Bauza et al (2014) | The work was funded by a We |
| WT098635 | Wellcome Trust | Warimwe GM | - | - | Warimwe et al (2016) | This work was conducted with |

| | | | | | | | |
|---|---|---|---|---|---|---|---|
| 97395 | Wellcome Trust | Reyes-Sandoval A | - | - | - | Betts et al (2012) | Funding was provided by NEW |
| - | Wellcome Trust | Hill AVS | - | - | - | Dicks et al (2012) | This work has been funded by |
| - | Wellcome Trust | - | - | - | - | O'Hara et al (2012) | Financial support. This work w |
| 084113/Z/07/Z | Wellcome Trust | - | - | - | - | Ogwang et al (2013) | This work was performed by t |
| WT 098051 | Wellcome Trust | Otto TD | - | - | - | Payne (2017) | This work was supported by a |
| 203077/Z/16/Z | Wellcome Trust | Prof Philip Bejon | 30/06/2016 | £26,595,243.00 | £ 26,595,243.00 | Stedman (2019) | This study was funded by the |
| 084113/Z/07/Z | Wellcome Trust | Prof Adrian Hill | 07/11/2007 | £3,400,000.00 | £ 3,400,000.00 | de Cassan et al (2011) | S.C.d.C. is a Ph.D. student sup |
| 084113/Z/07/Z | Wellcome Trust | Prof Adrian Hill | 07/11/2007 | £3,400,000.00 | £ 3,400,000.00 | Payne (2017) | This work was supported by a |
| 106325/Z/14/A | Wellcome Trust | A.V.S Hill | 19/12/2014 | £2,100,000.00 | £ 2,100,000.00 | Venkatraman N (2019) | This work was supported |
| 106917/Z/15/Z | Wellcome Trust | Draper SJ | 01/04/2015 | £1,901,424.00 | £ 1,901,424.00 | Payne (2017) | This work was supported by a |
| 097395/Z/11/Z | Wellcome Trust | Reyes-Sandoval A | 12/12/2011 | £1,081,461.00 | £ 1,081,461.00 | Atcheson (2018) | The work was funded by a We |
| 108734/Z/15/Z | Wellcome Trust | Rawlinson TA | 24/06/2015 | £271,399.00 | £ 271,399.00 | Payne (2017) | This work was supported by a |
| - | Wellcome Trust | A.V.S Hill | - | - | | Bowyer (2018) | The clinical trial was supporte |
| - | Wellcome Trust | Gola A | - | - | | Gola (2018) | A.G. is funded by the Wellcom |
| - | Wellcome Trust | - | - | - | | Mensah (2017) | This work was supported by a |
| - | Wellcome Trust Clinical Research Facilit | - | - | - | | Barnes et al (2012) | European Union (Framework |
