## Supplementary File 3 for "Who funded the research behind the Oxford-AstraZeneca COVID-19 vaccine? Approximating the funding to the University of Oxford for the research and development of the ChAdOx vaccine technology"

INFORMATION COMPLIANCE TEAM
University Offices, Wellington Square, Oxford OX1 2JD

UNIVERSITY OF OXFORD

Ref. FOI/20201025/03

19 November 2020

| Reply to request for information under the Freedom of Information Act | |
|---|---|
| Your ref | Email of 25 October 2020 |
| Request | 1. A breakdown of all funding (including all financial support, grants, donations, etc.) that has been received by the University of Oxford and the Jenner institute relating to ChAdOx vaccine platform, from the earliest date for which information is available to the present day. Please include:<br>1A. the grant number (if available) and date.<br>1B. Please break funding down into 1) funding from public (governmental) bodies, 2) private entities, and 3) philanthropies/charitable bodies.<br>1C. Please include the full name of the funding body.<br>1D. Please specify amounts in GBP.<br>Relevant keywords for such funding may include: ChAdOx1, ChAdOx2, VTP500.<br><br>2. What is the amount of grant funding from government entities (such as the Medical Research Council, or other research funders) that the University of Oxford and the Jenner Institute has received, since 1 January 2020, for projects relating to the development of the ChAdOx1 nCoV-19 vaccine? Please provide a breakdown of funding which includes the grant number (if available) and date and provide the full name of the funding body. Please specify amounts in GBP. Relevant keywords for such funding may include: AZD12222, ChAdOx1 nCoV-19, ChAdOx1, Oxford (COVID-19) vaccine, ChAdOx1 trials, SARS-COV-2 vaccine, VTP500.<br><br>3. Please specify the amount that the University of Oxford and the Jenner Institute has received from AstraZeneca, since 1 January 2020, for projects relating to the development of the ChAdOx1 nCoV-19 vaccine? Please provide a breakdown of funding giving the relevant dates and specifying amounts in GBP. Relevant keywords for such funding may include: AZD12222, ChAdOx1 nCoV-19, ChAdOx1, Oxford (COVID-19) vaccine, ChAdOx1 trials, SARS-COV-2 vaccine, VTP500. |

Dear Sarai Keestra,

I write in reply to your email to 25 October 2020, requesting the information shown above.

**Item 1**

We will not comply with this request in its current form, as we estimate that the time required to determine whether the information requested is held, and to locate, retrieve and extract it, would exceed the maximum amount of time a public authority is required to spend on a single request, namely, 18 hours.

The research covered by the request goes back over 10 years and involves a large number of investigators and research teams. As a result, the information requested is not readily available from a single system, file or set of files, but would need to be collected manually from different teams or individuals, and from information held on a number of different systems and files.  it is taking time to locate and extract the information. We apologise for the delay and hope to be able to provide the information no later than 11 December.

For these reasons, we are refusing your request under section 12 of the Freedom of Information Act (FOIA). Section 12 allows a public authority to refuse a request for information if the authority estimates that the cost of complying with it would exceed the 'appropriate limit' prescribed in the Freedom of Information and Data Protection (Appropriate Limit and Fees) Regulations 2004 (the 'Regulations'). The appropriate limit for universities is £450, which, because the Regulations fix staff costs at £25 an hour, corresponds to a time limit of 18 hours or just over two working days.

**Item 2**

The table below shows the funding received from charities as well as public sector bodies.

| Funder | Funder ref | Amount (£) | Start date |
|---|---|---|---|
| Coalition for Epidemic Preparedness Innovations | RROX2001 | 272,286.00 | 13/01/2020 |
| Medical Research Council | MC_PC_19055 | 2,174,847.97 | 01/01/2020 |
| Department of Health and Social Care | COV19 OxfordVacc-01 | 31,179,621.00 | 01/01/2020 |
| Wellcome Trust | 220991/Z/20/Z | 1,217,834.78 | 01/07/2020 |
| Chinese Academy of Medical Sciences | Mapping the human immune response post-vaccination with ChAdOx1 nCoV-19 | 68,106.34 | 01/03/2020 |

**Item 3**

I confirm that the University holds information that falls within the scope of this request. However, we are not yet in a position to respond, as we first need to consult Astra Zeneca in order to determine whether they consider any of the information to be exempt from disclosure. This will be done in accordance with section 3 of the Code of Practice on Freedom of Information issued under section 45 of the FOIA.

**INTERNAL REVIEW**

**THE INFORMATION COMMISSIONER**

If, after the internal review, you are still dissatisfied, you have the right under FOIA to apply to the Information Commissioner for a decision as to whether your request has been dealt with in accordance with the FOIA. You can do this online using the Information Commissioner's complaints portal.

Yours sincerely

**Information Compliance Team**

INFORMATION COMPLIANCE TEAM
University Offices, Wellington Square, Oxford OX1 2JD

UNIVERSITY OF OXFORD

Ref. FOI/20201025/03

14 December 2020

| Reply to request for information under the Freedom of Information Act | |
|---|---|
| Your ref | Email of 25 October 2020 |
| Request | 1. A breakdown of all funding (including all financial support, grants, donations, etc.) that has been received by the University of Oxford and the Jenner institute relating to ChAdOx vaccine platform, from the earliest date for which information is available to the present day. Please include:<br>1A. the grant number (if available) and date.<br>1B. Please break funding down into 1) funding from public (governmental) bodies, 2) private entities, and 3) philanthropies/charitable bodies.<br>1C. Please include the full name of the funding body.<br>1D. Please specify amounts in GBP.<br>Relevant keywords for such funding may include: ChAdOx1, ChAdOx2, VTP500.<br><br>2. What is the amount of grant funding from government entities (such as the Medical Research Council, or other research funders) that the University of Oxford and the Jenner Institute has received, since 1 January 2020, for projects relating to the development of the ChAdOx1 nCoV-19 vaccine? Please provide a breakdown of funding which includes the grant number (if available) and date and provide the full name of the funding body. Please specify amounts in GBP. Relevant keywords for such funding may include: AZD12222, ChAdOx1 nCoV-19, ChAdOx1, Oxford (COVID-19) vaccine, ChAdOx1 trials, SARS-COV-2 vaccine, VTP500.<br><br>3. Please specify the amount that the University of Oxford and the Jenner Institute has received from AstraZeneca, since 1 January 2020, for projects relating to the development of the ChAdOx1 nCoV-19 vaccine? Please provide a breakdown of funding giving the relevant dates and specifying amounts in GBP. Relevant keywords for such funding may include: AZD12222, ChAdOx1 nCoV-19, ChAdOx1, Oxford (COVID-19) vaccine, ChAdOx1 trials, SARS-COV-2 vaccine, VTP500. |

Dear Sarai Keestra,

I write in reply to your email to 25 October 2020, requesting the information shown above.

**Item 1**

We will not comply with this request in its current form, as we estimate that the time required to determine whether the information requested is held, and to locate, retrieve and extract it, would exceed the maximum amount of time a public authority is required to spend on a single request, namely, 18 hours.

The research covered by the request goes back over 10 years and involves a large number of investigators and research teams. As a result, the information requested is not readily available from a single system, file or set of files, but would need to be collected manually from different teams or individuals, and from information held on a number of different systems and files.

For these reasons, we are refusing your request under section 12 of the Freedom of Information Act (FOIA). Section 12 allows a public authority to refuse a request for information if the authority estimates that the cost of complying with it would exceed the 'appropriate limit' prescribed in the Freedom of Information and Data Protection (Appropriate Limit and Fees) Regulations 2004 (the 'Regulations'). The appropriate limit for universities is £450, which, because the Regulations fix staff costs at £25 an hour, corresponds to a time limit of 18 hours or just over two working days.

**Item 2**

The table below shows the funding received from charities as well as public sector bodies.

| Funder | Funder ref | Amount (£) | Start date |
|---|---|---|---|
| Coalition for Epidemic Preparedness Innovations | RROX2001 | 272,286.00 | 13/01/2020 |
| Medical Research Council | MC_PC_19055 | 2,174,847.97 | 01/01/2020 |
| Department of Health and Social Care | COV19 OxfordVacc-01 | 31,179,621.00 | 01/01/2020 |
| Wellcome Trust | 220991/Z/20/Z | 1,217,834.78 | 01/07/2020 |
| Chinese Academy of Medical Sciences | Mapping the human immune response post-vaccination with ChAdOx1 nCoV-19 | 68,106.34 | 01/03/2020 |

**Item 3**

At the date of your request, we had not received any funding from Astra Zeneca to meet the cost of the vaccine trials.

**INTERNAL REVIEW**

**THE INFORMATION COMMISSIONER**

If, after the internal review, you are still dissatisfied, you have the right under FOIA to apply to the Information Commissioner for a decision as to whether your request has been dealt with in accordance with the FOIA. You can do this online using the Information Commissioner's complaints portal.

Yours sincerely

**Information Compliance Team**

INFORMATION COMPLIANCE TEAM
University Offices, Wellington Square, Oxford OX1 2JD

UNIVERSITY OF OXFORD

Ref. FOI/20201130/03

27 January 2021

| Reply to request for information under the Freedom of Information Act | |
|---|---|
| Your ref | Email of 30 November 2020 |
| Request | 1. a spreadsheet with the funder, amount, title and date of all funding (including all financial support, grants, donations, etc.) to Sarah Gilbert from the year 2000 to the earliest date available.<br><br>2. a spreadsheet with the funder, amount, title and date of all funding (including all financial support, grants, donations, etc.) to Andrew [*sic*] Hill from the year 2000 to the earliest date available. |

Dear Sarai Keestra,

I write in reply to your email to 30 November 2020, requesting the information shown above. I apologise for the long delay in replying.

I attach a spreadsheet showing (a) the amount received in donations; and (b) the amount received in research grants.

**Donations**

We have withheld the names of three donors who wish to remain anonymous and the amount provided by Waif Said. We are exempting this information under section 43(2) of the Freedom of Information Act (FOIA). Section 43(2) provides that information is exempt where its disclosure would, or would be likely to, prejudice the commercial interests of any person. In our view, disclosure of the names of the donors that have requested anonymity would be likely to prejudice the University's commercial interests, by making it more difficult to raise funds from private donors in the future. The University's Donor Charter states that the University will respect anonymity where requested[1]. To breach this commitment by disclosing the identity of donors under the FOIA would undermine trust in the University and deter prospective donors. The University competes for a limited supply of donations with institutions that are not subject to the FOIA, such as charities or overseas universities. If it became known that the University could not ensure anonymity, prospective donors who would otherwise wish to support the University could decide to favour other institutions. Likewise, to disclose the size of a donation which a donor has requested be kept confidential would have a similarly negative effect on the University's ability to secure private funding.

Section 43(2) is a qualified exemption that requires the University to weigh the public interest in favour of disclosure, which is presumed from the FOIA, against the public interest in withholding the information.

We accept that there is a legitimate public interest in knowing who donates to the University and how much they give. Most donors are willing to be identified, and their gifts are acknowledged and given appropriate

[1] https://www.development.ox.ac.uk/donate/donor-charter

publicity. The names of those who have given £25,000 or more are published in the University's Campaign Report, with the consent of the donors. In addition, each year we publish in the University Calendar the names of the members of the Chancellor's Court of Benefactors and the Vice-Chancellor's Circle, which comprise the University's larger benefactors.[1]

However, a small number of donors wish to remain anonymous, or do not wish the size of their donation to be made public. It is important that we respect the wishes of these donors. There is a strong public interest in maintaining the University's ability to raise private funds, particularly at a time when public funding for higher education is likely in future to remain under severe pressure as a result of the pandemic. The funds raised from private sources are used to support the University's research, teaching and related activities, including the provision of bursaries for poorer students and activities to widen access. The University's high standards of research and teaching are valuable and important in their own right, and of public benefit. The development of a Covid vaccine, in partnership with Astra Zeneca, that is available at a lower price than some of the alternatives and that can be stored at relatively higher temperatures is a prime example. The University's research and teaching also benefit the UK's economy; and enhance the UK's international standing and reputation e.g. by attracting students and researchers from overseas to work at the University. The maintenance of these standards would be jeopardized if the University was less able to attract private funds, and this would not be in the public interest.

For these reasons, the University considers that the public interest in maintaining the exemption in Section 43(2) outweighs the public interest in disclosure.

**Research grants**

In accordance with section 3 of the Government's Code of Practice on Freedom of Information, it is our normal practice to consult companies before responding to a request for information that relates to them. However, since this response is already late, to avoid further delay, I have withheld the names of companies from the attached spreadsheet, where this information not already in the public domain. The names of public sector bodies and charities have been retained. Please let me know whether you wish to know the name of the private sector companies and I will consult them on disclosure.

**INTERNAL REVIEW**

**THE INFORMATION COMMISSIONER**

If, after the internal review, you are still dissatisfied, you have the right under FOIA to apply to the Information Commissioner for a decision as to whether your request has been dealt with in accordance with the FOIA. You can do this online using the Information Commissioner's complaints portal.

Yours sincerely

**Information Compliance Team**

[1] Further information is here: http://www.campaign.ox.ac.uk/contribute/recognising_your_gift/

INFORMATION COMPLIANCE TEAM
University Offices, Wellington Square, Oxford OX1 2JD

UNIVERSITY OF OXFORD

Ref. FOI/20210205/02

02 March 2021

| **Reply to request for information under the Freedom of Information Act** | |
|---|---|
| Your ref | Email of 5 February 2021 |
| Request | Freedom of Information request - Funding for Oxford AstraZeneca Vaccine<br>Under the Freedom of Information Act 2000 I would like to request a disclosure of all funding (including all financial support, research grants, contributions towards equipment and/or infrastructure, etc.) that the University of Oxford has received from AstraZeneca and the Serum Institute of India, since the 1st of January 2020 until the present date (5th of February 2021). Can you please provide this information in a spreadsheet with amounts in GBP and dates included as well. Thank you and I am looking forward to hear from you soon. |

Dear Ms Pugh-Jones,

I write in reply to your email of 5 February 2021, requesting the information shown above.

The University has not received any payments from AstraZeneca or the Serum Institute towards the cost of the Oxford-AstraZeneca covid-19 vaccine in the period covered by your request.

Yours sincerely

**Information Compliance Team**
