## Supplementary File 4 for "Who funded the research behind the Oxford-AstraZeneca COVID-19 vaccine? Approximating the funding to the University of Oxford for the research and development of the ChAdOx vaccine technology"

| SUMMARY OF DONATIONS (CASH RECEIVED) TO THE UNIVERSITY OF OXFORD IN SUPPORT OF PROFFESOR SARAH GILBERT AND PROFESSOR ADRIAN HILL to 17/12/2020 | | | |
|---|---|---|---|
| **Donor** | **Donation** | **Date** | **Title of funding** |
| Anonymous | £750,000 - £999,999 | 15/04/2020 | To support Professor Adrian Hill's work on COVID-19 vaccine |
| Anonymous | £100,000 - £249,999 | 15/04/2020 | To support Professor Adrian Hill's work on COVID-19 vaccine development |
| Anonymous | £100,000 - £249,999 | 21/04/2020 | To scale production and manufacturing of Oxford's Vaccine Candidate; Jenner Institute for Professor Adrian Hill |
| FIAP, International Federation of Photographic Art | £25,000 - £49,999 | 14/09/2020 | Professor Adrian Hill and Professor Sarah Gilbert – vaccine research |
| Karin B. Sinniger | £25,000 - £49,999 | 12/05/2020 | To support Professor Adrian Hill's vaccine research |
| Lakshmi Mittal | £500,000 - £750,000 | 03/07/2020 | Lakshmi Mittal and Family Professorship for Vaccinology, Professor Adrian Hill |
| Richard A. Sanders | £250,000 - £499,999 | 23/04/2020 | Professor Adrian Hill and Professor Sarah Gilbert – vaccine research |
| TrustBridge Global | £25,000 - £49,999 | 13/08/2020 | Professor Adrian Hill and Professor Sarah Gilbert – vaccine research |
| Wafic R. Saïd | Confidential | 28/07/2020 | Saïd Professorship of Vaccinology, Professor Sarah Gilbert |

| Sponsor | Project Number | Funding Type | Project Name | Principal Investigator | Project Start Date | Project End Date | Total Budget | Classification |
|---|---|---|---|---|---|---|---|---|
| European Commission | HCRGZJ00 | EU Government | AN INTEGRATED PROJECT FOR THE DESIGN & TESTING OF VACCINE CANDIDATES AGAINST TUBERCULOSIS:IDENTIFCATION | Hill, Prof. Adrian | 01-Jan-2004 | 31-Jan-2010 | 371,005.17 | Other vaccine research |
| European Commission | HCRGZJ00 | EU Government | AN INTEGRATED PROJECT FOR THE DESIGN & TESTING OF VACCINE CANDIDATES AGAINST TUBERCULOSIS:IDENTIFCATION | Hill, Prof. Adrian | 01-Jan-2004 | 31-Jan-2010 | 423,717.11 | Other vaccine research |
| Jenner Vaccine Foundation | HCRIPL00 | UK Charity (no QR) | EJIVR fellowship | Hill, Prof. Adrian | 01-Aug-2005 | 30-Sep-2014 | 12,723.28 | Not relevant to ChAdOx |
| Jenner Vaccine Foundation | HCRIPL00 | UK Charity (no QR) | EJIVR fellowship | Hill, Prof. Adrian | 01-Aug-2005 | 30-Sep-2014 | 247,066.09 | Not relevant to ChAdOx |
| Jenner Vaccine Foundation | HCRIPL00 | UK Charity (no QR) | EJIVR fellowship | Hill, Prof. Adrian | 01-Aug-2005 | 30-Sep-2014 | 597,272.77 | Not relevant to ChAdOx |
| Foundation for National Institutes of Health | HCRJFN00 | Non-EU Other | ENHANCING THE IMMUNOGENICITY AND EFFICACY OF VECTORED VACCINES | Hill, Prof. Adrian | 01-Aug-2005 | 30-Nov-2013 | 1,928,792.16 | ChAdOx technology |
| Foundation for National Institutes of Health | HCRJFN00 | Non-EU Other | ENHANCING THE IMMUNOGENICITY AND EFFICACY OF VECTORED VACCINES | Hill, Prof. Adrian | 01-Aug-2005 | 30-Nov-2013 | 390,640.00 | ChAdOx technology |
| Foundation for National Institutes of Health | HCRJFN00 | Non-EU Other | ENHANCING THE IMMUNOGENICITY AND EFFICACY OF VECTORED VACCINES | Hill, Prof. Adrian | 01-Aug-2005 | 30-Nov-2013 | 3,301,513.13 | ChAdOx technology |
| Foundation for National Institutes of Health | HCRJFN00 | Non-EU Other | ENHANCING THE IMMUNOGENICITY AND EFFICACY OF VECTORED VACCINES | Hill, Prof. Adrian | 01-Aug-2005 | 30-Nov-2013 | 108,347.00 | ChAdOx technology |
| European Commission | HCRNHU00 | EU Government | New Preventative and therapeutic Hepatits C Vaccines: from pre-clinical to phase 1 (HEPACIVAC) | Hill, Prof. Adrian | 01-Feb-2007 | 06-Nov-2012 | 905,482.61 | Other vaccine research |
| Department of Health and Social Care | HCRNFZ00 | UK Public Sector | Vaccines Theme | Hill, Prof. Adrian | 01-Apr-2007 | 30-Jun-2012 | 1,887,190.90 | ChAdOx technology |
| Department of Health and Social Care | HCRNFZ00 | UK Public Sector | Vaccines Theme | Hill, Prof. Adrian | 01-Apr-2007 | 30-Jun-2012 | 97,763.92 | ChAdOx technology |
| Department of Health and Social Care | HCRNFZ00 | UK Public Sector | Vaccines Theme | Hill, Prof. Adrian | 01-Apr-2007 | 30-Jun-2012 | 37,846.92 | ChAdOx technology |
| Department of Health and Social Care | HCRNFZ00 | UK Public Sector | Vaccines Theme | Hill, Prof. Adrian | 01-Apr-2007 | 30-Jun-2012 | 1,327,807.87 | ChAdOx technology |
| Department of Health and Social Care | HCRNFZ00 | UK Public Sector | Vaccines Theme | Hill, Prof. Adrian | 01-Apr-2007 | 30-Jun-2012 | 14,760.28 | ChAdOx technology |
| Wellcome Trust | HCROPS00 | UK Charity (QR) | Human and veterinary vaccinology | Hill, Prof. Adrian | 01-Jul-2008 | 30-Sep-2013 | 2,500,000.00 | ChAdOx technology |
| European & Developing Countries Clinical Trials Partnership | HCRRZI10 | EU Government | INTEGRATING CAPACITY BUILDING AND NETWORKING IN THE DESIGN AND CINDUCT OF PHASE 1 AND 11 CLINICAL TRIALS OF | Hill, Prof. Adrian | 01-Nov-2009 | 31-May-2015 | 2,038,038.98 | ChAdOx technology |
| European Commission | HCRRVU00 | EU Government | IDEA: DISSECTING THE IMMUNOLOGICAL INTERPLAY BETWEEN POVERTY RELATED DISEASES AND HELMINTH INFECTIONS: A | Hill, Prof. Adrian | 01-Mar-2010 | 31-Oct-2015 | 638,805.39 | Other vaccine research |
| James Martin (Individual) | HCRSKB00 | Non-EU Other | VACCINE DESIGN INSTITUTE (MATCHED FUNDING) | Hill, Prof. Adrian | 01-Jun-2010 | 31-Aug-2013 | 129,311.00 | Not relevant to ChAdOx |
| James Martin (Individual) | HCRSKB00 | Non-EU Other | VACCINE DESIGN INSTITUTE (MATCHED FUNDING) | Hill, Prof. Adrian | 01-Jun-2010 | 31-Aug-2013 | 145,796.00 | Not relevant to ChAdOx |
| James Martin (Individual) | HCRSKB00 | Non-EU Other | VACCINE DESIGN INSTITUTE (MATCHED FUNDING) | Hill, Prof. Adrian | 01-Jun-2010 | 31-Aug-2013 | 444,324.00 | Not relevant to ChAdOx |
| James Martin (Individual) | HCRSKB00 | Non-EU Other | VACCINE DESIGN INSTITUTE (MATCHED FUNDING) | Hill, Prof. Adrian | 01-Jun-2010 | 31-Aug-2013 | 247,210.00 | Not relevant to ChAdOx |
| European Vaccine Initiative | HCRUXL00 | EU Government | Construction and GMP manufacture of AdCh63 CSP and MVA CSP plus clinical trials | Hill, Prof. Adrian | 01-Aug-2010 | 22-Apr-2014 | 497,082.81 | ChAdOx technology |
| Wellcome Trust | HCRUZZ00 | UK Charity (QR) | T-Cell Inducing Vaccines | Hill, Prof. Adrian | 01-Sep-2011 | 31-Dec-2014 | 68,228.00 | ChAdOx technology |
| Wellcome Trust | HCRUZZ00 | UK Charity (QR) | T-Cell Inducing Vaccines | Hill, Prof. Adrian | 01-Sep-2011 | 31-Dec-2014 | 1,304,228.00 | ChAdOx technology |
| Name withheld | HCRVJB00 | EU Industry (QR) | Vaccine Thermostability | Hill, Prof. Adrian | 01-Oct-2011 | 28-Feb-2014 | 149,707.00 | ChAdOx technology |
| Program for Assessment Technology in Health | HCRWJS00 | Non-EU Other | Clinical Testing of Multi-Antigen Adnovirus-Vectored Malaria Vaccines in Prime-Boost Regimens with DNA and MVA (VAC045)igen Adnovirus | Hill, Prof. Adrian | 15-Mar-2012 | 31-May-2014 | 574,070.40 | ChAdOx technology |
| Department of Health and Social Care | HCRWAF00 | UK Public Sector | BRC2 VACCINES THEME | Hill, Prof. Adrian | 01-Apr-2012 | 31-May-2015 | 36,468.48 | ChAdOx technology |
| Department of Health and Social Care | HCRWAF00 | UK Public Sector | BRC2 VACCINES THEME | Hill, Prof. Adrian | 01-Apr-2012 | 31-May-2015 | 1,764,079.63 | ChAdOx technology |
| Department of Health and Social Care | HCRWAF00 | UK Public Sector | BRC2 VACCINES THEME | | 01-Apr-2012 | 31-May-2015 | 933,269.78 | ChAdOx technology |
| European Commission | HCRWMY00 | EU Government | Immunogene: Immunogenetics of Severe Bacterial Disease Susceptibility and Vaccine Responses in Humans | Hill, Prof. Adrian | 01-Jun-2012 | 30-Sep-2017 | 1,095,942.39 | Other vaccine research |
| European Commission | HCRWMY00 | EU Government | Immunogene: Immunogenetics of Severe Bacterial Disease Susceptibility and Vaccine Responses in Humans | Hill, Prof. Adrian | 01-Jun-2012 | 30-Sep-2017 | 243,881.65 | Other vaccine re Hill, Prof. Adriar |
| European Commission | HCRXKA00 | EU Government | MULTIMALVAX - A MULTI STAGE MALARIA VACCINE | Hill, Prof. Adrian | 01-Oct-2012 | 20-Jul-2017 | 3,696,129.35 | ChAdOx technology |
| James Martin 21st Century Foundation | HCRXNO10 | Non-EU Other | Vaccines To Tackle Variable Pathogens: Transforming Immunisation Effectiveness in the 21st Century | Hill, Prof. Adrian | 01-Oct-2012 | 30-Nov-2015 | 28,930.00 | Not relevant to ChAdOx |
| James Martin 21st Century Foundation | HCRXNO10 | Non-EU Other | Vaccines To Tackle Variable Pathogens: Transforming Immunisation Effectiveness in the 21st Century | Hill, Prof. Adrian | 01-Oct-2012 | 30-Nov-2015 | 121,073.62 | Not relevant to ChAdOx |
| James Martin 21st Century Foundation | HCRXNO10 | Non-EU Other | Vaccines To Tackle Variable Pathogens: Transforming Immunisation Effectiveness in the 21st Century | Hill, Prof. Adrian | 01-Oct-2012 | 30-Nov-2015 | 6,390.00 | Not relevant to ChAdOx |
| James Martin 21st Century Foundation | HCRXNO10 | Non-EU Other | Vaccines To Tackle Variable Pathogens: Transforming Immunisation Effectiveness in the 21st Century | Hill, Prof. Adrian | 01-Oct-2012 | 30-Nov-2015 | 13,950.00 | Not relevant to ChAdOx |
| James Martin 21st Century Foundation | HCRXNO10 | Non-EU Other | Vaccines To Tackle Variable Pathogens: Transforming Immunisation Effectiveness in the 21st Century | Hill, Prof. Adrian | 01-Oct-2012 | 30-Nov-2015 | 30,000.00 | Not relevant to ChAdOx |
| James Martin 21st Century Foundation | HCRXNO10 | Non-EU Other | Vaccines To Tackle Variable Pathogens: Transforming Immunisation Effectiveness in the 21st Century | Hill, Prof. Adrian | 01-Oct-2012 | 30-Nov-2015 | 30,000.00 | Not relevant to ChAdOx |
| James Martin 21st Century Foundation | HCRXNO10 | Non-EU Other | Vaccines To Tackle Variable Pathogens: Transforming Immunisation Effectiveness in the 21st Century | Hill, Prof. Adrian | 01-Oct-2012 | 30-Nov-2015 | 106,032.38 | Not relevant to ChAdOx |
| James Martin 21st Century Foundation | HCRXNO10 | Non-EU Other | Vaccines To Tackle Variable Pathogens: Transforming Immunisation Effectiveness in the 21st Century | Hill, Prof. Adrian | 01-Oct-2012 | 30-Nov-2015 | 109,486.00 | Not relevant to ChAdOx |
| James Martin 21st Century Foundation | HCRXNO10 | Non-EU Other | Vaccines To Tackle Variable Pathogens: Transforming Immunisation Effectiveness in the 21st Century | Hill, Prof. Adrian | 01-Oct-2012 | 30-Nov-2015 | 4,138.00 | Not relevant to ChAdOx |
| European & Developing Countries Clinical Trials Partnership | HCRYLI00 | EU Government | FIELD TRILAS OF A NEW COMBINATION MALARIA VACCINE IN WEST AFRICAN ADULTS AND CHILDREN (MVVC2) | Hill, Prof. Adrian | 01-Nov-2012 | 30-Nov-2015 | 171,708.16 | Other vaccine research |
| European Commission | HCRZBY00 | EU Government | VACTRAIN - Training network for next generation vaccinologists | Hill, Prof. Adrian | 01-Nov-2012 | 28-Feb-2017 | 178,033.83 | Other vaccine research |
| European Commission | HCRZBY00 | EU Government | VACTRAIN - Training network for next generation vaccinologists | Hill, Prof. Adrian | 01-Nov-2012 | 28-Feb-2017 | 302,698.29 | ChAdOx technology |
| Medical Research Council | HCRXMA00 | Research Councils | CLINICAL ASSESSMENT OF A NOVEL SIMIAN ADENOVIRUS-VECTORED INFLUENZA VACCINE DESIGNED TO INDUCE BROADLY I | Gilbert, Prof. Sarah | 01-Dec-2012 | 31-Jul-2016 | 793,586.40 | ChAdOx technology |
| Bill & Melinda Gates Foundation | HCRXXG00 | Multiple Funding Types | VAC052 - MVI PATH. ADRIAN HILL | Hill, Prof. Adrian | 01-Dec-2012 | 31-Jan-2015 | 560,650.41 | ChAdOx technology |
| Medical Research Council | HCRXWW00 | Research Councils | DCS- CLINICAL EVALUATION OF A SPOROZITE AND LIVER STAGE COMBINATION VACCINE FOR PLASMODIUM FALCIPARUM | Hill, Prof. Adrian | 31-Dec-2012 | 30-Apr-2016 | 916,661.20 | Other vaccine research |
| Bill & Melinda Gates Foundation | HCR00050 | Non-EU Other | VAC 055. A phase I/IIa Sporozite challenge study to assess the protective efficacy of the combination malaria vaccine candidate regime of RTS, S | Hill, Prof. Adrian | 01-May-2013 | 28-Feb-2015 | 1,034,355.94 | ChAdOx technology |
| Bill & Melinda Gates Foundation | HCRYOT00 | Non-EU Charity (QR) | VAC 055 - A PHASE I/IIa SPOROZITE CHALLENGE STUDY TO ASSESS THE PROTECTIVE EFFICACY OF THE COMBINATION MALARI | Hill, Prof. Adrian | 01-May-2013 | 30-Jun-2014 | 0.00 | ChAdOx technology |
| Wellcome Trust | HCRYBS00 | UK Charity (QR) | ADVANCING HUMAN & VETERINARY VACCINOLOGY | Hill, Prof. Adrian | 30-Jun-2013 | 31-May-2017 | 940,440.51 | Not relevant to ChAdOx |
| Wellcome Trust | HCRYBS00 | UK Charity (QR) | ADVANCING HUMAN & VETERINARY VACCINOLOGY | Hill, Prof. Adrian | 30-Jun-2013 | 31-May-2017 | 1,119,154.49 | Not relevant to ChAdOx |
| Innovate UK | HCRZJK00 | UK Public Sector | Development of a novel VLP based vaccine for malaria | Hill, Prof. Adrian | 01-Sep-2013 | 31-Oct-2014 | 99,998.08 | Other vaccine research |
| Wellcome Trust | HCR00040 | UK Charity (QR) | An investigation of cell dynamics, migration and differentiation resulting in the generation of a CD8+ T-cell memory response following vaccinati | Hill, Prof. Adrian | 01-Oct-2013 | 30-Nov-2017 | 80,000.00 | ChAdOx technology |
| Animal Health and Veterinary Laboratories Agency | HCR00200 | UK Public Sector | Influenza project - Jenner fellow | Gilbert, Prof. Sarah | 01-Jan-2014 | 28-Feb-2017 | 65,038.79 | Not relevant to ChAdOx |
| Name withheld | HCR00100 | UK Industry (QR) | H1N1_CS_01 clinical study | Gilbert, Prof. Sarah | 01-Feb-2014 | 31-Dec-2014 | 11,426.00 | ChAdOx technology |
| European Commission | HCR00120 | EU Government | Improving prostate Cancer Outcomes with Vectored Vaccines | Hill, Prof. Adrian | 01-Apr-2014 | 15-Nov-2019 | 3,435,675.99 | ChAdOx technology |
| European Commission | HCR00120 | EU Government | Improving prostate Cancer Outcomes with Vectored Vaccines | Hill, Prof. Adrian | 01-Apr-2014 | 15-Nov-2019 | 18,872.52 | ChAdOx technology |
| Biotechnology & Biological Sciences Research Council | HCR00260 | Research Councils | Understanding influenza A virus: linking transmission, evolutionary dynamics, pathogenesis and immunity in pigs | Gilbert, Prof. Sarah | 01-Apr-2014 | 31-May-2019 | 44,838.00 | Other vaccine research |
| Program for Appropriate Technology in Health (PATH) | HCR00170 | Non-EU Other | A phase I/IIa sporozoite challenge study to assess the safety and protective efficacy of concomitant administration of the Combination Malaria Va | Hill, Prof. Adrian | 15-May-2014 | 31-Mar-2015 | 271,432.82 | ChAdOx technology |
| Wellcome Trust | HCR00240 | Split-Funded | Accelerated Clinical Evaluation of a Monovalent Vectored Ebola vaccine | Hill, Prof. Adrian | 01-Sep-2014 | 28-Feb-2017 | 2,820,000.00 | ChAdOx technology |
| Wellcome Trust | HCR00340 | UK Charity (QR) | A multi-component high efficacy malaria vaccine | Hill, Prof. Adrian | 01-Sep-2014 | 31-Oct-2019 | 53,470.15 | ChAdOx technology |
| Wellcome Trust | HCR00340 | UK Charity (QR) | A multi-component high efficacy malaria vaccine | Hill, Prof. Adrian | 01-Sep-2014 | 31-Oct-2019 | 1,952,079.53 | ChAdOx technology |
| Name withheld | HCR00171 | Non-EU Other | A Phase I/IIa Sporozoite Challenge study to access the safety and protective efficacy of concomitant administration of the combination malaria va | Hill, Prof. Adrian | 01-Oct-2014 | 30-Nov-2016 | 1,166,941.27 | ChAdOx technology |
| European Commission | HCR00320 | EU Government | Development of a Chimpanzee Adenovirus Type 3 Ebolavirus Zaire Vaccine | Hill, Prof. Adrian | 07-Oct-2014 | 31-Jul-2018 | 698,208.30 | ChAdOx technology |
| Wellcome Trust | HCR00310 | UK Charity (QR) | Large Scale Biomanufacture of a Monovalent Ebola MVA Vector and Heterologous Boosting of Primed Subjects in Mali and the UK | Hill, Prof. Adrian | 01-Dec-2014 | 28-Feb-2017 | 2,179,000.00 | Other vaccine research |
| European Commission | HCR00350 | EU Government | Standardization & development of assays for assessment of influenza vaccine correlates of protection | Gilbert, Prof. Sarah | 01-Mar-2015 | 30-Apr-2021 | 90,006.92 | Other vaccine research |
| NIHR Biomedical Research Centre | HCRWAF04 | UK Public Sector | BRC2 Vaccines Theme Year 4 | Hill, Prof. Adrian | 01-Apr-2015 | 31-May-2016 | 172,911.59 | ChAdOx technology |
| NIHR Biomedical Research Centre | HCRWAF04 | UK Public Sector | BRC2 Vaccines Theme Year 4 | Hill, Prof. Adrian | 01-Apr-2015 | 31-May-2016 | 438,045.00 | ChAdOx technology |
| NIHR Biomedical Research Centre | HCRWAF04 | UK Public Sector | BRC2 Vaccines Theme Year 4 | Hill, Prof. Adrian | 01-Apr-2015 | 31-May-2016 | 299,291.00 | ChAdOx technology |
| Janssen Vaccines & Prevention B.V. | HCR00480 | EU Industry (QR) | RELATING TO A PHASE I, SAFETY AND IMMUNOGENICITY TRIAL OF THE HETEROLOGOUS PRIME-BOOST REGIMEN COMBININ | Hill, Prof. Adrian | 01-May-2015 | 28-Feb-2020 | 343,921.36 | ChAdOx technology |
| Name withheld | HCR00481 | EU Industry (QR) | RELATING TO A PHASE I, SAFETY AND IMMUNOGENICITY TRIAL OF THE HETEROLOGOUS PRIME-BOOST REGIMEN COMBININ | Hill, Prof. Adrian | 01-May-2015 | 28-Feb-2020 | 0.00 | ChAdOx technology |
| Name withheld | HCR00481 | EU Industry (QR) | RELATING TO A PHASE I, SAFETY AND IMMUNOGENICITY TRIAL OF THE HETEROLOGOUS PRIME-BOOST REGIMEN COMBININ | Hill, Prof. Adrian | 01-May-2015 | 28-Feb-2020 | 343,921.37 | ChAdOx technology |

| | | | | | | | | |
|---|---|---|---|---|---|---|---|---|
| Biotechnology & Biological Sciences Research Council | HCR00410 | Research Councils | Stabilisation of Newcastle disease vaccine formulated in sugar-glass on polypropylene membranes | Hill, Prof. Adrian | 01-Jun-2015 | 31-Jan-2017 | 82,989.96 | Other vaccine research |
| Biotechnology & Biological Sciences Research Council | HCR00410 | Research Councils | Stabilisation of Newcastle disease vaccine formulated in sugar-glass on polypropylene membranes | Hill, Prof. Adrian | 01-Jun-2015 | 31-Jan-2017 | 57,856.44 | Other vaccine research |
| Jenner Vaccine Foundation | HCR00450 | UK Charity (no QR) | JVF Public Engagement Award | Hill, Prof. Adrian | 01-Jul-2015 | 28-Feb-2017 | 50,000.00 | Not relevant to ChAdOx |
| Name withheld | HCR00500 | UK Industry (QR) | Agreement for the manufacture of a vaccine for use in clinical trials relating to ChAdOx2 | Gilbert, Prof. Sarah | 03-Aug-2015 | 28-Feb-2017 | 918,721.00 | ChAdOx technology |
| Name withheld | HCR00530 | UK Industry (QR) | In support of the manufacture of MVA HAV for use in clinical trials and related toxicology testing/clinical trial support activities | Gilbert, Prof. Sarah | 01-Oct-2015 | 31-Jul-2020 | 424,033.35 | Other vaccine research |
| Name withheld | HCR00531 | UK Industry (QR) | In support of the manufacture of MVA HAV for use in clinical trials and related toxicology testing/clinical trial support activities. | Gilbert, Prof. Sarah | 01-Oct-2015 | 30-Nov-2018 | 52,622.40 | Other vaccine research |
| Jenner Vaccine Foundation | HCR00630 | UK Charity (no QR) | Jenner Vaccine Foundation | Hill, Prof. Adrian | 01-Jan-2016 | 28-Feb-2017 | 93,000.00 | Not relevant to ChAdOx |
| Name withheld | HCR00730 | EU Industry (QR) | Preparatory activities in support of the FLU007 clinical trial | Gilbert, Prof. Sarah | 01-Feb-2016 | 30-Sep-2016 | 46,242.49 | Other vaccine research |
| Medical Research Council | HCR00670 | Research Councils | Pre-clinical development of an influenza vaccine to induce broad protection through multiple immune mechanisms | Gilbert, Prof. Sarah | 01-Mar-2016 | 31-Jul-2018 | 379,318.93 | ChAdOx technology |
| Medical Research Council | HCR00670 | Research Councils | Pre-clinical development of an influenza vaccine to induce broad protection through multiple immune mechanisms | Gilbert, Prof. Sarah | 01-Mar-2016 | 31-Jul-2018 | 300,241.00 | ChAdOx technology |
| NIHR Biomedical Research Centre | HCRWAF05 | UK Public Sector | BRC2 Vaccines Theme Year 5 | Hill, Prof. Adrian | 01-Apr-2016 | 31-May-2017 | 172,912.00 | ChAdOx technology |
| NIHR Biomedical Research Centre | HCRWAF05 | UK Public Sector | BRC2 Vaccines Theme Year 5 | Hill, Prof. Adrian | 01-Apr-2016 | 31-May-2017 | 438,223.10 | ChAdOx technology |
| NIHR Biomedical Research Centre | HCRWAF05 | UK Public Sector | BRC2 Vaccines Theme Year 5 | Hill, Prof. Adrian | 01-Apr-2016 | 31-May-2017 | 296,014.03 | ChAdOx technology |
| Wellcome Trust | HCR00720 | UK Charity (QR) | Prime-Target Vaccination in Malaria | Hill, Prof. Adrian | 01-Jun-2016 | 31-Oct-2019 | 201,600.00 | ChAdOx technology |
| Name withheld | HCR00940 | UK Industry (QR) | INVICTUS Year 1 costs | Gilbert, Prof. Sarah | 01-Aug-2016 | 28-Feb-2018 | 384,306.14 | Other vaccine research |
| Name withheld | HCR00940 | UK Industry (QR) | INVICTUS Year 1 costs | Gilbert, Prof. Sarah | 01-Aug-2016 | 28-Feb-2018 | 151,549.02 | Other vaccine research |
| Medical Research Council | H5R00610 | Research Councils | The Mexican Biobank Project: Building Capacity for Big Data Science in Medical Genomics in Admixed Populations | Hill, Prof. Adrian | 14-Aug-2016 | 18-Nov-2020 | 191,033.58 | Other vaccine research |
| Medical Research Council | H5R00610 | Research Councils | The Mexican Biobank Project: Building Capacity for Big Data Science in Medical Genomics in Admixed Populations | Hill, Prof. Adrian | 14-Aug-2016 | 18-Nov-2020 | 316,308.00 | Other vaccine research |
| Medical Research Council | HCR00810 | Research Councils | Development of a novel biocompatible matrix for sugar membrane technology for thermostability of an adjuvanted malaria vaccine | Hill, Prof. Adrian | 01-Oct-2016 | 31-May-2018 | 48,239.00 | Other vaccine research |
| EMD Millipore Corporation | HCR00910 | Non-EU Industry (QR) | Millipore/Sigma CBF collaboration | Hill, Prof. Adrian | 01-Dec-2016 | 31-Jan-2019 | 79,636.07 | Not relevant to ChAdOx |
| EMD Millipore Corporation | HCR00910 | Non-EU Industry (QR) | Millipore/Sigma CBF collaboration | Hill, Prof. Adrian | 01-Dec-2016 | 31-Jan-2019 | 59,629.45 | Not relevant to ChAdOx |
| Royal Society | HCR01070 | Research Councils | Transcriptomics of African Fruit Bats in response to Ebola virus antigens | Gilbert, Prof. Sarah | 01-Dec-2016 | 31-Jan-2018 | 5,000.00 | Other vaccine research |
| Name withheld | HCR00870 | UK Industry (QR) | HAV-Trial | Gilbert, Prof. Sarah | 19-Dec-2016 | 28-Feb-2021 | 202,673.09 | ChAdOx technology |
| European Commission | HCR00890 | EU Government | Optimizing a deployable high efficacy malaria vaccine | Hill, Prof. Adrian | 01-Jan-2017 | 28-Feb-2022 | 20,222.39 | ChAdOx technology |
| European Commission | HCR00890 | EU Government | Optimizing a deployable high efficacy malaria vaccine | Hill, Prof. Adrian | 01-Jan-2017 | 28-Feb-2022 | 118,934.30 | ChAdOx technology |
| European Commission | HCR00890 | EU Government | Optimizing a deployable high efficacy malaria vaccine | Hill, Prof. Adrian | 01-Jan-2017 | 28-Feb-2022 | 0.01 | ChAdOx technology |
| European Commission | HCR00890 | EU Government | Optimizing a deployable high efficacy malaria vaccine | Hill, Prof. Adrian | 01-Jan-2017 | 28-Feb-2022 | 588,848.64 | ChAdOx technology |
| European Commission | HCR00890 | EU Government | Optimizing a deployable high efficacy malaria vaccine | Hill, Prof. Adrian | 01-Jan-2017 | 28-Feb-2022 | 495,448.15 | ChAdOx technology |
| European Commission | HCR00890 | EU Government | Optimizing a deployable high efficacy malaria vaccine | Hill, Prof. Adrian | 01-Jan-2017 | 28-Feb-2022 | 980,784.98 | ChAdOx technology |
| European Commission | HCR00890 | EU Government | Optimizing a deployable high efficacy malaria vaccine | Hill, Prof. Adrian | 01-Jan-2017 | 28-Feb-2022 | 717,894.57 | ChAdOx technology |
| European Commission | HCR00890 | EU Government | Optimizing a deployable high efficacy malaria vaccine | Hill, Prof. Adrian | 01-Jan-2017 | 28-Feb-2022 | 788,672.49 | ChAdOx technology |
| European Commission | HCR00890 | EU Government | Optimizing a deployable high efficacy malaria vaccine | Hill, Prof. Adrian | 01-Jan-2017 | 28-Feb-2022 | 3,033,815.56 | ChAdOx technology |
| European Commission | HCR00890 | EU Government | Optimizing a deployable high efficacy malaria vaccine | Hill, Prof. Adrian | 01-Jan-2017 | 28-Feb-2022 | 850,687.38 | ChAdOx technology |
| European Commission | HCR00890 | EU Government | Optimizing a deployable high efficacy malaria vaccine | Hill, Prof. Adrian | 01-Jan-2017 | 28-Feb-2022 | 1,484,179.25 | ChAdOx technology |
| European Commission | HCR00890 | EU Government | Optimizing a deployable high efficacy malaria vaccine | Hill, Prof. Adrian | 01-Jan-2017 | 28-Feb-2022 | 910,006.65 | ChAdOx technology |
| European Commission | HCR00890 | EU Government | Optimizing a deployable high efficacy malaria vaccine | Hill, Prof. Adrian | 01-Jan-2017 | 28-Feb-2022 | 547,709.05 | ChAdOx technology |
| Jenner Vaccine Foundation | HCR00900 | UK Charity (no QR) | Jenner Investigator Allowance | Hill, Prof. Adrian | 01-Jan-2017 | 10-Mar-2018 | 105,000.00 | Not relevant to ChAdOx |
| Name withheld | HCR00920 | UK Industry (QR) | Process Development for manufacturing personalised vaccines | Gilbert, Prof. Sarah | 01-Feb-2017 | 28-Feb-2019 | 399,708.45 | Not relevant to ChAdOx |
| Wellcome Trust | HCR01170 | UK Charity (QR) | Development of a high efficacy vaccine against P.falciparum malaria | Hill, Prof. Adrian | 01-Feb-2017 | 30-Nov-2022 | 0.01 | ChAdOx technology |
| Wellcome Trust | HCR01170 | UK Charity (QR) | Development of a high efficacy vaccine against P.falciparum malaria | Hill, Prof. Adrian | 01-Feb-2017 | 30-Nov-2022 | 3,610,734.99 | ChAdOx technology |
| Wellcome Trust | HCR01170 | UK Charity (QR) | Development of a high efficacy vaccine against P.falciparum malaria | Hill, Prof. Adrian | 01-Feb-2017 | 30-Nov-2022 | 1,554,265.00 | ChAdOx technology |
| Oxford Biomedical Research Centre | HCR00951 | UK Public Sector | BRC3 - Vaccines for Emerging & Endemic Diseases - Year 1 | Hill, Prof. Adrian | 01-Apr-2017 | 31-May-2018 | 63,864.00 | ChAdOx technology |
| Oxford Biomedical Research Centre | HCR00951 | UK Public Sector | BRC3 - Vaccines for Emerging & Endemic Diseases - Year 1 | Hill, Prof. Adrian | 01-Apr-2017 | 31-May-2018 | 568,273.00 | ChAdOx technology |
| Oxford Biomedical Research Centre | HCR00951 | UK Public Sector | BRC3 - Vaccines for Emerging & Endemic Diseases - Year 1 | Hill, Prof. Adrian | 01-Apr-2017 | 31-May-2018 | 279,753.00 | ChAdOx technology |
| European Commission | HCR01110 | EU Government | European Vaccine Research and Development Infrastructure (TRANSVAC2) | Hill, Prof. Adrian | 01-May-2017 | 30-Jun-2022 | 107,313.89 | Not relevant to ChAdOx |
| European Commission | HCR01110 | EU Government | European Vaccine Research and Development Infrastructure (TRANSVAC2) | Hill, Prof. Adrian | 01-May-2017 | 30-Jun-2022 | 414,623.64 | Not relevant to ChAdOx |
| Innovate UK | HCR01100 | UK Public Sector | Preclinical Crimean Congo Haemorrhagic Fever Vaccine Development | Gilbert, Prof. Sarah | 15-May-2017 | 14-Jul-2018 | 350,780.00 | ChAdOx technology |
| Name withheld | HCR01090 | UK Industry (QR) | A phase I study to determine the safety and immunogenicity influenza vaccine MVA-NP+M1, manufactured on the AGE1.CR.pIX noval avian cell | Gilbert, Prof. Sarah | 08-Jun-2017 | 13-Mar-2018 | 63,426.26 | Other vaccine research |
| United States Agency for International Development | HCR01080 | Non-EU Other | Rebecca Ashfield Leidos April 17 | Ashfield, Dr. Rebecca | 14-Jun-2017 | 30-Aug-2019 | 66,925.52 | Other vaccine research |
| Name withheld | HCR01140 | UK Industry (QR) | INVICTUS Main Trial | Gilbert, Prof. Sarah | 09-Aug-2017 | 30-Nov-2019 | 1,698,717.96 | Other vaccine research |
| Name withheld | HCR01140 | UK Industry (QR) | INVICTUS Main Trial | Gilbert, Prof. Sarah | 09-Aug-2017 | 30-Nov-2019 | 130,328.95 | Other vaccine research |
| Name withheld | HCR01230 | UK Industry (QR) | Costs relating to GMP storage of Vaccitech's IMP products | Gilbert, Prof. Sarah | 30-Aug-2017 | 29-Oct-2022 | 98,318.67 | Not relevant to ChAdOx |
| Jenner Vaccine Foundation | HCR01120 | UK Charity (no QR) | Public Engagement - Jenner | Hill, Prof. Adrian | 01-Sep-2017 | 28-Feb-2018 | 22,200.00 | Not relevant to ChAdOx |
| Medical Research Council | HCR01130 | Research Councils | Development and testing of MVA VerOx | Gilbert, Prof. Sarah | 01-Sep-2017 | 30-Apr-2019 | 46,598.47 | Other vaccine research |
| Innovate UK | HCR01240 | UK Public Sector | A Nipah vaccine to eliminate porcine reservoirs and safeguard human health | Gilbert, Prof. Sarah | 01-Sep-2017 | 31-May-2021 | 53,374.03 | ChAdOx technology |
| Medical Research Council | HCR01180 | Research Councils | Clinical Evaluation of "Prime-Target" Immunisation | Hill, Prof. Adrian | 01-Feb-2018 | 30-Jun-2019 | 670,257.42 | Other vaccine research |
| Biotechnology & Biological Sciences Research Council | HCR01190 | Research Councils | A single dose vectored Taenia solium vaccine | Hill, Prof. Adrian | 06-Mar-2018 | 05-May-2019 | 26,800.00 | ChAdOx technology |
| Oxford Biomedical Research Centre | HCR00952 | UK Public Sector | BRC3 - Vaccines for Emerging & Endemic Diseases - Year 2 | Hill, Prof. Adrian | 01-Apr-2018 | 31-May-2019 | 67,895.00 | ChAdOx technology |
| Oxford Biomedical Research Centre | HCR00952 | UK Public Sector | BRC3 - Vaccines for Emerging & Endemic Diseases - Year 2 | Hill, Prof. Adrian | 01-Apr-2018 | 31-May-2019 | 564,350.00 | ChAdOx technology |
| Oxford Biomedical Research Centre | HCR00952 | UK Public Sector | BRC3 - Vaccines for Emerging & Endemic Diseases - Year 2 | Hill, Prof. Adrian | 01-Apr-2018 | 31-May-2019 | 294,005.00 | ChAdOx technology |
| European Commission | HCR01350 | EU Government | Multi-Stage Malaria Vaccine Consortium: field efficacy testing of a multi-stage malaria vaccine MMVC | Hill, Prof. Adrian | 01-Apr-2018 | 30-Nov-2023 | 2,110,089.62 | ChAdOx technology |
| European Commission | HCR01350 | EU Government | Multi-Stage Malaria Vaccine Consortium: field efficacy testing of a multi-stage malaria vaccine MMVC | Hill, Prof. Adrian | 01-Apr-2018 | 30-Nov-2023 | 2,886,824.83 | ChAdOx technology |
| European Commission | HCR01350 | EU Government | Multi-Stage Malaria Vaccine Consortium: field efficacy testing of a multi-stage malaria vaccine MMVC | Hill, Prof. Adrian | 01-Apr-2018 | 30-Nov-2023 | 162,251.10 | ChAdOx technology |
| Engineering & Physical Sciences Research Council | HCR01420 | Research Councils | The Future Vaccine Manufacturing Research Hub | Gilbert, Prof. Sarah | 01-Apr-2018 | 31-May-2021 | 1,021,747.04 | Not relevant to ChAdOx |
| Engineering & Physical Sciences Research Council | HCR01420 | Research Councils | The Future Vaccine Manufacturing Research Hub | Gilbert, Prof. Sarah | 01-Apr-2018 | 31-May-2021 | 1,590,456.91 | Not relevant to ChAdOx |
| Innovate UK | HCR01210 | UK Public Sector | An economically viable vaccine for CCHF virus | Gilbert, Prof. Sarah | 10-Apr-2018 | 30-Sep-2020 | 25,000.00 | Not relevant to ChAdOx |
| European Commission | HCR01260 | EU Government | Addressing the dual emerging threats of African Swine Fever and lumpy skin disease in Europe | Gilbert, Prof. Sarah | 01-Jun-2018 | 31-Jul-2023 | 87,363.60 | Other vaccine research |
| Medical Research Council | HCR01280 | Research Councils | Development of a vaccine to prevent Crimean-Congo haemorrhagic fever virus mediated disease. | Gilbert, Prof. Sarah | 01-Aug-2018 | 31-Oct-2020 | 47,786.83 | Not relevant to ChAdOx |
| Innovate UK | HCR01340 | UK Public Sector | CCHF Vaccine manufacturing and First in Human Clinical Trial | Gilbert, Prof. Sarah | 01-Sep-2018 | 31-May-2021 | 1,999,524.27 | ChAdOx technology |
| British Medical Association | H5R01110 | UK Industry (QR) | Delineating the role of the human leukocyte antigen locus in susceptibility to rheumatic heart disease in Oceania and South Asia | Hill, Prof. Adrian | 01-Sep-2018 | 31-Oct-2021 | 29,960.00 | Other vaccine research |
| British Medical Association | H5R01110 | UK Industry (QR) | Delineating the role of the human leukocyte antigen locus in susceptibility to rheumatic heart disease in Oceania and South Asia | Hill, Prof. Adrian | 01-Sep-2018 | 31-Oct-2021 | 10,000.00 | Other vaccine research |
| British Medical Association | H5R01110 | UK Industry (QR) | Delineating the role of the human leukocyte antigen locus in susceptibility to rheumatic heart disease in Oceania and South Asia | Hill, Prof. Adrian | 01-Sep-2018 | 31-Oct-2021 | 10,000.00 | Other vaccine research |
| Coalition for Epidemic Preparedness Innovations | HCR01320 | Non-EU Other | PADOVAX- MERS | Gilbert, Prof. Sarah | 01-Oct-2018 | 30-Nov-2023 | 0.01 | ChAdOx technology |
| Coalition for Epidemic Preparedness Innovations | HCR01320 | Non-EU Other | PADOVAX- MERS | Gilbert, Prof. Sarah | 01-Oct-2018 | 30-Nov-2023 | 4,901,551.72 | ChAdOx technology |

| | | | | | | | | |
|---|---|---|---|---|---|---|---|---|
| Coalition for Epidemic Preparedness Innovations | HCR01320 | Non-EU Other | PADOVAX- MERS | Gilbert, Prof. Sarah | 01-Oct-2018 | 30-Nov-2023 | 0.00 | ChAdOx technology |
| Coalition for Epidemic Preparedness Innovations | HCR01320 | Non-EU Other | PADOVAX- MERS | Gilbert, Prof. Sarah | 01-Oct-2018 | 30-Nov-2023 | 0.00 | ChAdOx technology |
| Coalition for Epidemic Preparedness Innovations | HCR01320 | Non-EU Other | PADOVAX- MERS | Gilbert, Prof. Sarah | 01-Oct-2018 | 30-Nov-2023 | 178,472.28 | ChAdOx technology |
| Coalition for Epidemic Preparedness Innovations | HCR01320 | Non-EU Other | PADOVAX- MERS | Gilbert, Prof. Sarah | 01-Oct-2018 | 30-Nov-2023 | 7,530.52 | ChAdOx technology |
| Coalition for Epidemic Preparedness Innovations | HCR01320 | Non-EU Other | PADOVAX- MERS | Gilbert, Prof. Sarah | 01-Oct-2018 | 30-Nov-2023 | 787,489.00 | ChAdOx technology |
| Coalition for Epidemic Preparedness Innovations | HCR01320 | Non-EU Other | PADOVAX- MERS | Gilbert, Prof. Sarah | 01-Oct-2018 | 30-Nov-2023 | 906,850.08 | ChAdOx technology |
| Coalition for Epidemic Preparedness Innovations | HCR01320 | Non-EU Other | PADOVAX- MERS | Gilbert, Prof. Sarah | 01-Oct-2018 | 30-Nov-2023 | 2,615,395.66 | ChAdOx technology |
| Coalition for Epidemic Preparedness Innovations | HCR01321 | Non-EU Other | PADOVAX-Nipah | Gilbert, Prof. Sarah | 01-Oct-2018 | 28-Feb-2021 | 0.01 | ChAdOx technology |
| Coalition for Epidemic Preparedness Innovations | HCR01321 | Non-EU Other | PADOVAX-Nipah | Gilbert, Prof. Sarah | 01-Oct-2018 | 28-Feb-2021 | 115,718.23 | ChAdOx technology |
| Coalition for Epidemic Preparedness Innovations | HCR01321 | Non-EU Other | PADOVAX-Nipah | Gilbert, Prof. Sarah | 01-Oct-2018 | 28-Feb-2021 | 0.00 | ChAdOx technology |
| Coalition for Epidemic Preparedness Innovations | HCR01321 | Non-EU Other | PADOVAX-Nipah | Gilbert, Prof. Sarah | 01-Oct-2018 | 28-Feb-2021 | 475,936.04 | ChAdOx technology |
| Coalition for Epidemic Preparedness Innovations | HCR01321 | Non-EU Other | PADOVAX-Nipah | Gilbert, Prof. Sarah | 01-Oct-2018 | 28-Feb-2021 | 281,021.66 | ChAdOx technology |
| Coalition for Epidemic Preparedness Innovations | HCR01321 | Non-EU Other | PADOVAX-Nipah | Gilbert, Prof. Sarah | 01-Oct-2018 | 28-Feb-2021 | 538,467.81 | ChAdOx technology |
| Coalition for Epidemic Preparedness Innovations | HCR01322 | Non-EU Other | Potent, scalable adenoviral vectored vaccines against Lassa | Gilbert, Prof. Sarah | 01-Oct-2018 | 31-Dec-2020 | 0.01 | ChAdOx technology |
| Coalition for Epidemic Preparedness Innovations | HCR01322 | Non-EU Other | Potent, scalable adenoviral vectored vaccines against Lassa | Gilbert, Prof. Sarah | 01-Oct-2018 | 31-Dec-2020 | 84,921.38 | ChAdOx technology |
| Coalition for Epidemic Preparedness Innovations | HCR01322 | Non-EU Other | Potent, scalable adenoviral vectored vaccines against Lassa | Gilbert, Prof. Sarah | 01-Oct-2018 | 31-Dec-2020 | 443,988.95 | ChAdOx technology |
| Coalition for Epidemic Preparedness Innovations | HCR01322 | Non-EU Other | Potent, scalable adenoviral vectored vaccines against Lassa | Gilbert, Prof. Sarah | 01-Oct-2018 | 31-Dec-2020 | 279,577.40 | ChAdOx technology |
| Coalition for Epidemic Preparedness Innovations | HCR01322 | Non-EU Other | Potent, scalable adenoviral vectored vaccines against Lassa | Gilbert, Prof. Sarah | 01-Oct-2018 | 31-Dec-2020 | 481,339.45 | ChAdOx technology |
| Bill & Melinda Gates Foundation | HCR01530 | Non-EU Charity (QR) | R21 Variant Malaria Vaccine Candidate | Hill, Prof. Adrian | 01-Jan-2019 | 31-Jul-2021 | 387,304.92 | Other vaccine research |
| Oxford Biomedical Research Centre | HCR00953 | UK Public Sector | BRC3 - Vaccines for Emerging & Endemic Diseases - Year 3 | Hill, Prof. Adrian | 01-Apr-2019 | 31-May-2020 | 68,147.00 | ChAdOx technology |
| Oxford Biomedical Research Centre | HCR00953 | UK Public Sector | BRC3 - Vaccines for Emerging & Endemic Diseases - Year 3 | Hill, Prof. Adrian | 01-Apr-2019 | 31-May-2020 | 569,583.00 | ChAdOx technology |
| Oxford Biomedical Research Centre | HCR00953 | UK Public Sector | BRC3 - Vaccines for Emerging & Endemic Diseases - Year 3 | Hill, Prof. Adrian | 01-Apr-2019 | 31-May-2020 | 303,265.00 | ChAdOx technology |
| Medical Research Council | HCR01490 | Research Councils | Broad and effective protection against influenza achieved by viral vectored vaccines | Gilbert, Prof. Sarah | 01-May-2019 | 30-Jun-2022 | 768,110.18 | Not relevant to ChAdOx |
| Medical Research Council | HCR01490 | Research Councils | Broad and effective protection against influenza achieved by viral vectored vaccines | Gilbert, Prof. Sarah | 01-May-2019 | 30-Jun-2022 | 839,580.80 | Not relevant to ChAdOx |
| Sepsis Research (FEAT) SCIO | H5R01240 | UK Charity (no QR) | Finding effective targets to modify the host response to sepsis | Hill, Prof. Adrian | 01-Dec-2019 | 31-Mar-2020 | 8,900.00 | Other vaccine research |
